# Clonal haematopoiesis without identified genetic drivers: insights from analyses of 407,512 individuals

**DOI:** 10.64898/2026.05.17.26353391

**Authors:** Sean Wen, Rafael Campos, Marcin Karpinski, Rahul Sharma, Veselin Manojlovic, Sri V. V. Deevi, Sean O’Dell, Xiaoyin Li, Fengyuan Hu, Jared O’Connell, Abhishek Nag, Karine Megy, Stewart MacArthur, Sebastian Wasilewski, Xueqing Zoe Zou, Dimitrios Vitsios, Quanli Wang, Slavé Petrovski, Andrew R. Harper, Margarete A. Fabre, George S. Vassiliou, Jonathan Mitchell

## Abstract

Clonal haematopoiesis (CH) becomes ubiquitous as humans age. The role of somatic driver mutations in its development has been studied widely, but little is known about CH without identified genetic drivers, also known as “CH with unknown drivers” (CH-UD). A fundamental unresolved question is whether CH-UD is driven by undiscovered somatic genetic drivers or by other cell-heritable traits. Here, to investigate this, we develop a new machine learning classifier to improve CH-UD detection from whole-genome sequencing data. After excluding 77,885 individuals with previously documented driver CH or mosaic chromosomal alterations (mCA), we applied our classifier to 407,512 UK Biobank participants and identified 26,963 (6.6%) with CH-UD. A genome-wide association study (GWAS) of common germline variants identified 31 polymorphic loci associated with predisposition to CH-UD. Of these, 25 were associated with other forms of CH at genome-wide significance. Linkage Disequilibrium Score Regression analyses revealed an unexpectedly high genetic correlation (r_g_=0.794) between CH-UD and non-*DNMT3A* driver CH, indicative of a remarkable overlap between the genetic aetiologies of the two phenomena. Analysis of 2,941 plasma protein measurements in 47,757 individuals revealed that TCL1A was the most significantly elevated plasma protein in CH-UD, mirroring the finding that the *TCL1A* locus was in the top two most significant associations of CH-UD GWAS and *TET2*-CH and *ASXL1*-CH GWAS, the two most common forms of non-DNMT3A-CH. Furthermore, TCL1A plasma levels rose steadily with age even in those without detectable CH, particularly among carriers of the common *TCL1A* risk variant (rs2887399-G), potentially via stochastic promoter demethylation as described in *TET2*-CH and *ASXL1*-CH. Phenome-wide association analysis of 13,225 binary and 1,682 quantitative traits revealed that, similarly to non-*DNMT3A*-CH, CH-UD was significantly associated with several malignant (haematological and solid organ) and non-malignant (including cardiovascular and renal) diseases. Our findings reveal striking genetic and phenotypic similarities between CH-UD and non-*DNMT3A* driver CH, including a strong dependence on TCL1A, a protein recently found to inhibit DNA methylation. Collectively, these observations propose that CH-UD develops through selection acting on ageing-associated epigenetic changes that mirror those of non-*DNMT3A*-CH, but without the need for somatic genetic drivers.

## Introduction

Clonal haematopoiesis (CH) is a pre-leukaemic condition characterised by the clonal expansion of a haematopoietic stem cell (HSC) and its progeny. Large-scale genetic epidemiology studies have identified multiple determinants of CH risk, including common and rare germline polymorphisms^1–3^, smoking^1–3^, ancestry^4^, and inherited leukocyte telomere length (LTL)^5^. Beyond its association with haematological malignancies, CH is also associated with increased risk of several non-haematological pathologies such as prostate and lung cancer^6^, cardiovascular^7^, pulmonary^8^, renal^9^, and liver^10^ diseases.

In most population-scale CH studies, somatic clones were identified by the presence of somatic driver mutations in leukaemia-associated genes (driver gene-mutant CH or henceforth “driver CH” for conciseness) and chromosome-scale gains and losses (mosaic chromosomal alterations, mCA). More recently, large-scale whole-genome sequencing (WGS) studies of blood DNA have used passenger mutations to detect CH clones, without relying on the detection of known driver mutations^11–15^. Intriguingly, such studies revealed that a large fraction of individuals with CH clones identified in this way had no detectable known driver variants. Separately, phylogenetic analyses and DNA methylation-based tracing of large numbers of haematopoietic stem and progenitor cells (HSPCs) from single individuals provided further evidence for the ubiquitous development after the 7^th^ decade of life of “CH without identified genetic drivers” or “CH with unknown drivers” ^16–20^ (hereafter referred to as “CH-UD” for consistency with published abbreviations^21^). However, many epidemiological studies either aggregated all CH classes (driver CH, mCA, and CH-UD) into a single category^11,12^ or treated CH or CH-UD as a quantitative trait based on mutation burden (number of passenger variants)^13,14^, which precluded the deep interrogation of CH-UD as a distinct entity.

To overcome the technical challenge of distinguishing somatic from germline variants in the absence of a matched ‘normal’ sample, previous population scale approaches for identifying CH-UD relied on enriching for somatic passenger mutations by filtering for variants present only once in the entire population of large cohorts (“singletons”)^11,12,15^. Here, to improve CH-UD detection, we develop a novel machine learning (ML) classifier that incorporates multiple additional features of singletons. By applying our ML classifier to whole-genome sequenced UK Biobank (UKB) participants, in whom driver CH and mCA were previously identified^4,22^, we were able to robustly identify CH-UD, determine its causes and consequences and contrast these to other forms of CH in the same population. Our findings uncover the causes, consequences, and genetic associations of CH-UD at unprecedented scale. Furthermore, we reveal a striking degree of genetic correlation between CH-UD and non-*DNMT3A* driver CH and provide evidence suggesting that most CH-UD cases truly lack somatic genetic drivers and are primarily epigenetically driven.

## Results

### Detection of CH-UD

We first obtained the driver CH and mCA status of UKB participants as determined in previous studies using whole-exome sequencing (WES) and genotyping array data, respectively^4,22^ (Supplementary Table 1). We then used WGS data to identify putative passenger mutations, defined as genetic variants which occurred in only a single individual (“singletons”) with variant allele frequency (VAF) <25% (see Methods; Supplementary Figure 1) from 407,512 individuals of European descent (Supplementary Figures 2 and 3, Supplementary Table 2, and Supplementary Notes 1).

Starting with this set of singletons, we applied a random forest classifier to identify individuals with CH-UD among those without driver CH or mCA (Figure 1). We trained and tested our classifier using individuals with driver CH (positive class; n = 15,848) and individuals 44 years old or younger without driver CH or mCA (negative class; n = 36,749). To confidently identify those with CH-UD, the ML classifier considered the following features for each individual with one or more singletons: number of singletons, point mutation type (single nucleotide substitution, deletion, insertion), nucleotide context (C>A, C>G, C>T, T>A, T>C, T>G), chromatin accessibility, and mutation tolerance score. In a univariate analysis, each of these features showed significant differences between positive class versus negative class (Supplementary Figure 4 and Supplementary Notes 2). We also noted variable sequencing metrics across the different WGS batches (Supplementary Figure 5). Therefore, we trained, tested, and applied the classifier separately on each WGS batch.

**Figure 1:**
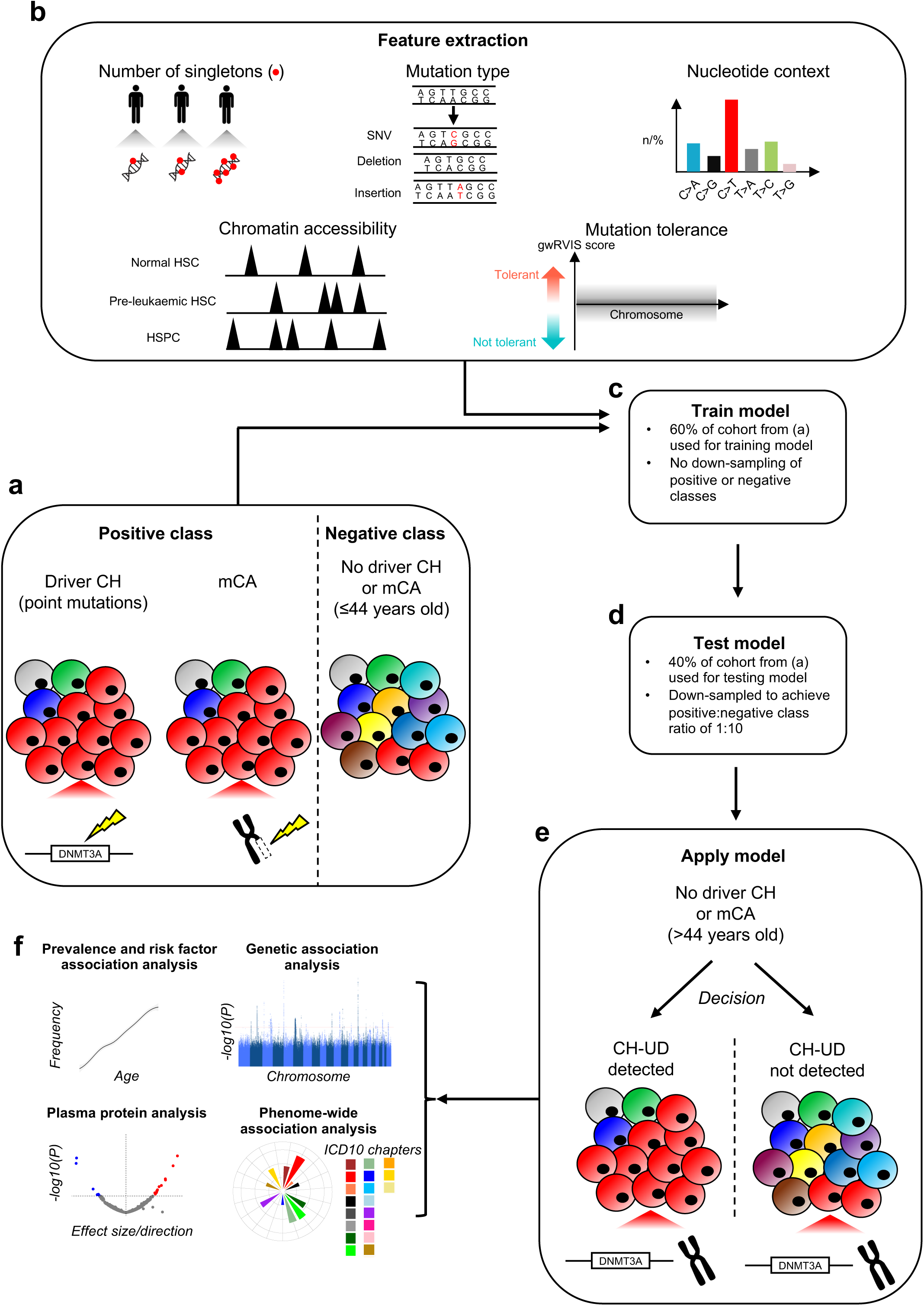
Overview on the machine learning classifier for inferring CH-UD and downstream applications. **a,** Positive control set was defined as individuals with identifiable driver CH (such as *DNMT3A*) or mCA. Negative control set was defined as individuals ≤44 years old without any identifiable driver CH or mCA. **b,** Features of singletons were extracted, namely number of singletons, type of point mutation, nucleotide context (both single-base context, as shown here, and also tri-nucleotide context), location of singletons in opened or closed chromatic regions as defined by ATAC-sequencing inferred peaks, and mutation tolerance score at singleton sites. **c,** Positive and negative control sets from **(a)** and singleton features from **(b)** were used to train the machine learning (ML) classifier. Specifically, 60% of the control set from **(a)** were used to train the classifier. The classifier was trained on three different positive control sets, namely driver CH, mCA, or combined driver CH and mCA. **d,** The trained classifier was subsequently tested using 40% of the control set from **(a)**. The positive and negative control sets were down-sampled to yield 1:10 ratio to simulate the approximate the reported population frequency of driver CH. The ML classifiers assessed here were random forest, XGBoost, SVM, logistic regression. **e,** The trained classifier was then applied to predict the presence or absence of CH among individuals >44 years old without any identifiable driver CH or mCA. **f,** CH-UD was used as a phenotype for downstream association analyses including risk factor association analysis, genetic association analysis, differential plasma protein level analysis, and phenome-wide association analysis. HSC, haematopoietic stem cell, HSPC, haematopoietic stem and progenitor cell, mCA, mosaic chromosomal alterations. SVM, support vector machine.

When applied to the test set consisting of driver CH or mCA, our classifier demonstrated an area under the curve (AUC) of 63.1%, a sensitivity of 29.9%, and a negative predictive value (NPV) of 93.2% (Supplementary Figures 6a-e and Supplementary Notes 3). We observed clone size-dependent performance, with improved detection of larger clones (an AUC of 81.5%, a sensitivity of 66.9%, and a NPV of 99.2% for cell fraction ≥0.2; versus 54.5%, 12.7% and 95.6%, respectively, for cell fraction <0.1). When applied to the test set consisting of driver CH only, our classifier demonstrated an AUC of 71.1%, a sensitivity of 46.0%, and a NPV of 98.3% with improved detection for larger clones (an AUC of 83.7%, a sensitivity of 71.2%, a NPV of 99.6% for cell fraction ≥0.2; versus 54.8%, 13.3% and 99.5%, respectively, for cell fraction <0.1) Applied to the full cohort, the classifier demonstrated a positive association between CH-UD frequency and age (Supplementary Figures 6f-k).

We further validated our approach by: i) demonstrating that singletons were enriched for variants of somatic origin (evidenced by an increasing singleton burden with age; Supplementary Figures 7a-d), ii) showing enrichment of age- and HSC-related mutational signatures but depletion of sequencing artefact-related mutational signatures among singletons (Supplementary Figures 7e-j), iii) revealing increasing singleton-inferred clone size with age (Supplementary Figures 8a-e), iv) recapitulating the previously reported association between LTL and gene-specific driver CH risk^23^ (Supplementary Figures 8f and g), v) recapitulating previously reported relative fitness (singleton burden/PACER^14^) of CH driven by different driver genes (Supplementary Figures 9a-c), and vi) recapitulating previously reported genomic loci associated with singleton burden, namely *SMC4*, *TERT*, *TCL1A*, and *NRIP1*^13^ (Supplementary Figures 9d).

### Frequency of CH-UD

CH-UD was identified in 26,963 of 407,512 individuals (6.62%), making this the second most common class of CH in UKB, after mosaic loss of Y chromosome (mLOY) (Figure 2a). The prevalence of CH-UD demonstrated a strong association with advancing age (Figure 2b). For example, we observed a 9-fold higher frequency in 70-compared to 40-year-olds (9.86% versus 1.10%, respectively; *P*_chi-square_ < 1.0 × 10^−16^). Notably, the frequency of CH-UD was higher than that of driver CH across all age groups. Also, CH-UD was more prevalent in females than males (7.83% versus 5.19%, respectively; *P*_chi-square_ < 1.0 × 10^−16^; Figures 2c and d).

**Figure 2:**
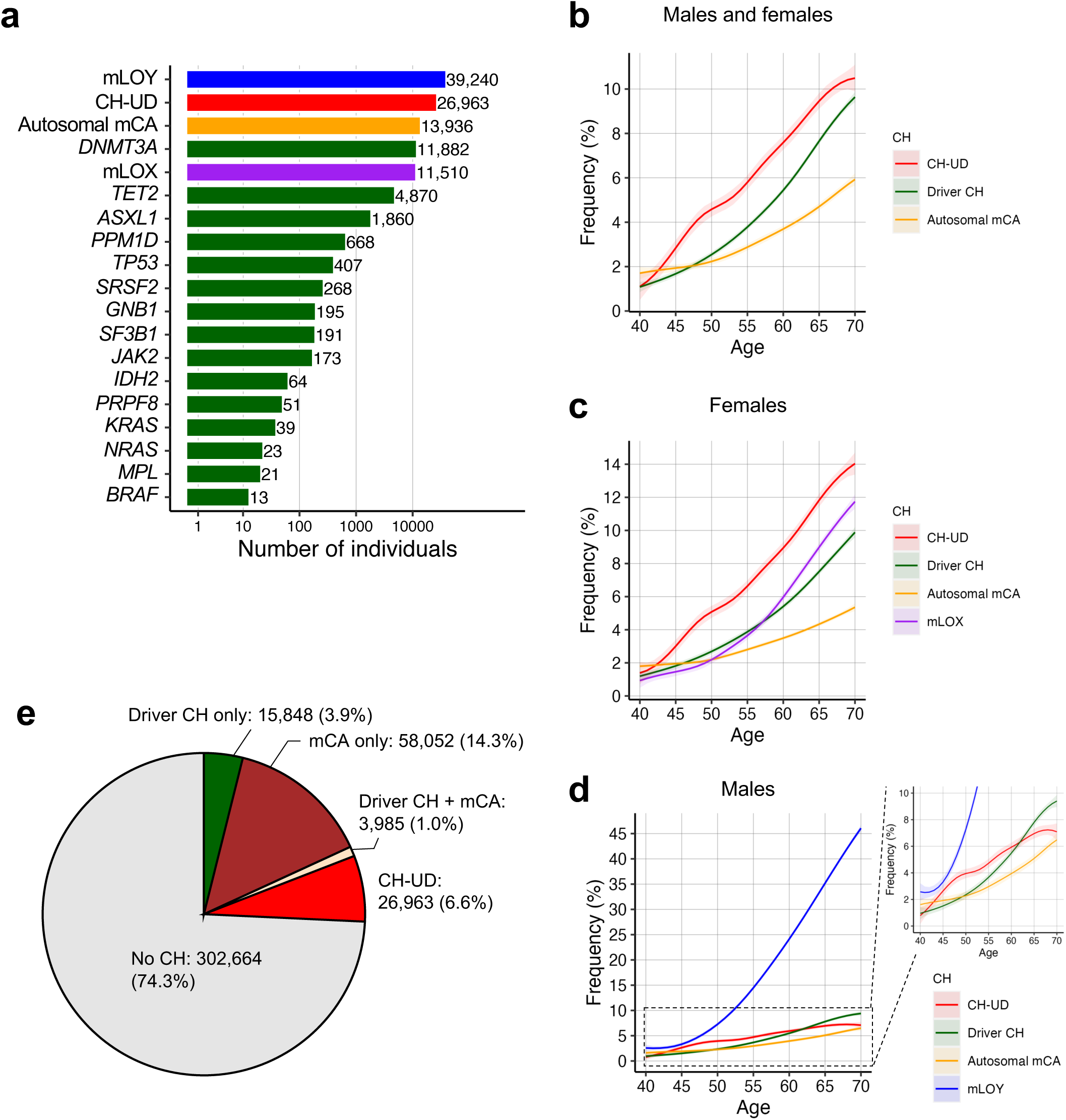
Frequency of CH. **a,** Frequency of CH-UD, driver CH (stratified by gene-specific driver CH), and mCA (stratified into mosaic loss of autosome, chrX, and chrY). **b-d,** Frequency by age for CH-UD, driver CH, and mCA among all samples **(b)**, females only **(c)**, and males only **(d)**. The center line represents the fitted values from the general additive model with P-spline smooth class while the shaded regions represent the lower and upper bounds of the 95% CIs of the fitted values **(a-d)**. **e,** Frequency and corresponding proportion of individuals with CH-UD, driver CH (without mCA), or mCA (without driver CH), and driver CH and mCA detected in the same individual, and individuals without any detectable CH.

The detection of CH-UD increased the overall CH frequency estimates from 19.1% (driver CH and mCA) to 25.7% (driver CH, mCA, and CH-UD) in UKB (Figure 2e). The increase was more pronounced in females (12.4% to 20.2%) than in males (27.0% to 32.2%), corresponding to an approximately threefold greater relative increase in females. Collectively, these findings reveal that more than 1 in 4 UKB participants had at least one form of CH detectable by WES, WGS or SNP array.

### Risk factor association analyses

To determine the extent to which established CH risk factors predispose to CH-UD, we looked for potential associations using multivariable logistic regression models including key covariates, namely age, sex, smoking status, and the first four genetic principal components (PCs). For every year increase in age, the odds of CH-UD increased by 8.5% (OR [95% CI] = 1.085 [1.083,1.087], *P* < 1.0 × 10^−16^; Figure 3a and Supplementary Table 3), an effect comparable to that seen with driver CH.

**Figure 3:**
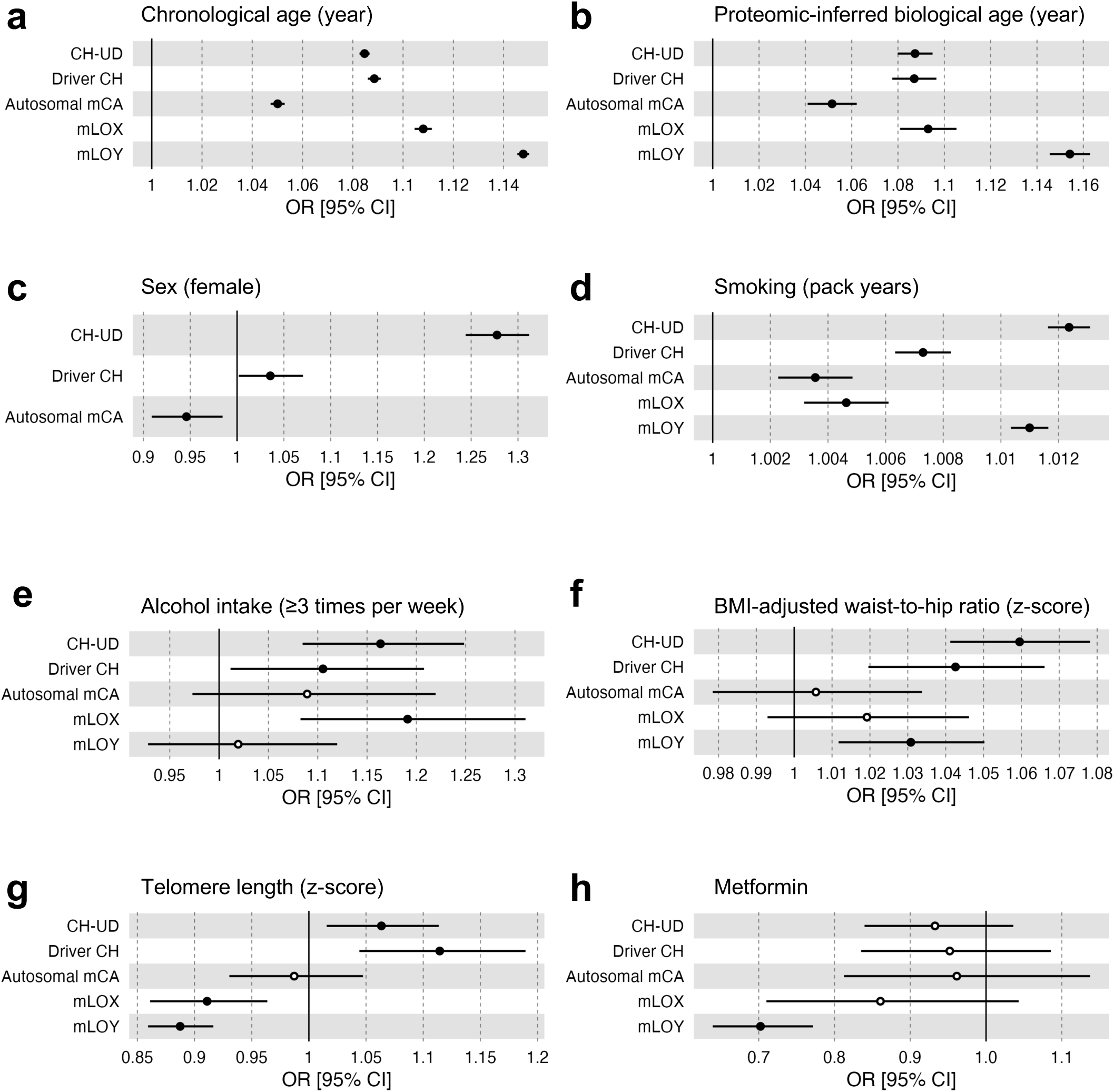
Assessment of established CH risk factors with CH-UD, driver CH, and mCA. **a,** Association between chronological age and CH. Odds ratio derived from logistic regression with CH as the outcome and chronological age as the main predictor with sex, smoking, and first four *peddy*-inferred genetic principal components (PCs) as co-variates. **b,** Association between biological age, as inferred using plasma proteins, and CH. Odds ratio derived from logistic regression with CH as the outcome and biological age as the main predictor with sex, smoking, Olink batch, time between measurement and sampling plasma protein, and first four *peddy*-inferred genetic PCs as co-variates. **c,** Association between sex and CH. Odds ratio derived from logistic regression with CH as the outcome and sex as the main predictor (females relatives males) with age, smoking, and first four *peddy*-inferred genetic PCs as co-variates. **d,** Association between smoking and CH. Odds ratio derived from logistic regression with CH as the outcome and smoking as the main predictor with age, sex, and first four *peddy*-inferred genetic PCs as co-variates. Smoking is represented in pack years where it reflects the lifetime exposure to tobacco exposure by multiplying packs smoked per day by years smoked. **e,** Association between alcohol intake and CH. Odds ratio derived from logistic regression with CH as the outcome and alcohol intake as the main predictor (≥3 times weekly relative to never drinkers) with age, sex, smoking, and first four *peddy*-inferred genetic PCs as co-variates. **f,** Association between waist-to-hip ratio and CH. BMI was regressed out from waist-to-hip ratio in a linear regression model to obtain the residuals. The residuals were subsequently rank inverse transformed, and then centred and scaled. Odds ratio here derived from logistic regression with CH as the outcome and BMI-adjusted waist-to-hip ratio as the main predictor with age, sex, smoking, and first four *peddy*-inferred genetic PCs as co-variates. **g,** Association between leukocyte telomere length (LTL) and CH (related to Supplementary Figure 8f). LTL was derived from the first principal component (PC1) of qPCR-inferred telomere length and TelSeq/WGS-inferred telomere length (Burren *et al*.), and then centred and scaled. Odds ratio derived from logistic regression with CH as the outcome and telomere length PC1 as the main predictor with age, sex, smoking, and first four *peddy*-inferred genetic PCs as co-variates. Only small CH clones, defined as <10^th^ quantile of singleton-derived cell fraction, were included for analysis. **h,** Association between pre-recruitment metformin intake and CH. Individuals on metformin who were also on other diabetic medications (insulin, sulphonylureas, thiazolidinediones, meglitinides or acarbose) were excluded. Odds ratio derived from logistic regression with CH as the outcome and metformin intake as the main predictor with age, sex, smoking, first four *peddy*-inferred genetic PCs, and BMI-adjusted waist-to-hip ratio as co-variates. Solid circles represent significant associations (*P* < 0.05) while hollow circles represent non-significant associations (*P* ≥ 0.05).

We next hypothesised that CH may be more strongly associated with biological than chronological age^24^. To test this, we determined the plasma proteome-derived biological age of 37,934 individuals (see Methods, Supplementary Figure 10 and Supplementary Table 4) and found that while CH-UD risk increased with this estimate of biological age (OR [95% CI] = 1.087 [1.080,1.095], *P* < 1.0 × 10^−16^; Figure 3b), there was no association with “accelerated aging”, defined as biological minus chronological age (OR [95% CI] = 1.01 [0.99,1.03], *P* = 0.31).

In addition to age, CH-UD was strongly associated with female sex, with a much larger effect size than was observed for driver CH (CH-UD: OR [95% CI] = 1.28 [1.24, 1.31], *P* < 1.0 × 10^−16^; driver CH: OR [95% CI] = 1.036 [1.002, 1.070], *P* = 0.039; Figure 3c).

Further analysis of known CH-related lifestyle and biological risk factors^2,3,25–28^ revealed several significant associations with CH-UD. Smoking in pack years showed a highly significant association with increased CH-UD risk (OR [95% CI] = 1.0124 [1.0116,1.0131], *P* = 1.0 × 10^−16^), beyond that observed for driver CH (OR [95% CI] = 1.0073 [1.0063,1.0083], *P* = 9.33 × 10^−50^; Figure 3d). CH-UD risk was also associated with drinking alcohol ≥3 times weekly (OR [95% CI] = 1.17 [1.09,1.25], *P* = 2.37 × 10^−5^; Figure 3e), central adiposity (as defined by waist-to-hip ratio; OR [95% CI] = 1.06 [1.04,1.08], *P* = 7.95 × 10^−11^; Figure 3f) and longer LTL (OR [95% CI] = 1.06 [1.02,1.11], *P* = 8.90 × 10^−3^; Figure 3g).

Mendelian randomisation (see Methods) supported causal relationships between CH-UD risk and smoking (cigarettes per day; beta [95%CI] = 0.24 [0.043,0.45], *P* = 0.018), and CH-UD risk and longer LTL (beta [95%CI] = 0.32 [0.15,0.50], *P* = 2.90 × 10^−4^; Extended Data Figures 1a-c and Supplementary Table 5).

Recent evidence has shown a protective effect for the commonly prescribed drugs metformin^27,29,30^ and statins^28^ against specific CH subtypes, and we examined their association with CH-UD here, including additional adjustment for body mass index (BMI)-adjusted waist-to-hip ratio. We observed reduced frequency of *DNMT3A* R882-CH amongst those taking metformin (OR [95% CI] = 0.39 [0.19,0.82], *P* = 0.013) and statins (OR [95% CI] = 0.80 [0.66,0.96], *P* = 0.019), but did not identify an association of CH-UD prevalence with either drug (metformin: OR [95% CI] = 0.93 [0.84,1.04], *P* = 0.19; statins: OR [95% CI] = 1.02 [0.98,1.06], *P* = 0.27; Figure 3h).

Given the strong association between sex and CH-UD, we investigated the impact of sex on the other risk factors amongst CH-UD versus non-CH individuals. We observed no sex-specific differences for chronological age, proteomics-inferred biological age, alcohol intake, central adiposity or LTL between CH-UD versus non-CH individuals. However, we did observe sex-specific differences for smoking between CH-UD versus non-CH individuals (sex:smoking interaction; *P* = 2.23 × 10^−9^; Supplementary Table 6). Specifically, the impact of smoking burden (pack years) on CH-UD risk was higher among females than males (females: OR [95% CI] = 1.015 [1.014,1.016], *P* < 1.0 × 10^−16^; males: OR [95% CI] = 1.010 [1.009,1.011], *P* < 1.0 × 10^−16^).

### Heritability and genetic correlation analysis

Using linkage disequilibrium score regression (LDSC) we estimated the SNP-based heritability of CH (Extended Data Figure 2). Our narrow-sense heritability estimates for driver CH (1.26%), autosomal mCA (0.16%), mLOX (1.27%), and mLOY (10.7%) were consistent with previous heritability estimates for driver CH (0.78-3.57)^2,31^, autosomal mCA (0.30%)^31^, mLOX (1.34%)^31^, and mLOY (10.9%)^31^. We estimated the heritability of CH-UD to be 2.5%. We observed inherited genetic correlation of 0.807 between CH-UD and driver CH (*P* = 5.10 × 10^−23^) and 0.269 with mLOY (*P* = 7.00 × 10^−4^; Figure 4a). Notably, the genetic correlation between CH-UD was stronger with non-*DNMT3A* CH (0.794, *P* = 7.60 × 10^−6^) compared to *DNMT3A* CH (0.663, *P* = 8.65 × 10^−15^).

**Figure 4:**
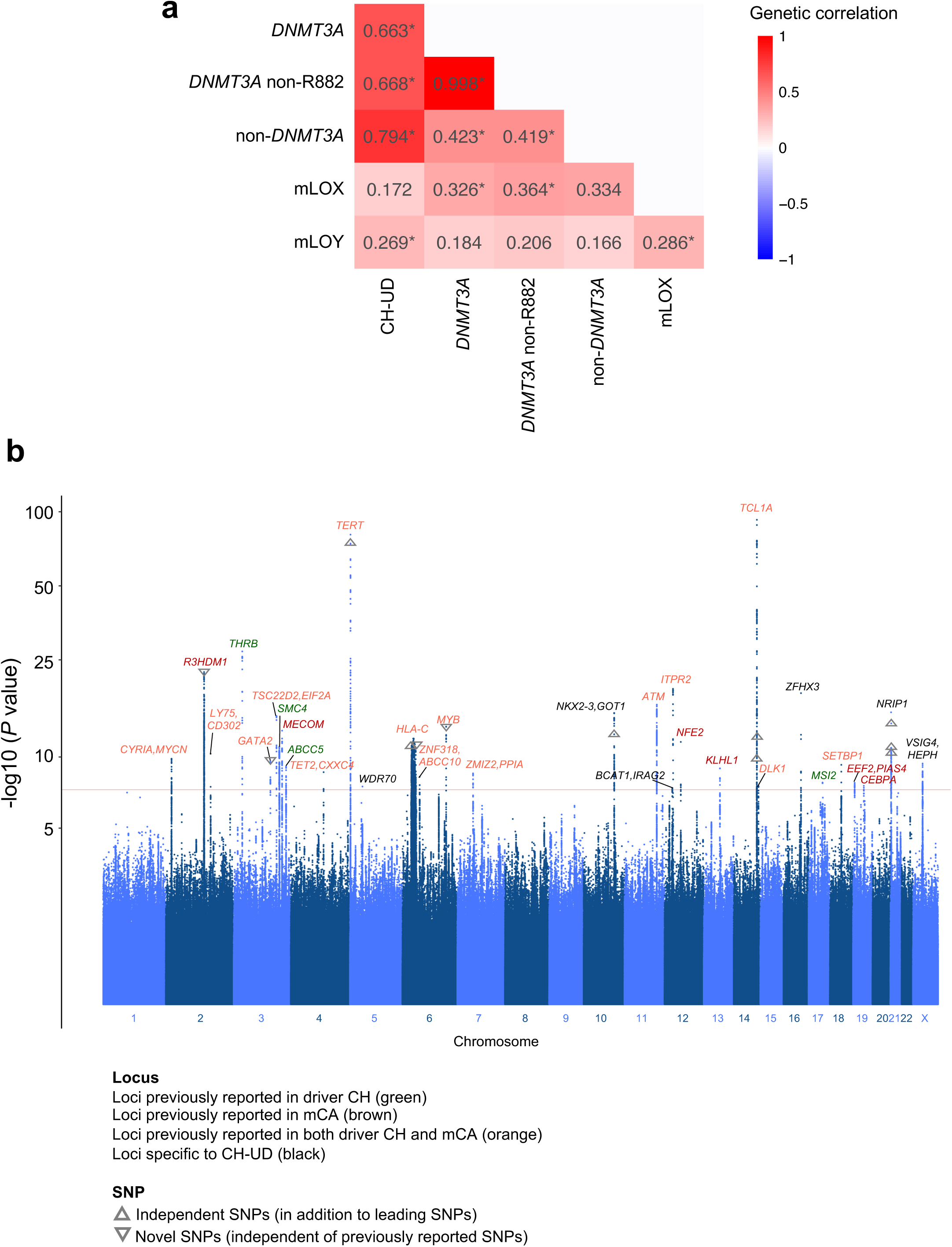
Genome-wide association study (GWAS) of CH-UD and genetic correlation between the different CH subtypes. **a,** Manhattan plot representing the common germline variants included for CH-UD GWAS. *P* values on the y-axis were derived from METAL software. GWAS was ran on each WGS batch separately using Firth logistic regression implemented using SAIGE software, and then meta-analysed using METAL. For SAIGE, we specified age, sex, smoking, *peddy*-inferred European probability, and first ten genetic principal components as co-variates. In total, 19 loci were reported in driver CH GWAS from our study or the literature (orange and green), 21 loci were reported in mCA (autosomal, mLOX or mLOY) GWAS from our study or the literature (orange and brown), and six loci were specific to CH-UD GWAS in this study (black). Eight independent SNPs, in addition to the leading SNPs, were identified using COJO (up-right triangles). Within loci previously reported in driver CH or mCA GWAS, four SNPs independent of previously reported SNPs were identified from conditional analysis (upside-down triangles). **b,** Genetic correlation computed using LDSC software. Only CH subtypes with statistically significant heritability estimates (Extended Data Figure 2) were included for analysis here. * *P* < 0.05. COJO, conditional & joint association analysis; LDSC, linkage disequilibrium score regression; WGS, whole-genome sequencing.

### Common germline variants and CH-UD risk

To determine common germline variants associated with CH-UD, we performed a genome-wide association study (GWAS) comparing 25,187 CH-UD cases to 296,490 controls without CH. This identified 31 loci associated with CH-UD at genome-wide significance (*P* < 5 × 10^−8^; Figure 4b, Supplementary Tables 7 and 8). In addition to the leading SNPs for each locus, conditional analysis identified another 8 independent SNPs across 5 of the loci (Supplementary Table 9).

Comparative analysis revealed both shared and distinct inherited genetic architectures across CH subtypes. Of the 31 loci associated with CH-UD, 15 were also associated with both driver CH (overall driver CH and/or gene-specific driver CH) and mCA (autosomal mCA, mLOX, or mLOY), four with driver CH-only and six with mCA-only (Supplementary Figures 11-13 and Supplementary Tables 10 and 11). At loci previously reported to be associated with driver CH or mCA, we identified four SNPs which remained significantly associated with CH-UD after conditioning on the previously reported SNPs (Supplementary Table 12).

The most significant CH-UD association was at the *TCL1A* locus, where a common *TCL1A* promoter variant (rs2887399; G allele frequency = 0.79) conferred increased risk (OR [95% CI] = 1.34 [1.30,1.37], *P* = 1.85 × 10^−93^). This contrasts with most other forms of CH, where the strongest associations have been observed with *TERT*^1–4^. As previously reported, the rs2887399 G allele is associated with an increased risk of *TET2-* and *ASXL1-*CH but a decreased risk of *DNMT3A-*CH^4,14^. This mirrors our earlier observation on the stronger genetic correlation between CH-UD with non-*DNMT3A-*CH compared to *DNMT3A-*CH.

Six loci were specific to CH-UD at the genome-wide significance threshold. Of these, four (*NKX2-3/GOT1*, *ZFHX3*, *NRIP1*, and *VSIG4/HEPH*) were previously reported in ‘barcode-CH’ (defined as CH inclusive of CH-UD, driver CH and mCA^12^), and two (*WDR70* and *BCAT1/IRAG2*) not previously linked to CH-UD. Notably, both *NRIP1* and *NKX2-3* loci have been associated with multiple blood traits (platelets, red blood cells, monocytes^32^), as well as somatic mutation burden in prior work^13^ and this study (Supplementary Figure 9d). Knockout of *NRIP1* has recently been shown to reduce HSPC abundance^13^ while the transcription factor NKX2-3 is essential for HSC self-renewal^33^ and up-regulated in acute myeloid leukaemia (AML)^34^. While five of the six loci demonstrated a reduced level of association (*P* < 0.05) with driver CH or mCA, the *WDR70* locus was unique in showing no association with other subtypes of CH (Extended Data Figure 3a-f).

Given the strong association between CH-UD and female sex (Figure 3c), we performed sex-stratified GWAS to identify sex-specific genetic determinants of CH-UD. We identified 20 genome-wide significant loci in females and 14 in males, of which nine were shared between sexes (Extended Data Figures 4a and b). Of the 11 female-specific, nine were previously reported in driver CH or mCA GWAS, one was reported in our CH-UD GWAS (*NRIP1*), and one was novel to the female-only CH-UD GWAS (*SLC45A2*). Of the five loci specific to male-only CH-UD, two were previously reported for driver CH or mCA GWAS, one was reported in our CH-UD GWAS (*VSIG4/HEPH*), and two were novel to the male-only CH-UD GWAS (*L3MBTL3* and *NKX2-2/NKX2-4*). Both NKX2-2 and NKX2-4 are transcription factors involved in regulation of cellular differentiation, and *NKX2-4* is aberrantly activated in AML^35^. Lastly, nine overlapping loci were identified between female- and male-only CH-UD GWAS, and the effect sizes conferred by the leading SNPs of these loci demonstrated significant concordance (Pearson coefficient = 0.99, *P* = 3.34 × 10^−8^; Extended Data Figures 4c and d).

### Rare germline variants and CH-UD risk

To complement the common variant GWAS approach, we performed exome-wide association analysis (ExWAS) focusing on variants with a minor allele frequency < 1%. ExWAS identified two rare variants (rs41552812 and rs41542812) at the *HLA-DQB1* locus associated with CH-UD at exome-wide significance threshold (*P* < 1× 10^−8^; Supplementary Tables 13 and 14). The leading variant rs41552812, a missense variant on exon 2 of *HLA-DQB1* converting aspartic acid (negatively charged) to asparagine (hydrophilic but neutral), was associated with increased CH-UD risk (T allele: OR [95% CI] = 1.38 [1.26,1.51], *P* = 8.59 × 10^−12^). rs41552812 was no longer exome-wide significant (*P* = 0.030) after conditioning on rs41542812. This signal was independent of the earlier identified common variant GWAS loci.

Collapsing analysis, which aggregates rare germline variants within genes to increase statistical power^36,37^, identified significant associations with *CHEK2* and *DCLRE1B.* Both genes showed consistent associations across multiple qualifying variant (collapsing) models consisting of protein truncating, in-frame, and missense variants that collectively suggest loss-of-function mechanisms for both *CHEK2* (“flexdmg” collapsing model: OR [95% CI] = 1.022 [1.015,1.028], *P* = 6.86 × 10^−11^) and *DCLRE1B* (“ptvraredmg”: OR [95% CI] = 1.039 [1.027,1.051], *P* = 1.43 × 10^−10^; Extended Data Figures 5a-e; Supplementary Tables 15 and 16).

While *CHEK2* was also associated with increased risk of *DNMT3A*-CH and mLOY, *DCLRE1B* was exclusively associated with CH-UD at study-wide significance threshold (*P* < 1 × 10^−8^; Extended Data Figures 5f and g). *DCLRE1B* is a conserved gene involved in DNA damage repair and telomere maintenance^38,39^. Consistent with our observation that longer inherited LTL is associated with increased risk of CH-UD (Extended Data Figure 1c), rare loss-of-function germline variants in *DCLRE1B* are associated with longer LTL^23^.

### Plasma protein association analyses

To systematically investigate the relationship between CH-UD and the plasma proteome, we tested for associations between 2,941 plasma proteins and CH status in the 47,757 individuals for whom such data was available. In total, 13 plasma proteins were significantly elevated, and nine significantly depleted among individuals with CH-UD versus those without CH (*P* < 1.71 × 10^−5^, Bonferroni corrected) (Figure 5a and Supplementary Tables 17 and 18).

**Figure 5:**
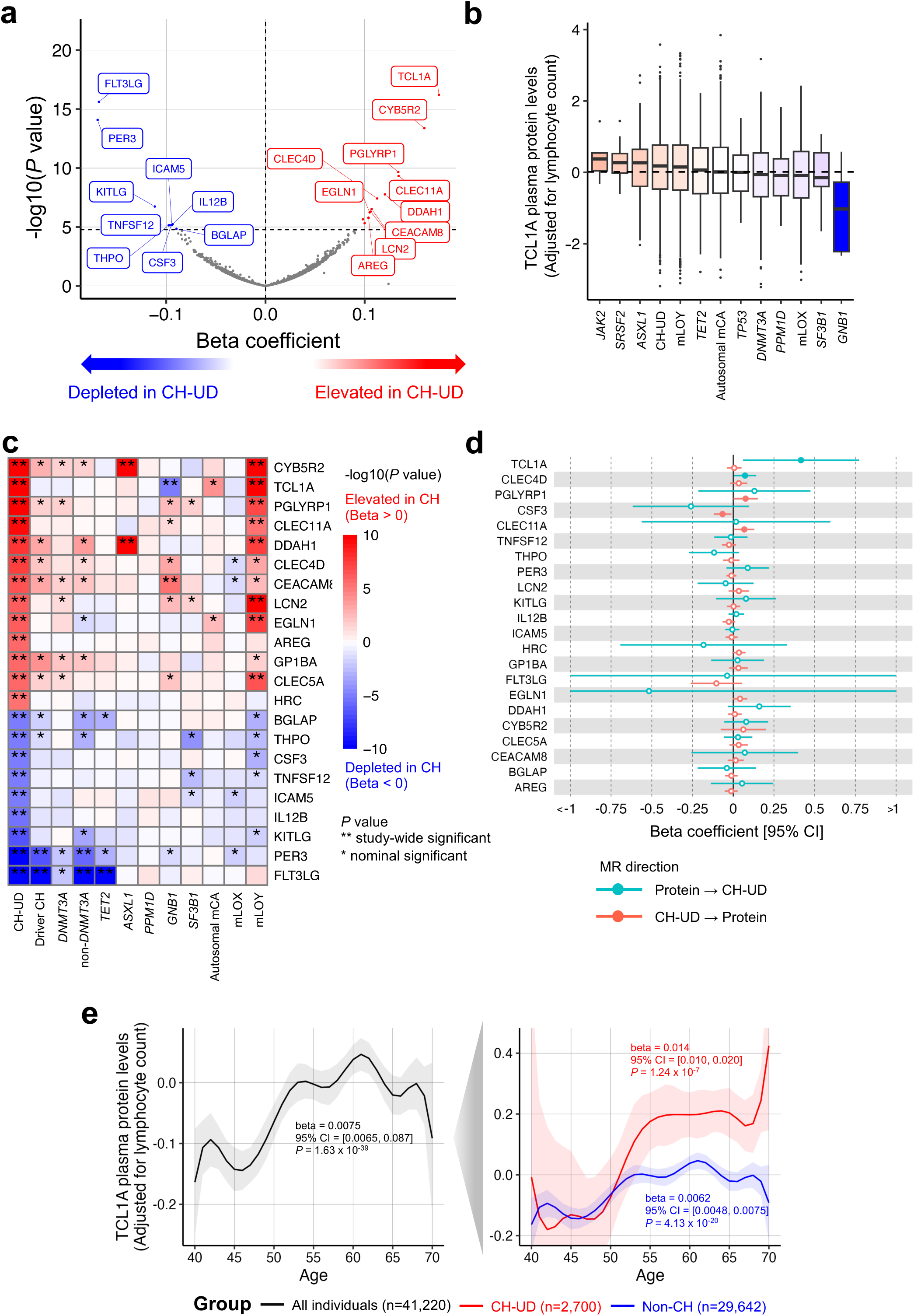
Plasma protein profile of CH-UD. **a,** Differential plasma protein level analysis in CH-UD versus non-CH individuals. Beta coefficients and *P* values were derived from linear regression with CH-UD status as the main predictor (non-CH as reference) and plasma protein level as the outcome adjusted for age, sex, smoking, waist circumstance, BMI, Olink batch, time between sample collection and plasma protein measurement, and first four *peddy*-inferred genetic principal components. Dashed line on the y-axis represents Bonferroni threshold (*P* = 1.71 × 10^−5^). **b,** Plasma protein level of TCL1A across the different classes of CH in descending order. Lymphocyte counts have been regressed out from the TCL1A protein levels here. Specifically, the TCL1A protein levels here were the residuals computed using linear regression model with protein level as the outcome and lymphocyte count as the predictor. Only CH subtypes with ≥5 individual with residual data available were included here. **c,** *P* values, with direction of coefficient indicated, for statistically significant plasma proteins in CH-UD versus non-CH individuals from **(a)** across CH subtypes. Only CH subtypes with ≥1 significant plasma protein at Bonferroni threshold and ≥10 carriers with plasma protein data available were included here. **d,** Bi-directional Mendelian randomisation with plasma protein level as the exposure and CH-UD as the outcome (teal blue), and conversely, CH-UD as the exposure and plasma protein level as the outcome (coral red). Only statistically significant plasma proteins in CH-UD versus non-CH individuals from **(a)** were included for analysis here. Beta coefficients and *P* values were derived from the median of inverse variance weighted, MR Egger, and weighted median. Solid circles represent significant associations (*P* < 0.05) while hollow circles represent non-significant associations (*P* ≥ 0.05). **e,** TCL1A protein levels by age for all individuals with plasma proteome data available (black), CH-UD individuals (red), and non-CH individuals (blue). The center line represents the fitted values from the general additive model with P-spline smooth class while the shaded regions represent the lower and upper bounds of the 95% CIs of the fitted values. Beta coefficient and *P* values were derived from linear regression with TCL1A protein level as the outcome and age as the main predictor adjusted for sex, smoking, and first four *peddy*-inferred PCs. NPX, normalised protein expression.

Notably, protein levels of TCL1A were the most significantly associated with CH-UD (beta [95 CI%]: 0.18 [0.13,0.22], *P* = 6.00 × 10^−17^), concordant with our common variant GWAS findings implicating the *TCL1A* locus. The strength of association of TCL1A protein levels with the different CH subtypes correlated with GWAS effect size of the G allele of the promoter variant (rs2887399; Figure 5b and Extended Data Figure 6a).

The most depleted plasma proteins in individuals with CH-UD were FLT3LG (beta [95 CI%]: −0.168 [−0.208,−0.128], *P* = 2.41 × 10^−16^) and PER3 (beta [95 CI%]: −0.169 [−0.212,−0.126], *P* = 8.28 × 10^−15^), which were also significantly depleted in driver CH (Figure 5c). Among the elevated proteins in CH-UD, CEACAM8 was also elevated in *GNB1*-CH, while CYB5R2 and DDAH1 were also elevated in *ASXL1*-CH and mLOY. Additionally, PGLYRP1, CLEC5A, EGLN1, LCN2, TCL1A, and CLEC11A plasma proteins were also elevated in mLOY. The remaining proteins were specifically elevated or depleted in CH-UD, namely CLEC4D, AREG, GP1BA, HRC, BGLAP, THPO, CSF3, TNFSF12, ICAM5, IL12B, and KITLG.

Unsupervised clustering using Euclidean distance revealed that CH-UD’s plasma protein profile more closely resembled mLOY, and not driver CH (Extended Data Figure 6b). However, most plasma proteins were significantly associated with only 1-3 of the 11 CH subtypes analysed (Extended Data Figure 6c). Although many of the significant changes in plasma protein levels were CH subtype-specific, gene set enrichment analysis revealed immune and inflammatory response pathways to be enriched across multiple CH subtypes (Supplementary Table 19), and particularly driver CH and autosomal mCA, consistent with a prior report^40^. In contrast, these pathways were not enriched in the plasma profile of CH-UD.

To investigate whether associations between plasma protein levels and CH subtypes were causal and, if so, the direction of effect, we conducted bi-directional Mendelian randomisation analyses. Genetic liability for higher TCL1A and CLEC4D plasma protein levels was associated with increased risk of CH-UD (TCL1A: beta [95 CI%]: 0.42 [0.06,0.77], *P* = 0.022; CLEC4D: beta [95 CI%]: 0.073 [0.004,0.14], *P* = 0.038; Figure 5d and Supplementary Table 20). Notably, genetic liability for higher TCL1A plasma protein levels was also associated with increased risk of *TET2-* and *ASXL1*-CH, (*TET2*-CH: beta [95 CI%]: 0.55 [0.16,0.93], *P* = 5.76 × 10^−3^; *ASXL1*-CH: beta [95 CI%]: 0.63 [0.18,1.08], *P* = 5.93 × 10^−3^) but decreased risk of *DNMT3A*-CH (beta [95 CI%]: −0.67 [−1.27,−0.063], *P* = 0.039) aligning with prior evidence for subtype-specific causal roles of TCL1A in driver CH^14^. Conversely, genetic liability for CH-UD was associated with increased plasma protein levels of CLEC11A (beta [95 CI%]: 0.068 [0.007,0.13], *P* = 0.029) and PGLYRP1 (beta [95 CI%]: 0.076 [0.001,0.151], *P* = 0.048), and decreased CSF3 levels (beta [95 CI%]: −0.066 [−0.12,−0.012], *P* = 0.017; Supplementary Table 21).

The presence of driver CH mutations, in genes like *TET2* and *ASXL1*, has been previously shown to increase chromatin accessibility at the *TCL1A* promoter which in turn leads to increased *TCL1A* expression and clonal expansion^14^. Given the prominent and causal association between TCL1A plasma protein levels and CH-UD, we hypothesised that TCL1A protein levels may drive clonal expansion even in the absence of driver CH mutations. Indeed, among CH-UD individuals, we observed TCL1A protein levels was associated with clonal expansion (PACER score; beta [95% CI] = 1.18 [0.059, 2.30], *P* = 0.039). We next investigated whether CH-UD risk factors were associated with TCL1A protein levels in both CH-UD and non-CH individuals. Elevated TCL1A protein levels were observed in individuals with at least one G allele for rs2887399 (Supplementary Figures 14a and b), elderly individuals (Figure 5e and Supplementary Figures 15a and b), females (Supplementary Figures 15c and d), and current smokers (Supplementary Figures 15e and f). Among these factors, the rs2887399 genotype had the largest influence on the inter-individual variability in TCL1A protein levels (Supplementary Figures 16a and b). Notably, the G allele of rs2887399 is associated with demethylation of the *TCL1A* promoter, which may in part explain this increased expression (Supplementary Figures 17a-c and Supplementary Notes 4). Finally, using a Bayesian hierarchical model, we demonstrated that clonal expansion in CH-UD individuals may be explained, in part, by stochastic *TCL1A* activation (Supplementary Figures 18a-j and Supplementary Notes 5).

### Phenome-wide association analyses

To explore phenotypic associations with CH-UD, we performed phenome-wide association analyses in 404,643 UKB participants, spanning 13,225 binary traits/diseases and 1,682 quantitative measurements. CH-UD demonstrated significant associations with 139 binary phenotypes (*P* < 3.78 × 10^−6^) and 24 quantitative phenotypes (*P* < 2.97 × 10^−5^).

Across phenotypes, CH-UD exhibited a broad disease-association pattern similar to other forms of CH, with the strongest associations observed for neoplasms (Figure 6a and Extended Data Figure 7a). Specifically, of the 139 phenotypes significantly associated with CH-UD, 58 (42%) were linked to neoplasm (Supplementary Tables 22 and 23). CH-UD demonstrated particularly strong associations with haematological malignancies, including both myeloid leukaemia (ICD-10 = C92, OR [95% CI] = 3.08 [2.30,4.13], *P* = 1.22 × 10^−7^) and lymphoid leukaemia (ICD-10 = C91, OR [95% CI]: 2.84 [2.20,3.67], *P* = 1.98 × 10^−8^). The strongest association was with essential thrombocythaemia (ET; ICD-10 = D473, OR [95% CI] = 5.20 [4.03,6.07], *P* = 5.79 × 10^−19^), a subtype of chronic myeloproliferative neoplasm (MPN). Two-sample Mendelian randomisation analyses for binary phenotypes using genetic association results from the FinnGen study^41^ provided support for the association between genetic liability for CH-UD and increased risk of ET (OR [95% CI] = 1.76 [1.32,2.36], *P* = 1.41 × 10^−4^; Supplementary Table 24).To address the possibility that this purported relationship was due to undetected, low-level driver CH mutations associated with MPN drivers, we applied a more sensitive (but less precise) approach to exclude individuals with *JAK2* V617F mutations, and additionally excluded individuals with *CALR* mutations in the sequencing data (Supplementary Figure 19 and Supplementary Notes 6). The association between CH-UD and ET remained highly significant after excluding these individuals from analysis (OR [95% CI]: 4.31 [3.24,5.73], *P* = 3.15 × 10^−12^; Supplementary Figure 20).

**Figure 6:**
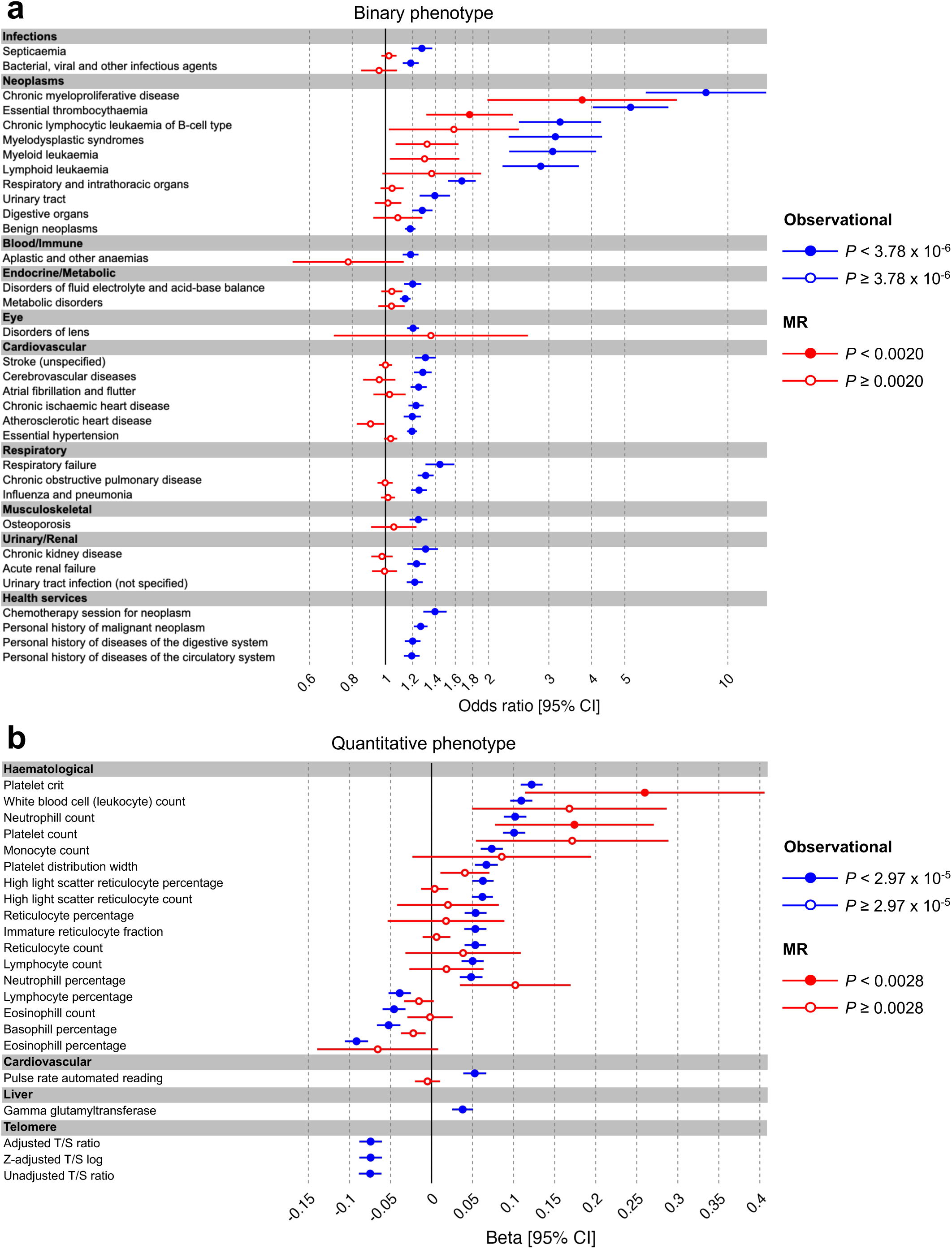
Association between CH-UD versus selected binary and quantitative phenotypes. a, Association between CH-UD versus selected binary phenotypes. For association analyses in UKB (observational; blue), odds ratios and *P* values were derived from Firth logistic regression with trait or disease as the outcome and CH-UD as the main predictor (non-CH as reference) adjusted for age, sex, smoking, waist circumferences, body mass index, and first four *peddy*-inferred genetic principal components. Phenotypes were ordered by ICD10 chapters and all associations shown here were significant at Bonferroni threshold (*P* < 3.78 x 10^−6^). Complete list of significant phenotypes provided in Supplementary Table 23. For two-sample MR (red), CH-UD was defined as the exposure and phenotypes from FinnGen were defined as the outcomes. Odds ratios and *P* values were derived from the median of inverse variance weighted, MR Egger, and weighted median approaches. Significant associations at Bonferroni threshold (*P* < 0.0020) indicated by the solid circles. **b,** Association between CH-UD versus selected quantitative phenotypes. For association analyses in UKB (observational; blue), beta coefficients and *P* values were derived from linear regression with quantitative measurement as the outcome and CH-UD was the main predictor (non-CH as reference) adjusted for age, sex, smoking, waist circumferences, body mass index, and first four *peddy*-inferred genetic principal components. All phenotypes presented here belonged to “Chapter XVIII Symptoms signs and abnormal clinical and laboratory findings not elsewhere classified” and all associations shown here were significant at Bonferroni threshold (*P* < 2.97 x 10^−5^). Complete list of significant phenotypes provided in Supplementary Table 26. For one-sample MR (red), CH-UD was defined as the exposure and phenotypes from UKB (Neale’s lab) were defined as the outcomes. Beta coefficients and *P* values were derived from the median of inverse variance weighted, MR Egger, and weighted median approaches. Significant associations at Bonferroni threshold (*P* < 0.0028) indicated by the solid circles. MR, Mendelian randomisation.

CH-UD also demonstrated association with myelodysplastic syndrome (MDS; ICD-10: D46, OR [95% CI] = 3.13 [2.29,4.29], *P* = 5.49 × 10^−7^), chronic lymphocytic leukaemia (ICD-10: C911, OR [95% CI] = 3.24 [2.45,4.27], *P* = 5.04 × 10^−9^), and several solid tumours, including neoplasms of the respiratory and intrathoracic organs (ICD-10: C30-C39, OR [95% CI]: 1.67 [1.52,1.83], *P* = 6.34 × 10^−16^), urinary tract (ICD-10: C64-C68, OR [95% CI]: 1.39 [1.26,1.54], *P* = 1.94 × 10^−6^), and digestive organs (ICD-10: C15-C26, OR [95% CI]: 1.28 [1.20,1.37], *P* = 8.30 × 10^−8^). Additional associations, as with other types of CH, included aging-related phenotypes affecting the eye, cardiovascular, respiratory, musculoskeletal and renal systems. Using Cox proportional hazards regression, CH-UD was associated with 20% higher all-cause mortality (HR [95% CI] = 1.20 [1.15,1.24], *P* = 7.17 × 10^−21^), comparable to driver CH (HR [95% CI] = 1.26 [1.21,1.32], *P* = 1.48 × 10^−23^; Extended Data Figure 8 and Supplementary Table 25).

Among quantitative phenotypes, 20 of 24 (83%) significant associations (*P* < 2.97 × 10^−5^) with CH-UD involved LTL and blood count parameters (Figure 6b, Extended Data Fig 9a, and Supplementary Table 26). CH-UD was associated with shorter LTL, likely due to progressive telomere attrition during clonal HSC division^23^ (Extended Data Figure 10). The most prominent blood count-related association was increased platelet crit (beta [95% CI] = 0.122 [0.109,0.135], *P* = 1.25 × 10^−51^), a feature related, but not exclusive, to ET. The association between CH-UD and platelet crit remained significant after removing *CALR* indels, and more stringent removal of *JAK2* V617F mutation carriers, from the CH-UD group (OR [95% CI] = 0.11 [0.10,0.13], *P* = 3.71 × 10^−29^; Supplementary Figures 21 and Supplementary Notes 7).

Additional significant blood count-related phenotypes included elevated neutrophil count (beta [95% CI] = 0.102 [0.088,0.116], *P* = 1.06 × 10^−34^), monocyte count (beta [95% CI] = 0.073 [0.060,0.087], *P* = 3.89 × 10^−19^), reticulocyte percentage (beta [95% CI] = 0.054 [0.040,0.067], *P* = 4.83 × 10^−11^), and reduced eosinophil count (beta [95% CI] = −0.046 [−0.060,−0.032], *P* = 9.43 × 10^−8^) and basophil percentage (beta [95% CI] = −0.052 [−0.067,−0.038], *P* = 2.30 × 10^−9^). Mendelian randomisation analysis in UKB supported causal effects on blood parameters including elevated platelet crit (beta [95% CI] = 0.260 [0.114,0.406], *P* = 1.32 × 10^−3^) and neutrophil count (beta [95% CI] = 0.174 [0.077,0.271], *P* = 1.20 × 10^−3^; Supplementary Table 27).

Comparative analysis revealed that CH-UD shared many associations with other CH subtypes while maintaining its own distinct pattern. Among binary phenotypes, 20 of 33 CH-UD-associated traits shown in Figure 6a were also significantly associated with driver CH, 8 with autosomal mCA, 3 with mLOX, and 20 with mLOY (Extended Data Figure 7b). For quantitative phenotypes, 14 of 22 CH-UD-associated traits shown in Figure 6b were shared with driver CH, 3 with autosomal mCA, 5 with mLOX, and 18 with mLOY (Extended Data Figure 9b).

## Discussion

We present the largest population-scale investigation of CH-UD to date, enabled through the development and application of a new ML framework to determine the CH-UD status of 407,512 UKB participants with blood WGS data. Our approach enables improved CH-UD case ascertainment by integrating singleton detection with mutation type and context, chromatin accessibility and mutational tolerance. We identified 26,963 individuals with CH-UD who, when added to those with driver CH and mCAs, reveal that more than 1 in 4 UKB participants had a form of CH of sufficient clonal size to be detectable by WGS, WES or SNP array. Using the extensive genomic, proteomic and phenotypic resources of the UKB, we went on to conduct a comprehensive genetic and phenotypic characterisation of CH-UD. By comparing this to analogous analyses of CH and mCA, we revealed the shared and distinct causes and consequences of these forms of CH.

The first study to capture CH-UD at population scale relied on WES data, thus underestimating the true prevalence of this class of CH^21^. The detection of CH-UD in our study employed WGS data, which increased the number of detectable passenger variants^42^. Three recent studies also used WGS to identify CH, including cases without evidence of driver CH variants^11–13^. The first two studies aggregated individuals with detectable CH clones as a single CH class, irrespective of the presence or absence of driver CH or mCA^11,12^. While this approach yielded novel insights into CH as a whole, it could not capture heterogeneity between different CH classes. It is clear that different subtypes of CH can have different – and sometimes even opposite – germline associations, as observed for different types of driver gene-specific and long versus short inherited LTL^5^, or major versus minor alleles of a *TCL1A* promoter variant^2–4^. The third study modelled passenger variants in non-driver CH individuals as a continuous trait and analysed a much smaller number of samples^13^. In contrast, in our study, we used an enhanced ML-based approach for CH-UD identification, applied this to the largest WGS cohort analysed for this purpose to date, to investigate the causes and consequences of CH-UD as a distinct entity.

Comparative analysis of CH-UD, driver CH, and mCA revealed similarities and differences with respect to their population frequencies, genetic determinants and phenotypic associations/consequences. First, we found that CH-UD was more prevalent than driver CH, autosomal mCA or mLOX, making it the second most common form of CH after mLOY. Second, CH-UD was more common in females, in whom the effect of smoking on CH-UD prevalence was greater compared to males. Third, 30 of 31 loci associated with CH-UD at GWAS (n=25) or nominal (n=5) significance were previously linked to driver CH and/or mCA. Fourth, we characterised the plasma proteome of CH-UD, and through Mendelian Randomisation discovered that elevated levels of both TCL1A and CLEC4D are causally associated with increased CH-UD risk. Finally, we found CH-UD to be associated with 163 binary and quantitative traits, with causal relationships demonstrated for myeloid neoplasms and a range of blood parameters, as well as a 20% increase in all-cause mortality. Notably, as CH-UD and driver CH share genetic determinants, we used MR-Egger to mitigate pleiotropy. Nevertheless, further work is required to confirm that the relationships between CH-UD and these phenotypes are indeed causal.

It is conceivable that a fraction of CH-UD may harbour known somatic driver variants located outside the current consensus list of CH drivers^43^. Indeed, of 26,963 individuals with CH-UD, we identified 25 (0.093%) cases with *CALR* mutation using focused somatic variant calling with Mutect2, and 274 (1.02%) cases with 1-2 sequencing reads reporting the *JAK2* V617F mutation using *in silico* genotyping (pileup). It is noteworthy that *CALR* is currently not on the consensus list of CH driver genes, and *in silico* pileup genotyping of mutational hotspots is currently not within recommended workflows for CH detection^43^. Furthermore, we recently identified ∼150 individuals with somatic TERT promoter mutations^5^, whilst another study analysing UKB WGS data identified additional carriers of mCAs previously missed by studies using SNP arrays^44^, Therefore, it is likely that some individuals within our CH-UD group carry hitherto undetected genetic driver variants, and this represents an opportunity to discover or validate novel CH driver genes^15,45^. However, the frequency of CH-UD was approximately 1.7-fold higher to that of driver CH. This is very unlikely to be explained by undiscovered or missed driver CH variants, particularly as our approach for CH-UD detection is biased towards large clones in which driver mutations are easier to detect.

Our observation that 30 of 31 CH-UD GWAS loci were previously identified in other forms of CH suggests shared mechanisms of clonal expansion. To examine which form of CH is most similar to CH-UD, we used LDSC analysis to examine the genetic correlation between CH subtypes. This revealed a very high genetic correlation (r_g_= 0.794) between CH-UD and non-*DNMT3A*-CH, suggesting that the two forms of CH share global mechanisms of clonal expansion. Other forms of CH showed much lower r_g_ with CH-UD or between them (Figure 4a). This high degree of similarity should not come as a surprise as ∼¾ of non-DNMT3A CH is driven by mutations in *TET2* and *ASXL1*, two important epigenetic genes that directly or indirectly influence DNA methylation^46^. To put the degree of genetic corelation in perspective, it is notable that cancers with known shared heritability, like breast and ovarian only have an LDSC-derived rg of ∼0.2^47^.

Acquisition of such CH-associated mutations in human HSCs only leads to detectable CH in the minority of cases, with most resulting in multiple tiny CH clones in older humans^17^. This proposes that the epigenetic changes responsible for the increased clonal fitness of *TET2*-CH and *ASXL1*-CH are not inevitable, but rely on selectable semi-stochastic epigenetic changes. In the absence of such somatic genetic drivers, the epigenetic changes associated with HSC ageing may operate to drive the clonal selection associated with the development CH-UD. In fact, there are substantial similarities in the epigenetic changes observed with *TET2* loss-of-function (LoF) mutations and HSC ageing^48,49^.

Beyond their global genetic similarity, a key connection between CH-UD and non-*DNMT3A* CH is the strong association of both with polymorphisms in the *TCL1A* gene, particularly the G-allele of the rs2887399 SNP located within its promoter. In fact, *TCL1A* was the most significant GWAS hit for CH-UD and *ASXL1*-CH and second only to *TERT* for *TET2*-CH. *TCL1A* is not normally expressed in human HSPCs, but the introduction of *TET2* and *ASXL1* truncating mutations into these cells led to increased chromatin accessibility at the *TCL1A* locus and associated TCL1A protein expression in a proportion of HSPCs^14^. Mirroring this, we show here that TCL1A was the most significantly elevated protein in the plasma of people with CH-UD and provide evidence that this association is causal using MR. This supports the premise that clonal expansion in CH-UD is epigenetically driven, with *TCL1A* activation playing a central role. Our finding that TCL1A plasma levels rise with advancing age even in those without CH, proposes that ageing-related epigenetic changes can activate *TCL1A* expression without the need for somatic genetic driver variants. In support of this we show that the G allele of rs2887399 is associated with demethylation of the *TCL1A* promoter in the absence of CH mutations. In turn, the presence of this, and possibly other, epigenetic driver variants provides an explanation for the ubiquitous presence of CH-UD in virtually everyone over the age of 70 years (Supplementary Figure 22). The recent demonstration that TCL1A forms a multimeric complex with and inhibits the function of DNMT3A^50,51^, gives this hypothesis further credence.

Mirroring the above similarities, we demonstrated that, like driver CH and mCA, CH-UD was associated with diverse adverse outcomes including haematological malignancies and overall survival. This suggests that CH-UD warrants equivalent consideration to driver CH and mCA in clinical practice. Current approaches for the detection of CH-UD clones or “barcode” CH^12^, including our own, highlight the challenges in detecting CH-UD. Specifically, we and others rely on finding singletons representing passenger variants to identify CH-UD clones. Singletons, in turn, can only be confidently ascertained using large sample sizes such as population-scale biobanks. However, novel approaches to identify the presence of clonal expansion such as the use of methylation barcodes^52^, may enable scalable CH-UD detection.

In summary, our work represents the largest and most comprehensive investigation of CH-UD to date, revealing novel insights into its determinants, phenotypic consequences, and connections with ageing and age-associated diseases. Furthermore, while CH-UD displays unique characteristics, cementing its status as a distinct entity, substantial overlaps with other CH subtypes demonstrate important shared clinical consequences and suggest convergence on shared epigenetic pathways responsible for their clonal expansion.

## Methods

### Study population

Participants were sourced from the U.K. Biobank (UKB). The UKB study has approval from the North-West Multi-centre Research Ethics Committee (11/NW/0382).

### Sample selection

Samples were selected as described in our previous works^4^. Specifically, samples were selected on the basis of (1) WES data available, (2) contamination <4% as inferred by *verifyBamID* software^53^, (3) sex concordant between clinically reported and chromosome X:Y consensus coding sequence (CCDS) coverage ratios, (4) ≥94.15% of CCDS r22 bases^54^ covered with ≥10x coverage, (5) *peddy*-inferred^55^ probability ≥0.95 of European ancestry (6) no pairs with kinship >0.1769 as inferred with *KING* software^56^, (7) within 4 standard deviations (SDs) of mean genetic PCs 1-4 as computed by the *peddy* software v0.3.2^55^, (8) not included as part of the panel of normals (PoN) for driver CH detection, (9) consent not withdrawn as of April 2024, and (10) no diagnosis of haematological malignancies prior to blood sample collection. In this study, samples were further selected on the basis of (1) WGS data available, (2) mCA data available^22^, and (3) not “hypermutants” as defined as samples <95 percentile for the number of singletons detected. In total, 407,512 samples were included in this study, of which 205,736, 158,426, and 43,350 samples were part of the deCODE, Wellcome Sanger Institute (WSI) main phase (WSI main), and WSI Vanguard phase (WSI Vanguard), respectively. The WSI Vanguard cohort included 4,503 samples as part of the Vanguard pilot phase (Supplementary Table 1).

### Driver CH detection

Putative driver somatic variants were identified in 15 pre-leukaemic driver genes, as described in our previous work^4^, namely *DNMT3A*, *TET2*, *ASXL1*, *PPM1D*, *TP53*, *SF3B1*, *SRSF2*, *GNB1*, *IDH2*, *JAK2*, *PRPF8*, *KRAS*, *NRAS*, *BRAF*, and *MPL*. Specifically, variants were retrieved using Mutect2, and annotated with transcript ID, exon number, cDNA change, amino acid change, and protein consequence with Ensembl Variant Effect Predictor software^57,58^. Variants were retained if they were (1) supported by ≥3 alternate allele reads, (2) had VAF of 3-40%, (3) demonstrated increased frequency with age, and (4) conformed to previously defined criteria as set out by the Trans-Omics for Precision Medicine (TOPMed) Program^1,43^ (Supplementary Table 2).

### Singleton detection

Germline variants were first identified from WGS data using Illumina DRAGEN software, and variants passing quality control (QC) defined as Fischer stand bias (FS) ≤60 for SNVs, FS ≤200 for indels, mapping qualify (MQ) ≥30, genotype quality (GQ) ≥20, variant quality score (QUAL) ≥20 or log of odds (LOD) ≥4 were retained and stored in our in-house variant platform GNEX. Singletons were then identified and retrieved from GNEX. A singleton is defined as a genetic variant identified only once in the cohort. In total, 422,295,944 singletons were identified among UKB participants. We further filtered singleton variants for those (1) with VAF ≤25%, (2) with sequencing depth 15-60, (3) supported by ≥3 alternate allele reads, (4) not reported in Genome Aggregation Database (gnomAD) database, (5) with genotype missing rate ≤10% and (6) located on autosomal chromosomes (Extended Data Figure 1). As the ability to confidently classify a genetic variant as singleton, and by extension, a putative passenger variant, depends on the sample size of the ancestry group, we only included the largest ancestry group available in the UKB, namely Europeans (Extended Data Figure 2).

### CH with unidentified driver detection

#### Feature selection

Previous studies inferred pre-leukaemic clones based on the number of singletons or mutation burden^11,12,15,21^ while a recent study have demonstrated that somatic single-nucleotide variant (SSNV) counts alone may not adequately distinguish CH samples, with or without driver mutations, from neutrally evolving samples^18^.

To this end, we employed a multi-parametric approach to detect pre-leukaemic clones^13^. Specifically, we defined several genetic features to infer pre-leukaemic clones in addition to the (1) number of singletons, namely (2) count or fraction of singletons by variant type (SNV, deletion, insertion), (3) count or fraction of nucleotide change in single-nucleotide context (C>A, C>G, C>T, T>A, T>C, T>G), (4) count of nucleotide change in trinucleotide context (e.g., TCA>TAA, TCA>TGA, TCA>TAA), (5) count or fraction of singletons located in the open chromatin region of haematopoietic stem cells (HSCs)^13,59^, and (6) genomic position in terms of tolerance to mutations (genome-wide residual variation intolerance score (gwRVIS)) score^60^. For each sample, the count or fraction of singletons with gwRVIS scores >0 and mean gwRVIS score were computed. In total, 120 features were identified, specifically 1, 6, 12, 96, 2, and 3 features from (1), (2), (3), (4), (5), and (6) category, respectively.

To identify redundant features, we performed correlation test for all possible pairs of features. Correlated feature pairs were defined with Pearson coefficient >0.9^61,62^. This revealed five pairs of correlated features namely, (1) number of singletons vs. SNV count, (2) number of singletons vs. number of singletons with gwRVIS score >0, (3) SNV count vs. number of singletons with gwRVIS score >0, (4) C>A count vs. CCA>CAA count, and (5) C>A count vs. CCT>CAT count. We selected number of singletons and C>A count as non-redundant features from this list of feature pairs. This results in 116 non-redundant features for downstream analysis.

#### Training and testing set selection

We applied a ML approach that incorporated the non-redundant features to identify CH-UD. As the different WGS batches demonstrated variable sequencing metrics, such as sequencing depth, we proceeded to train and test our ML classifiers on each batch separately.

First, we identified our positive control sample set as individuals with driver CH^4^ or mCA^22^ (n=39,571 from deCODE, n=30,112 from WSI main, and n=8,202 from WSI Vanguard). For the latter, we further retained individuals with cell fraction available. Second, we identified our negative control samples set as individuals ≤44 years old and without any detectable driver CH or mCA (n=18,883 from deCODE, n=14,140 from WSI main and n=3,776 WSI Vanguard). The frequency of driver CH or mCA among individuals ≤44 years old were 4.96%, 4.97%, and 5.14% in deCODE, WSI main, and WSI Vanguard, respectively.

For each WGS batch, we split our individuals into 60% training and 40% test set. For our training set, we split our positive controls into samples with driver CH only, samples with mCA only, and samples with any driver CH or mCA. We further subset 50% of our training set for the purpose of fine tuning the hyperparameters of our ML classifiers. For our test set, we performed down-sampling to obtain a 1:10 ratio of positive and negative controls to recapitulate the approximate rate of CH reported in previous epidemiological studies^1,2,4^.

#### Machine learning classifier selection

We first used random forest to assess which training set (driver CH, mCA, or any CH) yielded the best performance in the test set (any CH). Measurements of performance were accuracy, area under curve (AUC), F1 score, sensitivity, negative predictive value (NPV), and precision. We observed the classifier trained on driver CH outperformed the classifiers trained on mCA or any CH on four out of the six performance metrics, namely accuracy, AUC, F1, and precision. Therefore, we subsequently trained and assess additional ML classifiers using driver CH as the training set.

In total, we trained and assessed four ML classifiers, namely, XGBoost, random forest, SVM, and logistic regression. We first tuned the hyperparameters for XGBoost and SVM. Specifically, we tested a range of values for the learning rate and maximum tree depth parameters for XGBoost and for the constraints violation parameter for SVM. XGBoost, random forest, SVM, and logistic regression were implemented with the *xgboost*, *randomForest*, *e1071*, and *stats* R package, respectively, with the default parameters, with the exception of XGBoost where *eta* was set to 0.1 and *max_depth* to 3 for all WGS batches and SVM where *cost* was set to 1 for deCODE and WSI main and 5 for WSI Vanguard.

We additionally assessed an approach based solely on the number of singletons. For this singleton-only approach, we first defined a range of number of singletons (1-100) for assessment in the training set. The best threshold was based on the number of singletons that yielded the highest accuracy in the training set (≥19 deCODE; ≥32 WSI main; ≥12 WSI Vanguard). These thresholds were then assessed in the test set and the performance metrics from the test set were reported and compared against the performance of the ML classifiers.

All four ML classifiers and the singleton-only approach were applied on individuals without detectable driver CH or mCA and with at least one singleton detected to infer the presence or absence of CH-UD (n=165,021 from deCODE, n=127,739 from WSI main, and n=33,885 from WSI Vanguard). While all ML classifiers demonstrated comparable performance, each classifier identified a subset of individuals with CH-UD which were not identified by the other classifiers. We therefore defined a set of heuristic criteria to determine the best classifier, namely (1) association between detected CH-UD with age (2) expected low frequency of detected CH-UD at 40 years old, and (3) agreement in age-dependency between sequencing centres. To this end, individuals with CH-UD inferred using the random forest approach was brought forward for downstream analysis.

It is noteworthy that there were individuals without detectable driver CH or mCA or detectable singletons (n=1,144 from deCODE, n=575 from WSI main, and n=1,263 from WSI Vanguard). These individuals are presumed to have no CH-UD and were treated as controls (non-CH) in our downstream analysis.

### Combining CH calls

For all association analyses, we created exclusive CH phenotypes^3^. Specifically, only mono-mutant CH individuals were included for association analyses, while multi-mutant CH individuals were excluded from association analyses. Mono-mutant CH individuals were defined as individuals with either driver CH only, mCA only or CH-UD. Among individuals with driver CH, only individuals with mutation(s) in one driver CH gene were included for association analyses. Among individuals with mCA, only females with either autosomal mCA or mLOX and only males with either autosomal mCA or mLOY were included for association analyses. In total, 15,848 CH driver, 10,346 autosomal mCA, 10,798 mLOX, 36,898 mLOY, 26,963 CH-UD, and 302,664 non-CH individuals were included for association analyses.

### Singleton VAF adjustment

Restricting genetic variants to singletons enriches for passenger somatic variants. However, it is conceivable that while singletons detected are enriched for variants of somatic origin, they may contain some degree of private, *de novo*, or ultra rare germline variants^13^.

To this end, we made adjustments to the allele frequency of each singleton to reflect its probability of somatic and germline origin. First, we fitted a beta distribution to VAF for a repertoire of putative somatic variants, specifically the pre-leukemic driver variants identified from individuals with driver CH. Next, we fitted a beta distribution to VAF for a repertoire of putative germline variants, specifically the singletons identified from individuals ≤44 years old and without detectable CH (driver CH, mCA, or CH-UD). The beta distributions were fitted using the maximum likelihood estimation implemented by *fitdist* function from the *fitdistrplus* R package.

Subsequently, we computed the likelihood that a given singleton is of somatic or germline origin. First, we retrieved the hyperparameters (α and β) from the beta distribution fitted with the putative somatic variants above to determine the likelihood a given singleton VAF is under the somatic model (likelihood_somatic_). Similarly, we retrieved the hyperparameters from the beta distribution fitted with the putative germline variants above to determine the likelihood a given singleton VAF is under the germline model (likelihood_germline_). These likelihoods were computed using the *dbeta* function from the *stats* R package. We then defined a heuristic prior somatic probability (prior_somatic_) of 0.75 and, conversely, a heuristic prior germline probability (prior_germline_) of 0.25.

We next implemented Bayes’ rule to compute the posterior probability a given singleton is somatic of origin:

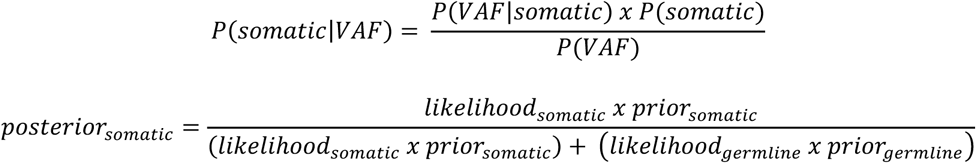

The posterior somatic probability is then used as weights to adjust the raw VAF. Specifically, the raw VAF of each singleton is multiplied by its respective posterior somatic probability. We used the median of the weighted VAF across the singletons as the representative weighted VAF for each sample.

### Singleton and singleton-derived VAF validation

To demonstrate enrichment of genetic variants of somatic origin among our singleton call set, we evaluated the association between singleton burden and VAF against age at blood sample collection.

Singletons were further assessed for the enrichment or depletion of specific mutational signatures previously reported in myeloid neoplasms^63^, namely SBS1, SBS5, SBS8, SBS18, SBS19, SBS2, SBS13, SBS45, and SBS46. We additionally assessed the enrichment of HSC-related somatic mutational signature^64,65^ by appending this signature to the original list of signatures during the second iteration of mutational signature extraction. Mutational signatures were computed using non-negative matrix factorization (NMF) as implemented in SigProfilerAssignment^66^.

Singleton-derived VAF were further assessed by performing association analysis between measured LTL and risk of driver CH or mCA^23^. Only individuals with small clones, defined as individuals with <10^th^ quantile of singleton-derived VAF, were included in this analysis. This is based on the premise that the measured LTL in individuals with small clones are more representative of LTL from non-mutant cells, and by extension, inherited or background LTL.

Finally, both singletons and singleton-derived VAF were further assessed by inferring clone fitness and GWAS of singleton burden. As previously described^14^, we fitted a negative binomial generalised linear model with number of singletons as the outcome and CH as the predictor (non-*DNMT3A* R882 individuals as reference) with adjusted singleton-derived VAF and WGS batch as co-variates. The negative binomial generalised linear model was implemented using the *glm.nb* function from the *MASS* R package. The adjusted singleton-derived VAF were the residuals after regressing out the following variables from the singleton-derived VAF in a linear regression model: sequencing depth at singleton site, age, sex, smoking, and first four *peddy*-computed genetic PCs.

For GWAS of singleton burden, see “Genome-wide association analyses” below.

### Proteomics-based biological age quantification

We assessed biological age as a potential risk factor for CH. Biological age for UKB participants were quantified using plasma proteins as previously described^24^. In total, 2,941 Olink measurements for 47,757 individuals were available. Six proteins (CXCL8, IDO1, IL6, LMOD1, SCRIB, and TNF) corresponded to multiple Olink assay measurements, and for each of these proteins, we derived a single measure by calculating the mean across those assay measurements. This resulted in 2,923 unique plasma proteins. We further excluded 324 individuals with >49.9% of proteins with missing values and six proteins with missing values in >10% of samples. For each of the remaining plasma protein, we imputed the missing values by calculating the median across all individuals. The values of each plasma protein were then scaled and centred. Fifteen individuals without age information were removed. In total, 47,418 individuals and 2,917 plasma proteins were included for downstream analysis.

We computed the biological age using net elastic regression. We first fine-tuned the alpha hyperparameter by testing a range of values for this hyperparameter using a k-fold cross-validation approach, where k=5. Samples were split into five groups (folds) of equal size. The first fold (k=1) is defined as the training set and the remaining folds collectively formed the testing set. For a given value of the hyperparameter, we trained the model on chronological age using the *cv.glmnet* function from the *glmnet* R package. The model was then applied on the testing set to predict each sample’s age using the *predict* function from the *stats* R package. The mean absolute error (MAE) between the chronological ages and predicted ages was then calculated. This is repeated for a range of values for the hyperparameter, and then for k=2,3,4, and 5 as the training set. The mean MAE for each value of the hyperparameter was computed, and the value of the hyperparameter that yielded the smallest MAE is selected (0.05).

This alpha value was then used to train the model on chronological age from 9,484 individuals in the training set (first fold) and then used to predict the biological age among 37,934 individuals in the testing set (remaining folds). The age gap among individuals in the testing set were also computed as the residuals from the linear model with biological age as the outcome and chronological age as the predictor. Only individuals from the testing set, and their corresponding biological age and age gap, were included for downstream analysis. The intercept and list of plasma proteins with non-zero coefficients are provided in Supplementary Table 4.

### Genome-wide association analysis

Sample- and variant-level QC were performed as described in our previous study^4^. Specifically, sample-level QC was performed on the basis of (1) retaining individuals of European descent, removing (2) samples with genotype missingness >5%, (3) samples with non-XX or -XY chromosome configuration, and (4) samples with high heterozygosity. Variant-level QC was perform on the basis of removing (1) variants with missingness ≥2%, (2) non-autosomal variants, (3) variants with C>G, G>C, A>T, or T>A base change, (4) variants in long-range LD regions or in commonly inverted regions, insertions or deletions, and (5) variants with different allele frequencies between UKBB and UKBL (defined with *P* < 10^−12^ when Fisher exact test applied on genotype counts), and retaining (6) variants with MAF ≥1%, (7) variants with HWE *P* < 10^−6^, and (8) variants pruned for LD r2 <0.1 (windows of 50 SNPs and a size step of 5 SNPs). The UKBB and UKBL represent two closely-related purpose-designed Affymetrix array, namely UK BiLEVE Axiom array and UK Biobank Axiom array, respectively. The remaining samples and variants were used in step 1 of SAIGE to compute the genetic relationship matrix that was subsequently used to adjust for population stratification in step 2 of SAIGE^67^. Variants imputed with Haplotype Reference Consortium (HRC) panel, and additionally with UK10K + 1000 Genomes panel for variants not present in HRC^68^, were used in step 2 of SAIGE. In step 2 of SAIGE, genetic association analysis was performed using Firth logistic regression adjusted for age, sex, smoking, *peddy*-inferred European probability, and first ten genetic PCs. Both step 1 and step 2 were performed for each WGS batch separately. The variant coordinates were additionally converted from GRCh37 to GRCh38 human reference genome assembly with CrossMap^69^. Each variant was then annotated relative to its nearest genes and the distance relative to each gene using GENCODE v48 as the reference^70^.

GWAS of singleton burden was performed using linear regression implemented by SAIGE^67^ as previously described^13^. Specifically, the outcome was singleton burden among individuals with CH (driver CH, mCA, and CH-UD) and the co-variates were age, sex, smoking, *peddy*-inferred fraction of European probability, first ten genetic PCs, and adjusted singleton-derived VAF. The adjusted singleton-derived VAF were the residuals after regressing sequencing depth at singleton site from the singleton-derived VAF in a linear regression model. Genome-wide significant variants were defined with MAF ≥1%, imputation score >0.6, and *P* < 5 × 10^−8^.

GWAS of driver CH, mCA, and CH-UD, was performed using Firth logistic regression implemented by SAIGE^67^. For gene-specific driver CH, only CH driver genes with at least 10 samples across all three WGS batches included for analysis, namely *DNMT3A*, *TET2*, *ASXL1*, *TP53*, *PPM1D*, *GNB1*, *JAK2*, *SRSF2*, and *SRSF2+SF3B1*. We ran the GWAS on each WGS batch separately, and included age, sex, smoking, *peddy*-inferred European probability, and first ten genetic PCs as co-variates.

To annotate genome-wide significant loci for CH-UD with that previously reported CH GWAS, we collated a list of previously reported genome-wide significant loci for driver CH GWAS (overall CH or gene-specific driver CH)^2–4,71^ and mCA (autosomal mCA, mLOX, mLOY) GWAS^3,22,72–76^. We further appended this list with genome-wide significant loci identified from driver CH GWAS and mCA GWAS from this study. For a given CH-UD locus, we considered this locus to have been previously reported if any given SNPs within this locus overlapped within 500kb, in both 5’ and 3’ directions, with any given SNPs of the loci identified from driver CH GWAS and mCA GWAS.

On top of the leading SNPs of genome-wide significant loci, we further identified additional independent SNPs using conditional & joint association analysis (COJO) as part of the Genome-wide Complex Trait Analysis (GCTA) software^77^. We first randomly sampled 50,000 males and females for determining LD structure for autosomal variants. We also randomly sampled 50,000 females for determining LD structure for variants on chromosome X. We next subset variants with *P* < 1 × 10^−6 78^ from the imputation file in BGEN format using the bgenix and qctool modules as part of the BGEN library toolkit^79^, and then converted the file format to PGEN using PLINK 2^80^. Next, stepwise model selection procedure, implemented by COJO, was used to select independently associated SNPs.

For CH-UD loci which overlapped with driver CH GWAS or mCA GWAS loci, we further identified novel variants in the loci of the former that were independent of variants in the loci of the latter. For each WGS batch, we fitted variants within CH-UD loci using Firth logistic regression implemented by SAIGE conditioning on each SNP within driver CH GWAS or mCA GWAS loci. Summary statistics from each WGS batch were then meta-analysed using METAL software as described later (see “Genetic association meta-analysis”). Novel variants were defined with *P* < 5 × 10^−8^. COJO was next implemented as described earlier to identify independent variants among the novel variants.

SNP-based heritability was estimated using linkage disequilibrium score regression (LDSC) software^81^, and the genetic correlation between pairs of CH classes were assessed for CH classes that demonstrated statistically significant heritability estimates (*P* < 0.05).

### Exome-wide association analysis

Exome-wide association analysis was performed on WES-identified germline variant calls for each WGS batch separately using Firth logistic regression, as implemented in SAIGE, adjusted for age, sex, smoking, *peddy*-inferred European probability, and first ten genetic PCs. The genetic relationship matrix computed earlier for the purpose of GWAS was also accounted for here to adjust for population stratification.

### Gene-collapsing association analysis

Eleven different sets of qualifying variant models were assessed on the basis of variant effect on protein-coding sequence, MAF from gnomAD and internal test cohort, Rare Exome Variant Ensemble Learner (REVEL) score^82^, and Missense Tolerance Ratio (MTR)^83^. The qualifying variant (QV) models were “syn” (synonymous, MAF ≤0.005%), “flexdmg” (non-synonymous, MAF_global_ ≤0.05%, MAF_any given ancestry_ ≤0.1%, REVEL score ≥0.25), “flexnonsynmtr” (non-synonymous, MAF_global_ ≤0.05%, MAF_any given ancestry_ ≤0.1%, MTR <0.78 or MTR centile <50), “UR” (non-synonymous, MAF_global_ = 0%, REVEL score ≥0.25), “URmtr” (“UR” and MTR <0.78 or MTR centile <50), “raredmg” (missense, MAF_global_ ≤0.005%, REVEL score ≥0.25), “raredmgmtr” (“raredmg” and MTR <0.78 or MTR centile <50), “ptv” (protein-truncating, MAF_global_ ≤0.1%, MAF_any given ancestry_ ≤0.1%), “ptv5pcnt” (protein-truncating, MAF_global_ ≤5%, MAF_any given ancestry_ ≤5%), “ptvraredmg” (“ptv” and “raredmg”), and, “rec” (non-synonymous, MAF_global_ ≤0.5%, MAF_any given ancestry_ ≤0.5%). All qualifying variants were assessed in a dominant model, with the exception of “rec”, which was assessed in a recessive model.

Gene-collapsing analysis was performed on WES-identified germline variant calls for each WGS batch separately using Firth logistic regression, as implemented in SAIGE, adjusted for age, sex, smoking, *peddy*-inferred European probability, and first ten genetic PCs. The genetic relationship matrix computed earlier for the purpose of GWAS was also accounted for here to adjust for population stratification.

### Genetic association meta-analysis

For GWAS, variants with MAF ≥1%, minor allele count (MAC) ≥5 in both cases and controls, and imputation score >0.6 were then meta-analysed across all WGS batches using the inverse variance weighted (IVW) approach implemented by the METAL software^84^. Cochran’s Q-test was additionally performed to assess for variant-level heterogeneity^85^. Genome-wide significant variants were defined with *P* < 5 × 10^−8^ and *P*_heterogeneity_ ≥ 0.05.

For ExWAS and gene-collapsing analysis, variants with MAC ≥1 in both cases and controls were meta-analysed across all WGS batches using IVW approach. Meta-analysis was additionally performed using *P* value approach (Stouffer’s method) because it is more robust for combining summary statistics with the large effect size and standard error expected by rare variants. Both IVW and *P* value approaches were implemented by the METAL software^84^. Cochran’s Q-test was additionally performed to assess for variant-level heterogeneity^85^. Study-wide significant variants and gene-collapsing models were defined with *P* < 1 × 10^−8^ and *P*_heterogeneity_ ≥0.05 from both IVW and *P* value approaches.

### Inflation assessment for genetic association analyses

For GWAS, genomic inflation factor was computed using the LDSC software and the inflation factor was represented by the LDSC intercept^86^. Pre-computed LD scores from 1000 Genomes European data were used as the reference panel. For GWAS of singleton burden, only variants with MAF ≥1%, imputation score >0.9, and overlapped with HapMap3 variant list were included for LDSC analysis. For GWAS of CH, we further retained variants with *P*_heterogeneity_ ≥ 0.05 from the meta-analysis and computed the LDSC intercept for each WGS batch separately and for the meta-analysis.

For ExWAS and gene-collapsing analysis, genomic inflation factors were computed with permutation approach. Specifically, the association analyses were repeated with the case and control groups randomly assigned to each individual to obtain the permuted *P* values for genomic inflation assessment for each WGS batch separately and for the meta-analysis. The λ_median_ was used as the measure of inflation.

No notable genomic inflation, under the expectational of polygenic inheritance and meta-analysis^87^, was observed for GWAS of singleton burden (λ_GC_=1.08), or GWAS, ExWAS, and gene-collapsing analysis of CH (Supplementary Table 7, 13, and 15). Specifically for CH-UD, GWAS λ_GC_ was 1.17, ExWAS of rare variants λ_median_ was 1.05, and mean λ_median_ across all gene-collapsing models was 1.00 (range = 0.93-1.04). For CH subtypes that demonstrated λ > 1.1, we adjusted the test statistics using the λ, specifically the -log10(*P* value) was divided by the respective λ.

### Plasma protein association analysis

Olink proteomics study cohort from UK Biobank (UKB) was described in detail in previous publications^88–90^. Linear regression was used to model the association between plasma protein measurements and CH adjusted for age, sex, smoking, waist circumstance, BMI, Olink batch, time between sample collection and plasma protein measurement, and first four *peddy*-inferred genetic PCs. Each CH class was compared to non-CH. In total, 2,941 assay measurements were included for analysis, based on Olink antibody pairs, representing 2,923 unique protein targets. Significant associations were defined with *P* < 1.71 × 10^−5^ (multiple testing correction: 0.05/2,941).

To identify biological pathways associated with each CH, we took the negative log10 transformation of each *P* value and indicated the corresponding direction of the beta coefficient (positive value for beta coefficient ≥ 0 and negative value for beta coefficient < 0). The plasma proteins were then ranked based on this test statistics in descending order. Therefore, plasma proteins were ranked from being most elevated to most depleted in CH individuals relative to non-CH individuals. This ranked list was used in a gene set enrichment analysis (GSEA) to identify biological pathways associated with CH. The pathways included for analysis here were that of Gene Ontology biological processes^91^, retrieved from the human Molecular Signature Database (MSigDB) collections^92^, and the *fgsea* R package^93^ was used to assess the enrichment of these pathways in the ranked list of plasma proteins.

### Bayesian hierarchical model of stochastic *TCL1A* activation and clonal expansion

We developed a Bayesian hierarchical model (BHM) of *TCL1A* activation and clonal expansion, and subsequently compared the model’s prediction to that observed among UKB participants with TCL1A plasma protein and singleton data available.

The model is based on the assumption that HSCs undergo stochastic epigenetic switching between a “neutral” (wild-type) state and an “active” (*TCL1A*-activated) state. In the active state, cells gain a fitness advantage (*s*). The transitions are governed by per-division activation (*μ*) and deactivation (*γ*) rates. The model couples two distinct signals: (1) singleton burden representing cell divisions, which act as a molecular clock and (2) singleton-derived VAF representing the size of specific clonal expansions, which capture the selective advantage of active clones. We defined the model as:

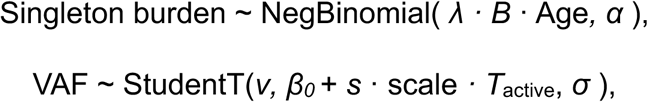

where *λ*, is the passenger mutation rate, *B* is the birth rate, *T*_active_ is the cumulative time a lineage has spent in the active state, governed by the activation rate *μ* and decaying rate:

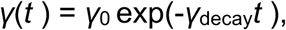

and the scale parameter represents the magnitude of clonal expansion per unit of selective advantage. Specifically, it maps the integrated fitness advantage (*s · T*_active_) to the observed relative clonal expansion size, with *β*_0_ the global intercept.

### Phenome-wide association analyses

We performed association analysis between CH and 13,225 binary phenotypes^37^. Specifically, we used Firth logistic regression model with CH as the predictor and binary phenotype as the outcome adjusted for age, sex, smoking, waist circumferences, body mass index, and first four *peddy*-inferred genetic PCs. Significant associations were defined with *P* < 3.78 × 10^−6^ (0.05 /13,225).

We also performed association analysis between CH and 1,682 quantitative phenotypes^37^. Specifically, we used linear regression model with CH as the predictor and quantitative phenotype as the outcome adjusted for age, sex, smoking, waist circumferences, body mass index, and first four *peddy*-inferred genetic PCs. Significant associations were defined with *P* < 2.97 × 10^−5^ (0.05 /1,682).

Both binary and quantitative phenotypes were taken from the April 2023 data release that was accessed on 28th June 2024 as part of UKB applications 68601 and 65851. To parse the UKB phenotypic data, we adopted our previously described PEACOK package^37^, located at https://github.com/astrazeneca-cgr-publications/PEACOK. Generation of the composite phenotypes (“unions” prefixed) as previously described^37^.

### *CALR* and additional *JAK2* V617F variant detection

To explore the possibility that the association between CH-UD with ET detected from PheWAS may be explained, in part, by hitherto unidentified driver CH variants within individuals with CH-UD, we performed somatic mutation calling for *CALR* and additional *JAK2* V617F variants.

For *CALR*, we detected two canonical hotspot variants, namely type I and type II variants. Type I variant is characterised by 52bp deletion (GCAGAGGCTTAAGGAGGAGGAAGAAGACAAGAAACGCAAAGAGGAGGAGGAG) whereas type II variant is characterised by 5bp insertion (TTGTC). Both variants are located on exon 9 and were detected using Mutect2 as described above (“Driver CH detection”). For *JAK2* V617F, we performed *in silico* genotyping at chr9: 5073770 using the perbase software (https://github.com/sstadick/perbase) with the following parameters: *--keep-zeros --min-mapq 1 --exclude-flags 1796*. Several genotype groups were created on the basis of a combination of minimum number of reads (1, 2, or ≥3) and VAF (<3%, 3-40%, or >40%) for benchmarking purpose. PheWAS were conducted with these newly detected *CALR* and *JAK2* V617F mutation carriers as well as with CH-UD after excluding these mutation carriers. It is noteworthy that Mutect2 detection of *CALR* variants and pileup deletion of *JAK2* V617F were not part of the recommended CH detection workflow^43^, and therefore were not included as part of our initial CH call set.

We benchmarked these newly detected *CALR* and *JAK2* V617F variants by assessing the (1) carrier frequency against age, (2) the association between carrier status and age using logistic regression model adjusted for sex, smoking, and first four *peddy*-inferred genetic principal component, (3) association between carrier status and PV or ET using Firth logistic regression model adjusted for age, sex, smoking, waist circumferences, body mass index, and first four *peddy*-inferred genetic PCs, and (4) association between carrier status and platelet crit using linear regression model adjusted for age, sex, smoking, waist circumferences, body mass index, and first four *peddy*-inferred genetic PCs.

### Inflation assessment for plasma protein and phenome-wide association analyses

Test statistic inflation factors were computed using the permutation approach. Specifically, the association analyses were repeated with the case and control groups randomly assigned to each individual to obtain the permuted *P* values. The λ_median_ was used as the measure of test statistic inflation. For CH classes that demonstrated λ_median_ > 1.1, we adjusted the *P* values by dividing this test statistic with the corresponding λ_median_ (Supplementary Tables 17 and 22). For CH-UD, we noted λ_median_ of 1.07, 1.59, and 1.40 for plasma protein, binary phenotype, and quantitative phenotype, respectively. For CH classes that demonstrated λ > 1.1, we adjusted the test statistics using the λ, specifically the -log10(*P* value) was divided by the respective λ.

### Mendelian randomisation

We assessed CH as the outcome with smoking (cigarettes per day and initiation), alcohol use, waist-to-hip ratio (BMI-adjusted), and leukocyte telomere length as the exposures. Genetic instruments and corresponding genetic association summary statistics for the exposures were derived from previous reports^2,78,94,95^.

We performed bi-directional Mendelian randomisation (MR) with plasma protein level as the exposure and CH as the outcome, and conversely, CH as the exposure and plasma protein level as the outcome. Genetic instruments for CH, consisting of genome-wide significant independent SNPs, were retrieved from this study (see “Genome-wide association analysis of common germline variants”) while genetic instruments for plasma protein levels, consisting of independent pQTL signals, were retrieved from UKB flagship plasma protein study^89^. We focused on CH-UD, driver CH, and *DNMT3A*-CH where there were reasonable number of genetic instruments and on plasma proteins that were associated with these CH classes at Bonferroni threshold. We additionally assessed the genetic liability of TCL1A plasma protein level on driver CH, *DNMT3A*-, *TET2*-, and *ASXL1*-CH although the association between TCL1A plasma protein level with these CH classes were not significant at Bonferroni threshold. This is because a previous study demonstrated a causal role for TCL1A in clonal expansion driven by these CH genes^14^. It is noteworthy that rs2887399 was removed from the genetic instruments of CH-UD when this CH class was assessed as the exposure on TCL1A plasma protein. This is because this SNP has been shown to directly affect *TCL1A* expression^14^, therefore the inclusion of this SNP in MR would violate the exclusion restriction assumption which assumes the instrument does not influence the outcome of interest directly^96^.

In a MR-PheWAS, binary and quantitative phenotypes associated with CH-UD at Bonferroni thresholds were assessed using MR with FinnGen and UKB (https://www.nealelab.is/uk-biobank), respectively. Specifically, CH-UD was defined the exposure and phenotypes were defined the outcomes.

MR was performed using the *TwoSampleMR* R package^97,98^ (valid in both one-sample and two-sample setting^99^). Significant associations were defined with *P* < 0.05 derived using the median of IVW, MR Egger, and weighted median approaches^100^. Specifically, for each association, we multiplied the *P* values by the direction of effect size (1 for positive and −1 for negative beta coefficients, respectively), ranked this test statistic, and reported the median test statistic and corresponding effect size and *P* value. Therefore, significant results represent associations supported by at least two MR models with significant *P* values and same direction of beta coefficients.

### Survival analysis

Association between all-cause mortality and CH was modelled using Cox proportional hazards regression adjusted for age, sex, smoking, first four *peddy*-inferred genetic PCs. Modelling was performed using the *coxph* function from the *survival* R package. The vital status of UKB participants for this analysis was censored on 31^st^ May 2024.

### Statistics and reproducibility

Except where specific software packages are named, all statistical analyses and plotting were performed using R (v4.2.2). No statistical methods were used to predetermine sample size.

## Data availability

Summary statistics for significant results from GWAS, ExWAS, gene burden association analysis, baseline phenotype association analysis, and mortality analysis are provided in the Supplementary Tables. Individual-level UKB data may be requested via applicable to the UKB. Full summary statistics for GWAS, ExWAS, gene burden association analysis will be provided upon publication of the manuscript.

## Code availability

The code for the Bayesian hierarchical model of stochastic TCL1A activation and clonal expansion code is available at https://github.com/vesmanojlovic/tcl1aBHMAll. All other analyses were performed using publicly available software and web-based applications as indicated in the Methods section.

## Acknowledgement

Our sincere thanks to the participants and researchers of the UKB, who made this effort possible.

---

**Extended Data Figure 1:**
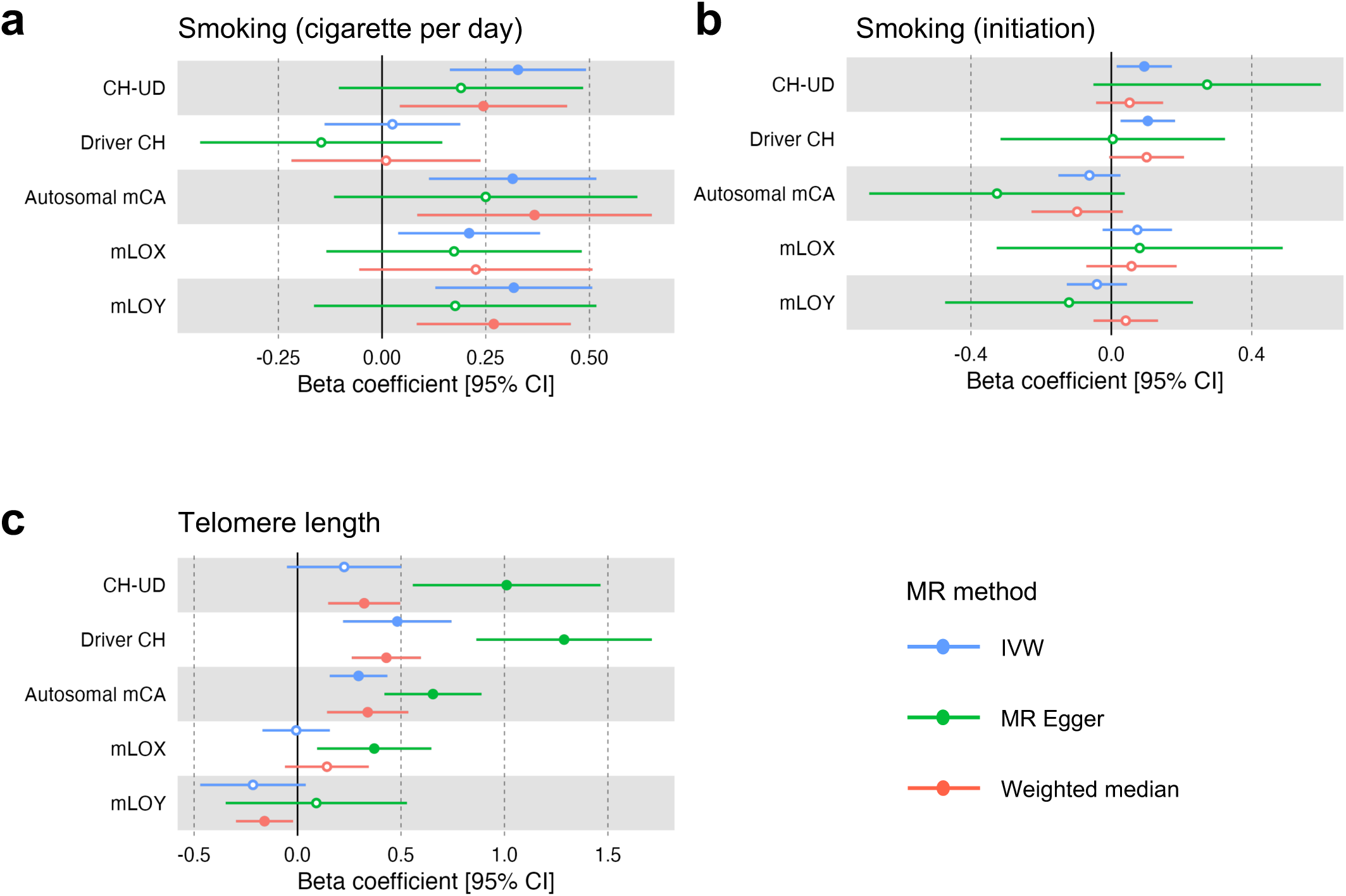
Mendelian randomisation with CH as the outcome and risk factors as the exposures. **a-c,** Mendelian randomisation with CH as the outcome and number of cigarettes per day **(a)**, smoking initiation **(b)**, or leukocyte telomere length **(c)** as the exposures. Beta coefficients were derived from three different MR methods, namely, inverse variance weighted, MR Egger, and weighted median. Solid circles represent associations with *P* < 0.05; hollow circles represent associations with *P* ≥ 0.05. IVW, inverse variance weighted.

**Extended Data Figure 2:**
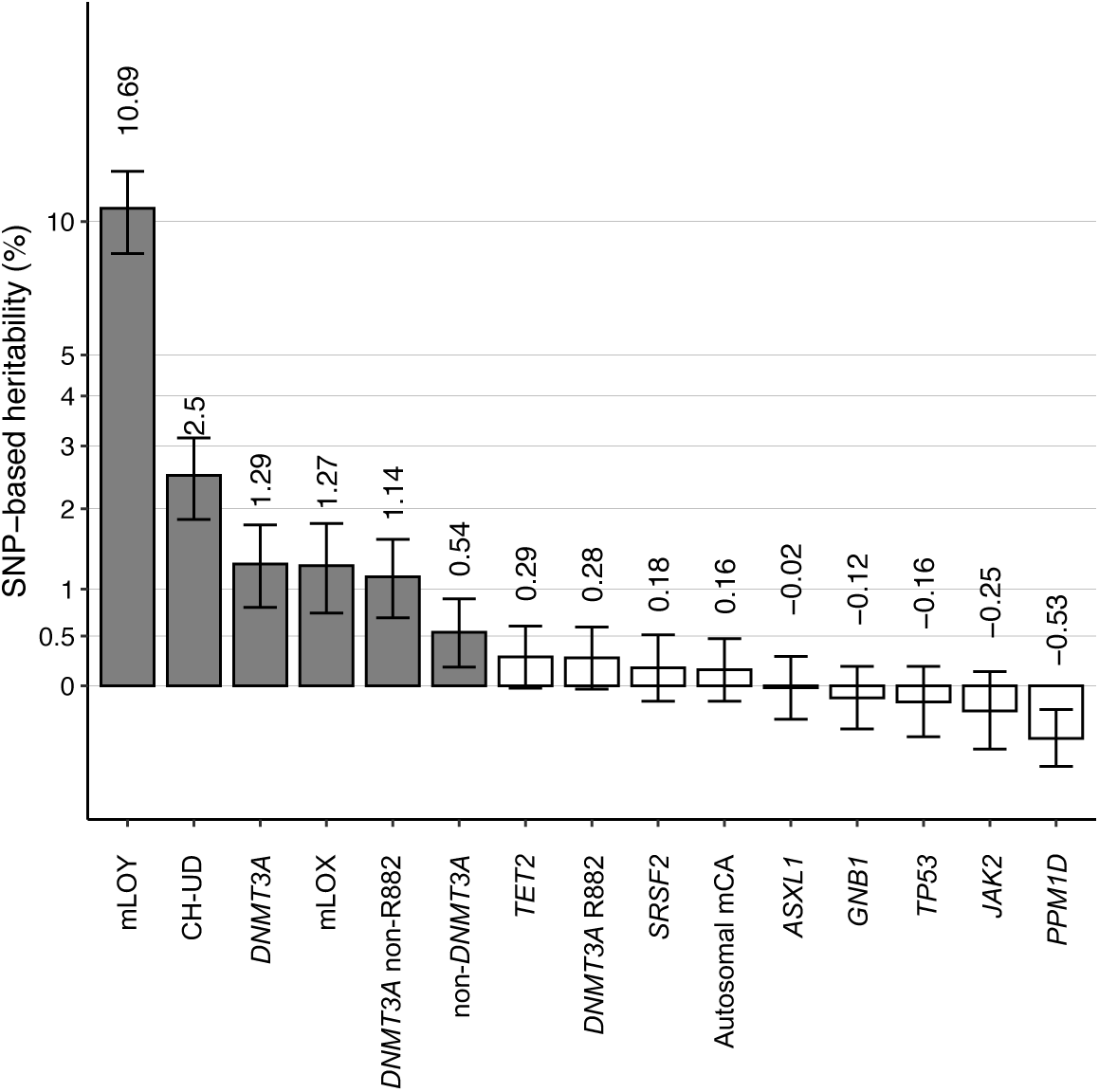
SNP-based heritability estimates of the different CH subtypes. Heritability estimates computed using LDSC software. Gray boxes indicate CH subtypes where the lower and upper bound of the 95% CI of the estimates are above zero (statistically significant estimates). White boxes indicate CH subtypes where either the lower and/or the upper bound of the 95% CI of the estimates are/is below zero (non-statistically significant estimates).

**Extended Data Figure 3:**
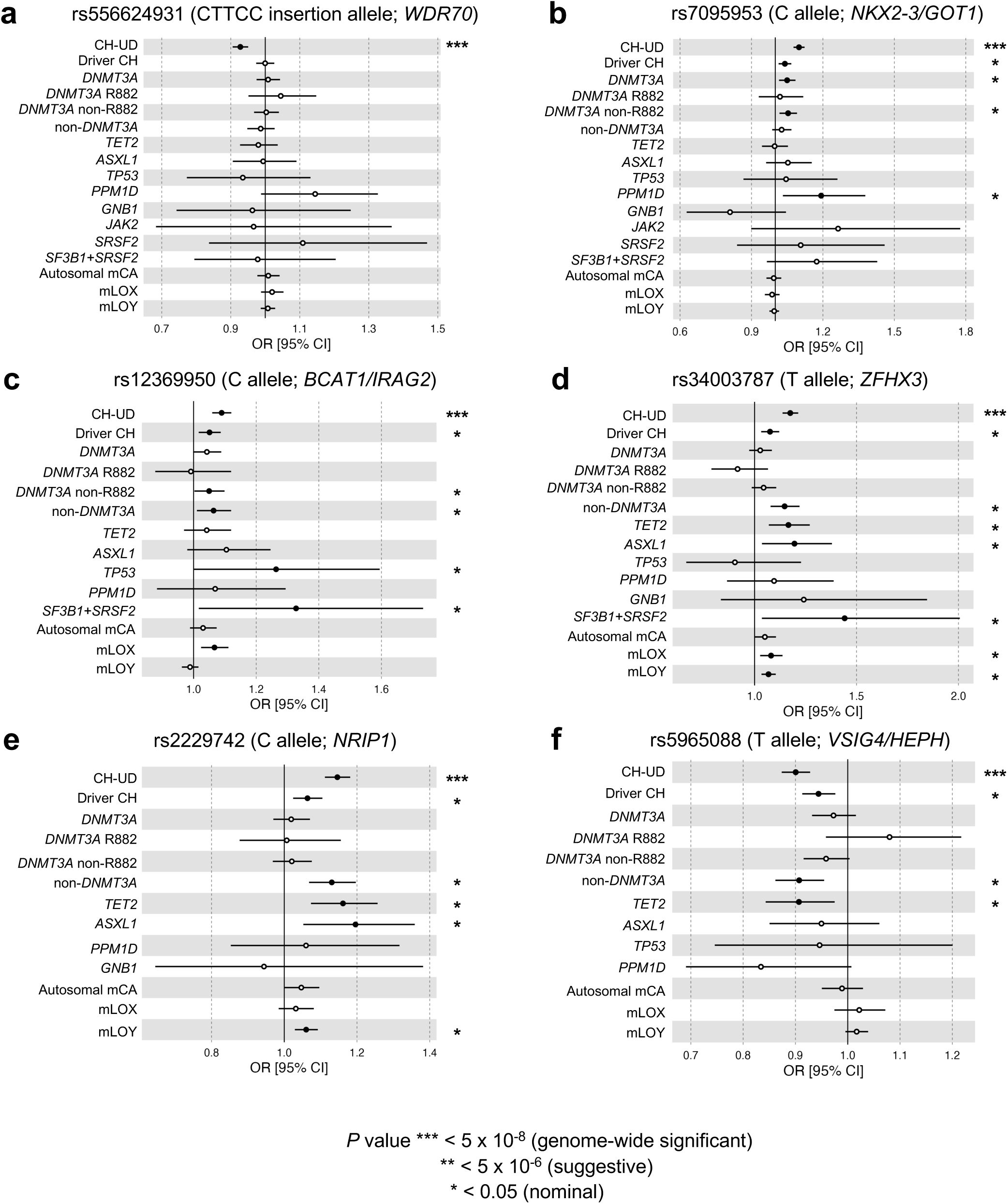
GWAS summary statistics across the different CH subtypes for leading SNPs of loci specific to CH-UD. Odds ratio and *P* values derived from METAL software. Firth logistic regression implemented using SAIGE software was ran for each WGS batch separately, and then meta-analysed using METAL. For SAIGE, we specified age, sex, smoking, *peddy*-inferred European probability, and first ten genetic principal components as co-variates. For each SNP, only CH subtypes with minor allele count (MAC) ≥1 in both cases and controls in each WGS batch were included here. *P* *** < 5 × 10^−8^ (genome-wide significant) ** < 5 × 10^−6^ (suggestive) * < 0.05 (nominal).

**Extended Data Figure 4:**
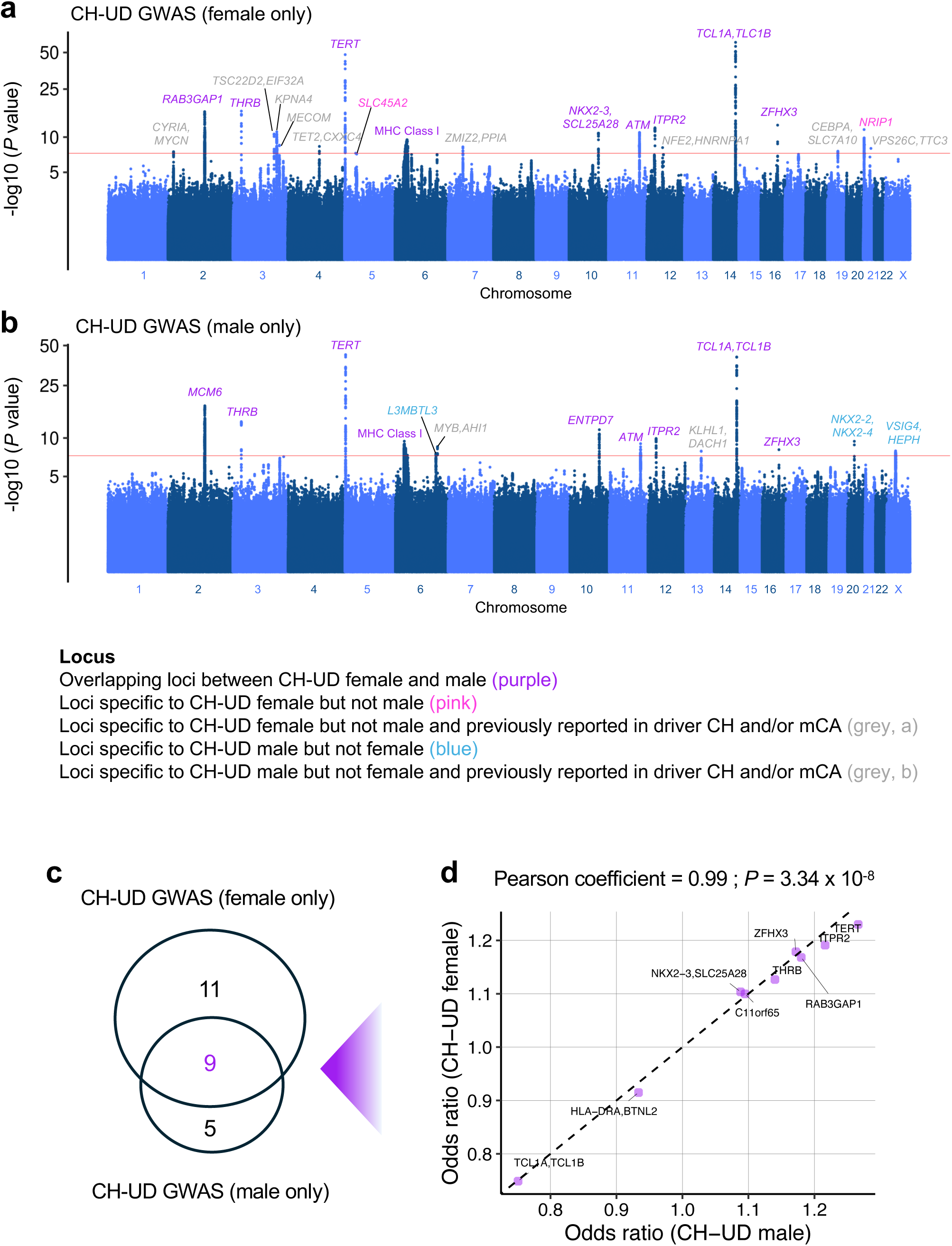
Sex-stratified CH-UD GWAS. **a,b,** Manhattan plot representing the common germline variants included for CH-UD GWAS among females only **(a)** and males only **(b)**. *P* values on the y-axis were derived from METAL software. GWAS was ran on each WGS batch separately using Firth logistic regression implemented using SAIGE software, and then meta-analysed using METAL. For SAIGE, we specified age, sex, smoking, *peddy*-inferred European probability, and first ten genetic principal components as co-variates. **(c)** Number of loci overlapping between female- and male-only CH-UD GWAS and number of loci specific to female- or male-only CH-UD GWAS. **(d)** Correlation between odds ratio of overlapping loci between female- and male-only CH-UD GWAS.

**Extended Data Figure 5:**
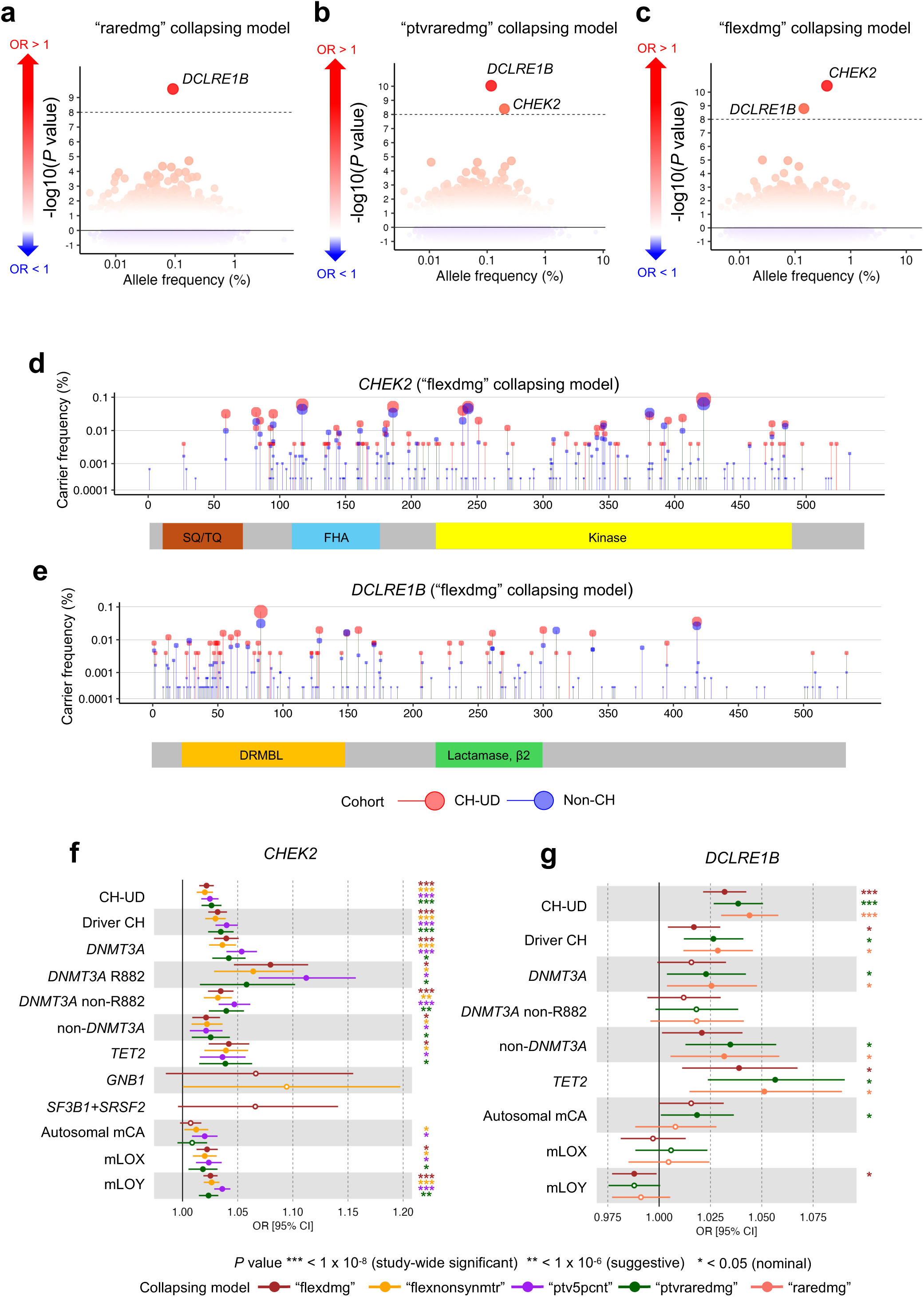
Gene-collapsing analysis of CH-UD. **a-c,** Gene-collapsing analysis of CH-UD for rare variants that qualified as “raredmg” **(a)**, “ptvraredmg” **(b)**, or “flexdmg” **(c)** aggregated at the gene level. Genes are ordered by population allele frequency on the x-axis and -log10(*P* value), with direction of odds ratios, on the y-axis. The dashed line indicates the study-wide significant threshold (*P* < 1 × 10^−8^). *P* values on the y-axis were derived from METAL software. ExWAS was ran on each WGS batch separately using Firth logistic regression implemented using SAIGE software, and then meta-analysed using METAL. For SAIGE, we specified age, sex, smoking, *peddy*-inferred European probability, and first ten genetic principal components as co-variates. **d,e,** Amino acid position distribution and carrier frequency of rare germline variants for *CHEK2* **(d)** and *DCLRE1B* **(e)** stratified by CH-UD and non-CH group. **f,g,** Odds ratio of *CHEK2* **(f)** and *DCLRE1B* **(g)** across the different CH subtypes for collapsing models that were study-wide significant from gene-collapsing analysis of CH-UD. Only CH subtypes whose collapsing model with minor allele frequency (MAF) ≥1 in both cases and controls in each WGS batch were included here. DRMBL, DNA repair metallo-beta-lactamase; FHA, forkhead-associated domain; SQ/TQ, serine-glutamine/threonine-glutamine cluster domain. *P* *** < 1 × 10^−8^ (study-wide significant) ** < 1 × 10^−6^ (suggestive) * < 0.05 (nominal).

**Extended Data Figure 6:**
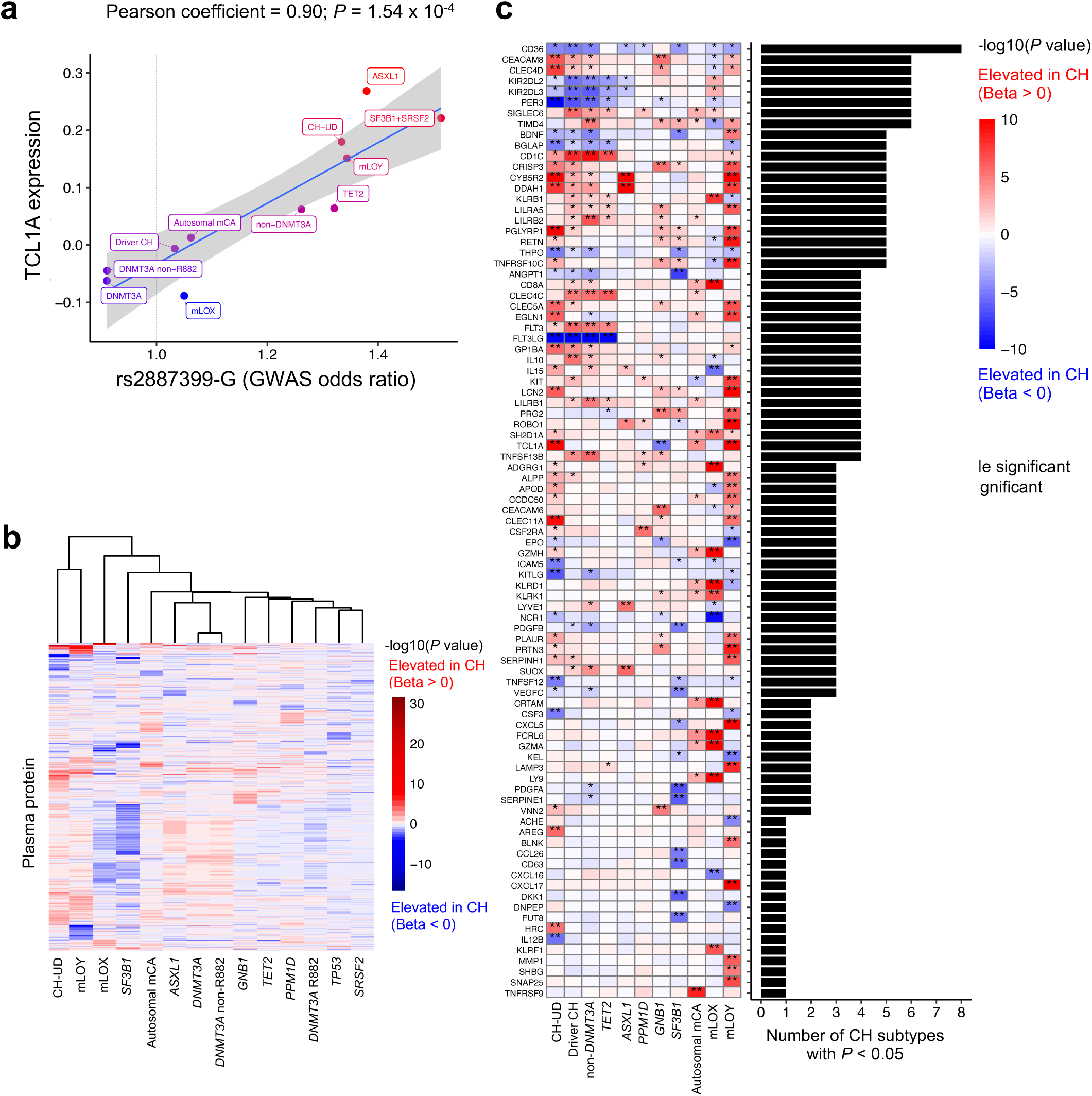
Plasma protein profile of CH-UD relative to other CH subtypes. **a,** Correlation between GWAS effect size of *TCL1A* promoter SNP (G allele of rs2887399) versus median plasma protein levels of TCL1A across the different CH subtypes. Only CH subtypes with nominal association with rs2887399 (*P* < 0.05) were included here. GWAS odds ratio and *P* values derived from METAL software. **b,** Unsupervised clustering of CH subtypes. Only plasma proteins with data available ≥10 CH individuals and CH subtypes associated with ≥1 plasma protein at nominal threshold (*P* < 0.05) were included here. CH subtypes (columns) and plasma proteins (rows) were clustered using Euclidean distance. **c,** *P* values, with direction of coefficient indicated, for plasma proteins across the different CH subtypes. Only plasma proteins with data available ≥10 CH individuals and CH subtypes associated with ≥1 plasma protein significant at Bonferroni threshold (*P* < 1.71 × 10^−5^) were included for analysis here. *P* value derived from linear regression with CH status as the predictor (non-CH as reference) and plasma protein level as the outcome adjusted for age, sex, smoking, waist circumstance, BMI, Olink batch, time between sample collection and plasma protein measurement, and first four *peddy*-inferred genetic principal components. NPX, normalised protein expression.

**Extended Data Figure 7:**
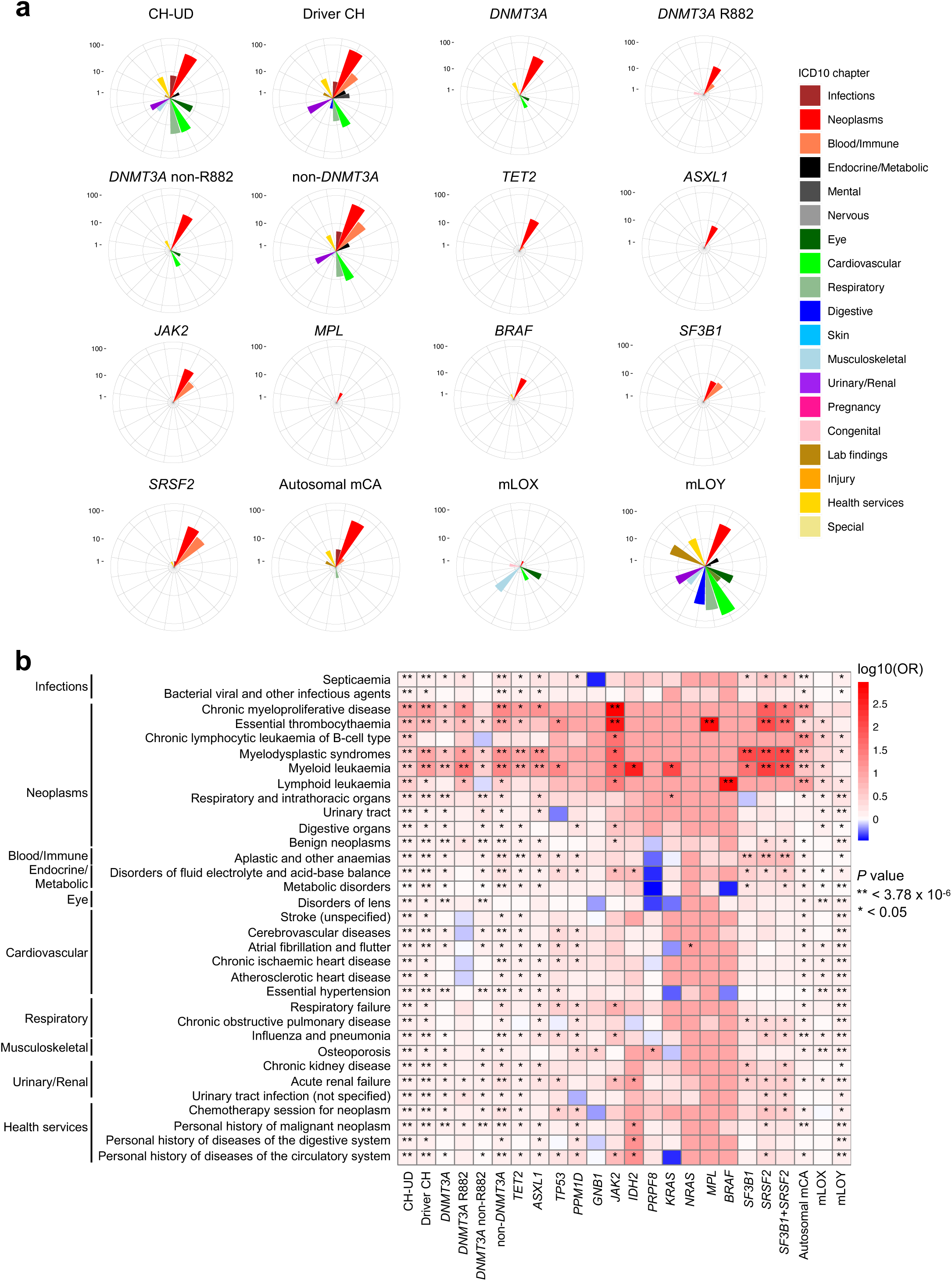
Comparison of binary phenotypes across the different CH subtypes. **a,** Number of significant phenotypes at Bonferroni threshold (*P* < 3.78 × 10^−6^), stratified by ICD10 chapters, across the different CH subtypes. Only CH subtypes with ≥1 significant phenotype shown here. **b,** Odds ratios and *P* values for selected significant phenotypes associated with in CH-UD across the different CH subtypes (related to Figure 6a). Odds ratios and *P* values were derived from Firth logistic regression with trait or disease as the outcome and CH-UD as the predictor (non-CH as reference) adjusted for age, sex, smoking, waist circumferences, body mass index, and first four *peddy*-inferred genetic principal components.

**Extended Data Figure 8:**
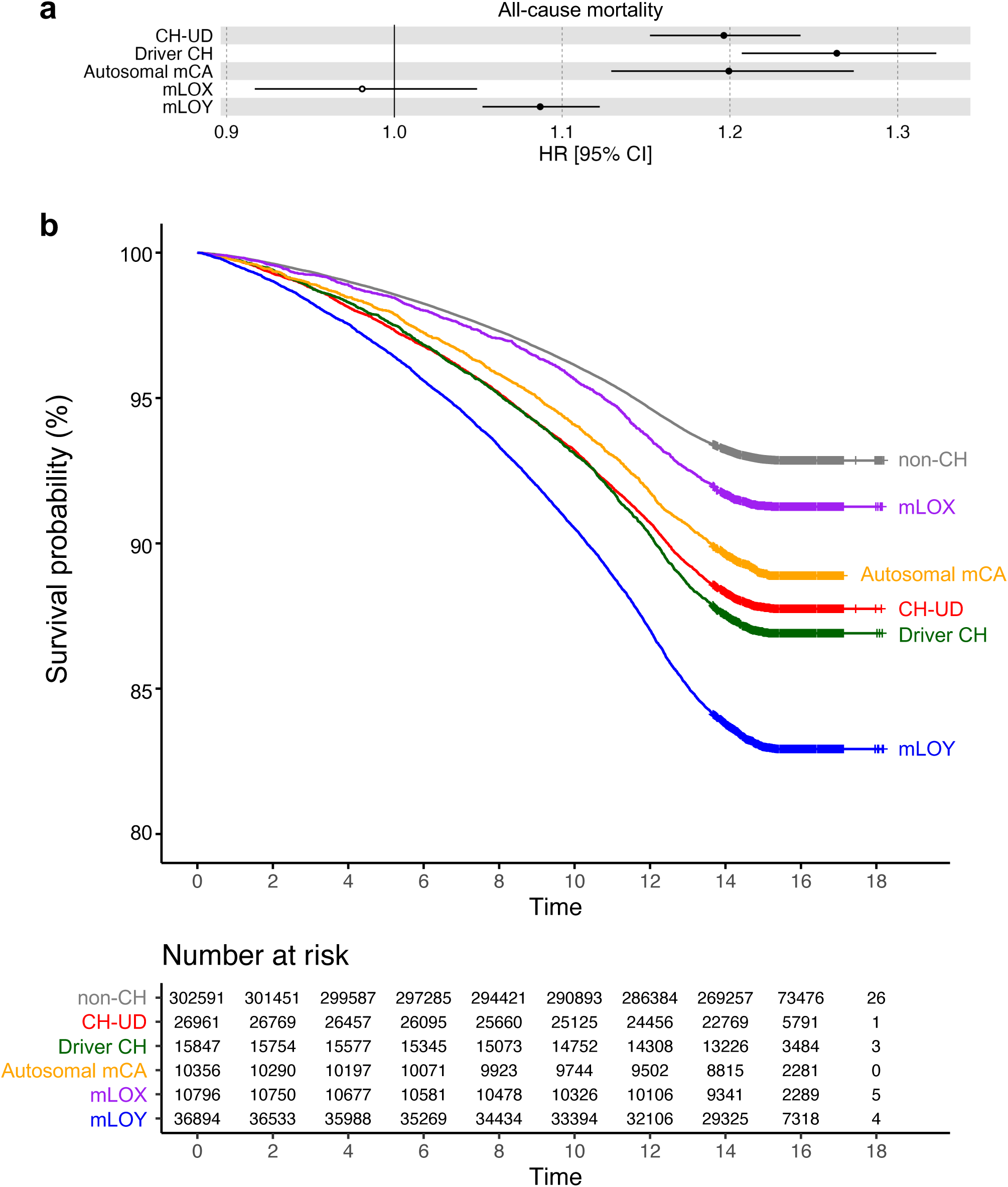
Association between the different CH classes and all-cause mortality. **a,** Hazard ratio and *P* values were derived from Cox proportional hazard regression with age, sex, smoking, first four *peddy*-inferred genetic PCs as covariates. **b,** Survival probabilities of CH and non-CH individuals. The vital status of UKB participants for this analysis was censored on 31^st^ May 2024. The duration is represented by the time between blood sample collection and the censored date.

**Extended Data Figure 9:**
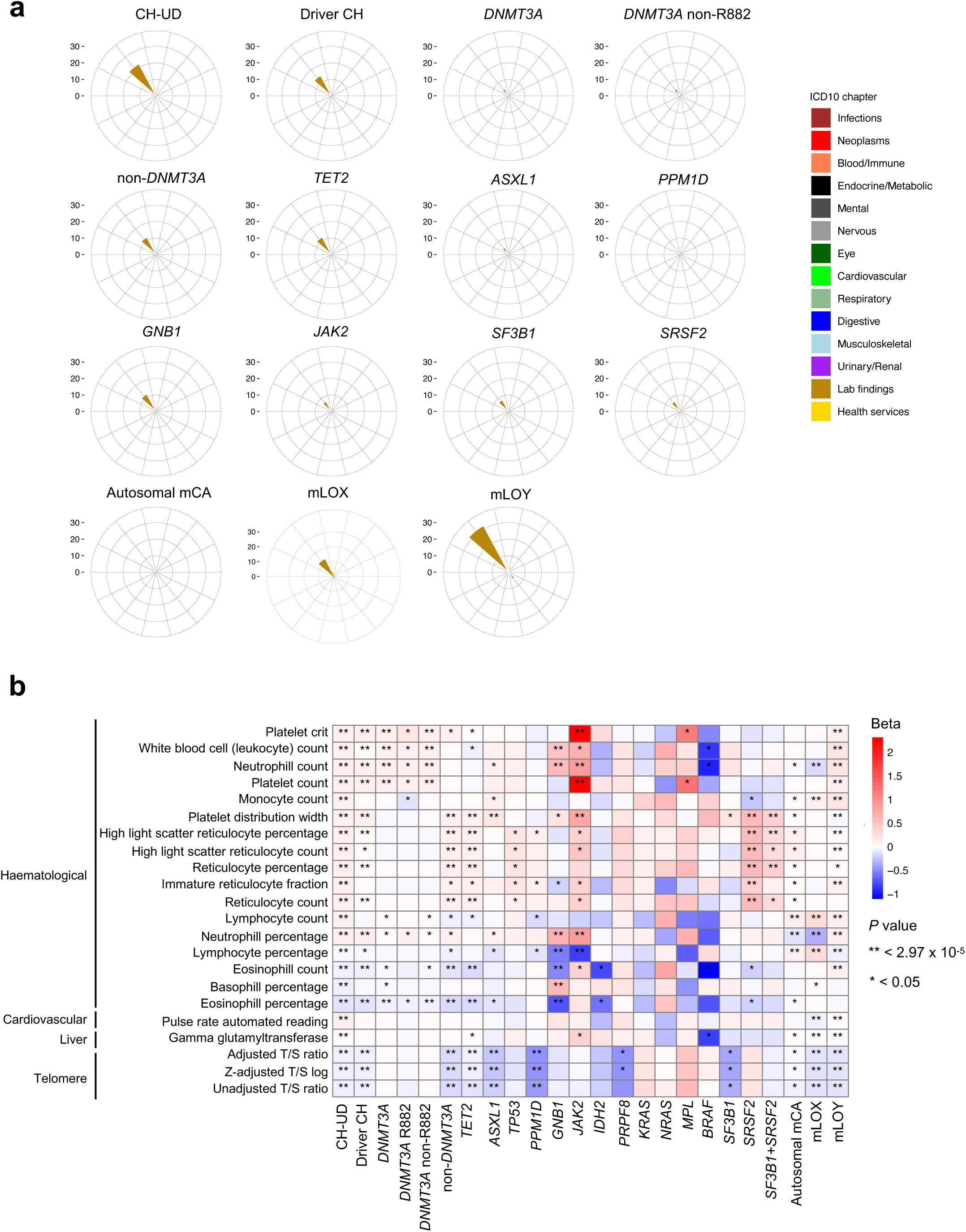
Comparison of quantitative phenotypes across the different CH subtypes. **a,** Number of significant phenotypes at Bonferroni threshold (*P* < < 2.97 × 10^−5^), stratified by ICD10 chapters, across the different CH subtypes. Only CH subtypes with ≥1 significant phenotype shown here. **b,** Beta coefficients and *P* values for selected significant phenotypes associated with in CH-UD across the different CH subtypes (related to Figure 6b). Beta coefficients and *P* values were derived from linear regression with quantitative measurement as the outcome and CH-UD as the predictor (non-CH as reference) adjusted for age, sex, smoking, waist circumferences, body mass index, and first four *peddy*-inferred genetic principal components.

**Extended Data Figure 10:**
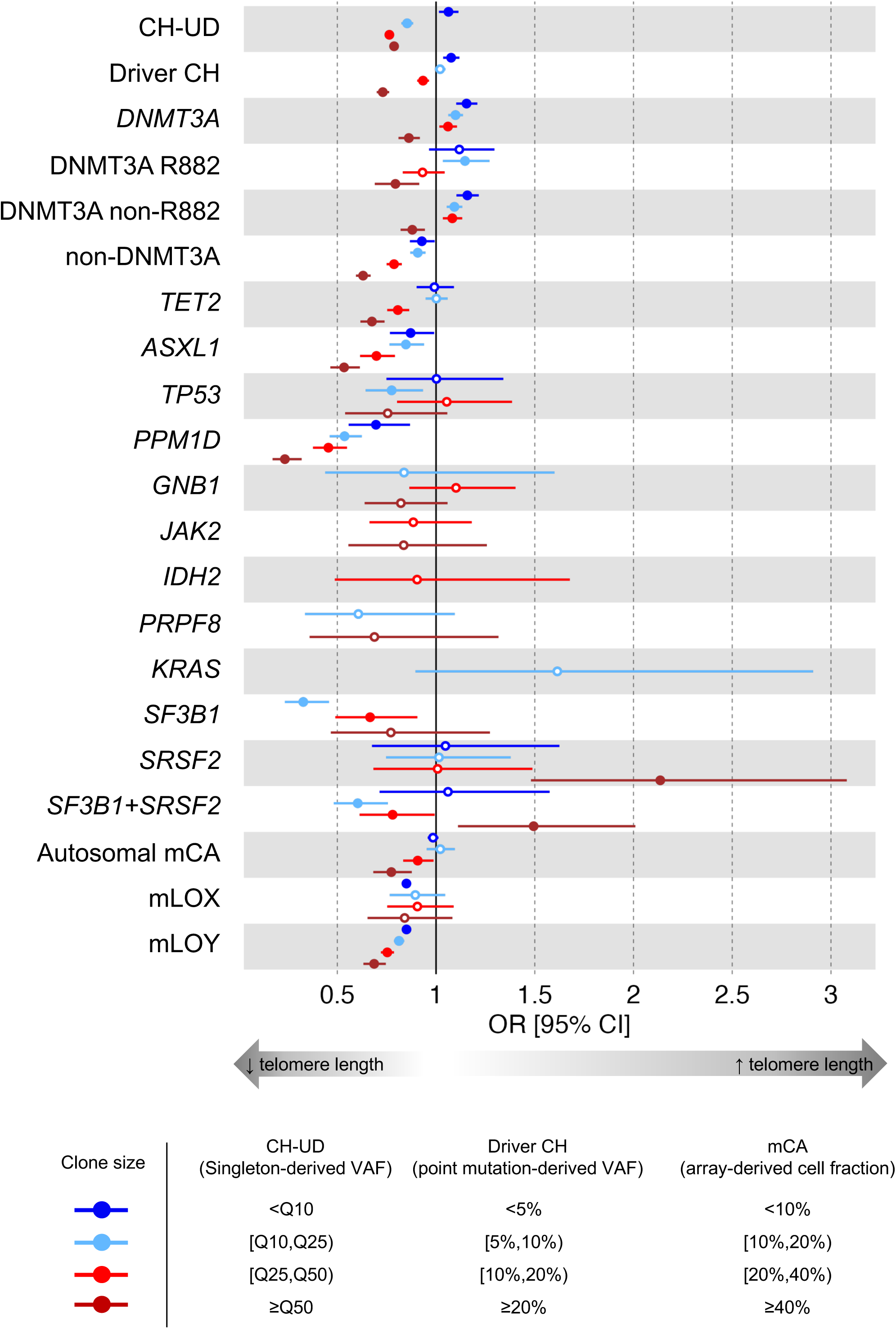
Association between measured leukocyte telomere length and CH stratified by clone size. For CH-UD, the clone size was inferred using singleton-derived cell fraction. For driver CH, the clone size was inferred using the variant allele frequency (VAF) of the driver gene mutation, e.g., *JAK2* V617F. For mCA, the clone size was inferred using the cell fraction determined from the genotyping array. Leukocyte telomere length (LTL) was derived from the first principal component (PC1) of qPCR-inferred telomere length and TelSeq/WGS-inferred telomere length (Burren *et al*.), and then centred and scaled. Odds ratios were derived from logistic regression model with CH as the outcome (non-CH as reference) and LTL PC1 as the main predictor, adjusted for age, sex, smoking, and first four *peddy*-inferred genetic PCs.

