## Supplementary Information for "Clonal haematopoiesis without identified genetic drivers: insights from analyses of 407,512 individuals"

Table of Contents

*Supplementary Figures* ... *3*

Supplementary Figure 1: Selection of singletons from whole-genome sequencing data. ... 5

Supplementary Figure 2: Selection of samples on the basis of ancestry. ... 7

Supplementary Figure 3: Characteristics of singletons stratified by singleton burden percentile. ... 9

Supplementary Figure 4: Characteristics of genomic features of singletons included in the machine learning classifier to detect CH-UD. ... 11

Supplementary Figure 5: Sequencing metrices by whole-genome sequencing batch. ... 13

Supplementary Figure 6: Comparison of different training sets and different machine learning classifiers for the detection of CH-UD. ... 15

Supplementary Figure 7: Validation of detected singletons. ... 17

Supplementary Figure 8: Validation of singleton-derived cell fractions. ... 19

Supplementary Figure 9: Validation of singletons and singleton-derived cell fractions using PACER. ... 21

Supplementary Figure 10: Validation of proteomic-based quantification of biological age and age gap. ... 23

Supplementary Figure 11: Manhattan plots representing the common germline variants included for driver CH, *DNMT3A*-, *DNMT3A* R882-, and *DNMT3A* non-R882-CH GWAS. ... 25

Supplementary Figure 12: Manhattan plots representing the common germline variants included for non-*DNMT3A*-, *TET2*-, and *ASXL1*-CH GWAS. ... 27

Supplementary Figure 13: Manhattan plots representing the common germline variants included for autosomal mCA, mLOX, and mLOY GWAS. ... 29

Supplementary Figure 14: Association between rs2887399 genotype and TCL1A plasma protein level. ... 31

Supplementary Figure 15: Association between age, sex, and smoking with TCL1A plasma protein level. ... 33

Supplementary Figure 16: Proportion of variance of TCL1A protein levels explained by rs2887399 genotype, age, sex, and smoking. ... 35

Supplementary Figure 17: Association between rs2887399 genotype and *TCL1A* promoter methylation. ... 37

Supplementary Figure 18: Age-stratified joint model fit to singleton burden. ... 39

**Supplementary Figure 22: Proposed model on the shared genetic predisposition and clonal dynamics leading up to the detection of CH-UD and non-*DNMT3A* CH.**. 47

### Supplementary Figures

**a**

422,295,944 singletons

↓

| Criteria | Variants (n) | Variants (%) |
|---|---|---|
| VAF ≤ 25% | 8,886,079 | 2.1 |
| 15 ≤ DP ≤ 60 | 416,949,753 | 98.7 |
| MAC ≥ 3 | 421,870,064 | 99.9 |
| Not reported in gnomAD | 420,910,469 | 99.7 |

↓

8,235,783 putative passenger variants

↓

| Criteria | Variants (n) | Variants (%) |
|---|---|---|
| Genotype missingness ≤ 10% | 8,201,225 | 99.6 |
| Autosomal variants | 7,933,615 | 96.3 |

↓

7,908,953 high-quality autosome passenger variants

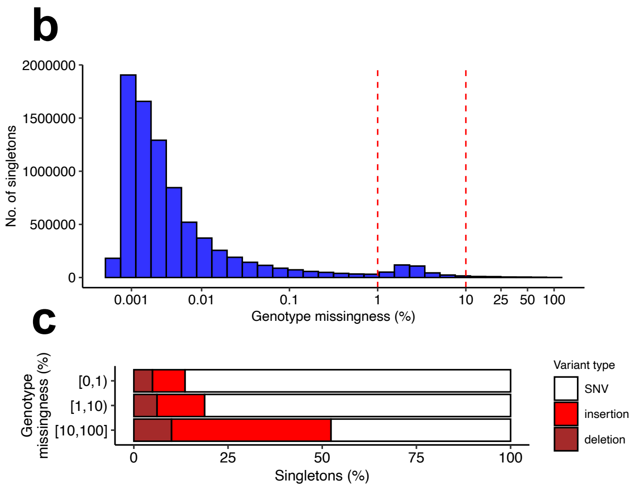

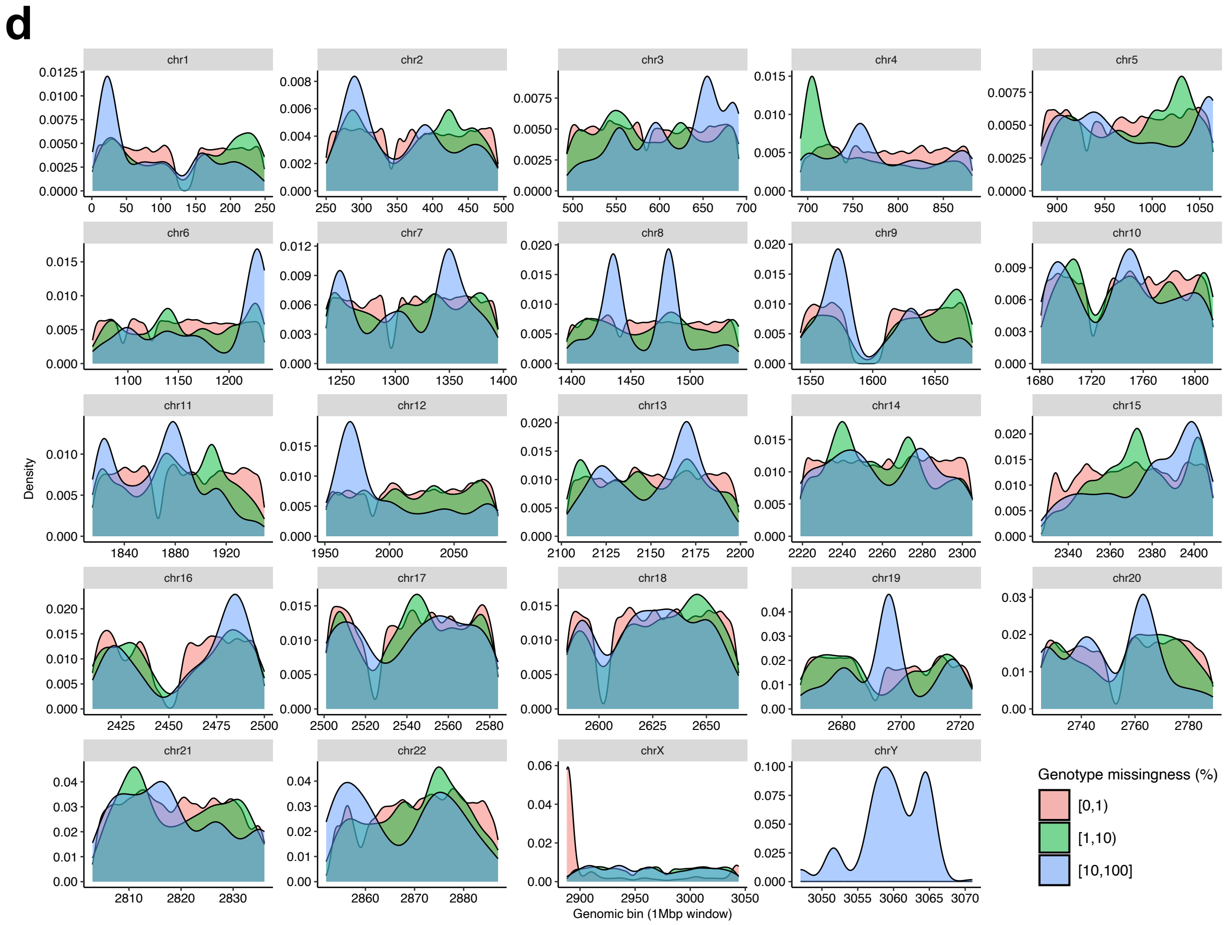

**Supplementary Figure 1: Selection of singletons from whole-genome sequencing data.**

**a,** Singletons selected on the basis of variant allele frequency (VAF), sequencing depth at singleton site, number of reads supporting the alternative allele, gnomAD status, genotype missingness, and chromosome location. **b,c,** Distribution of singletons by genotype missingness rate **(b)** and proportion of SNV, insertion, and deletion by category of genotype missingness rate **(c)**. **d,** Genotype missingness rate across all chromosome position binned by 1Mbp windows. gnomAD, Genome Aggregation Database; MAC, minor allele frequency; SNV, single nucleotide variant; VAF, variant allele frequency.

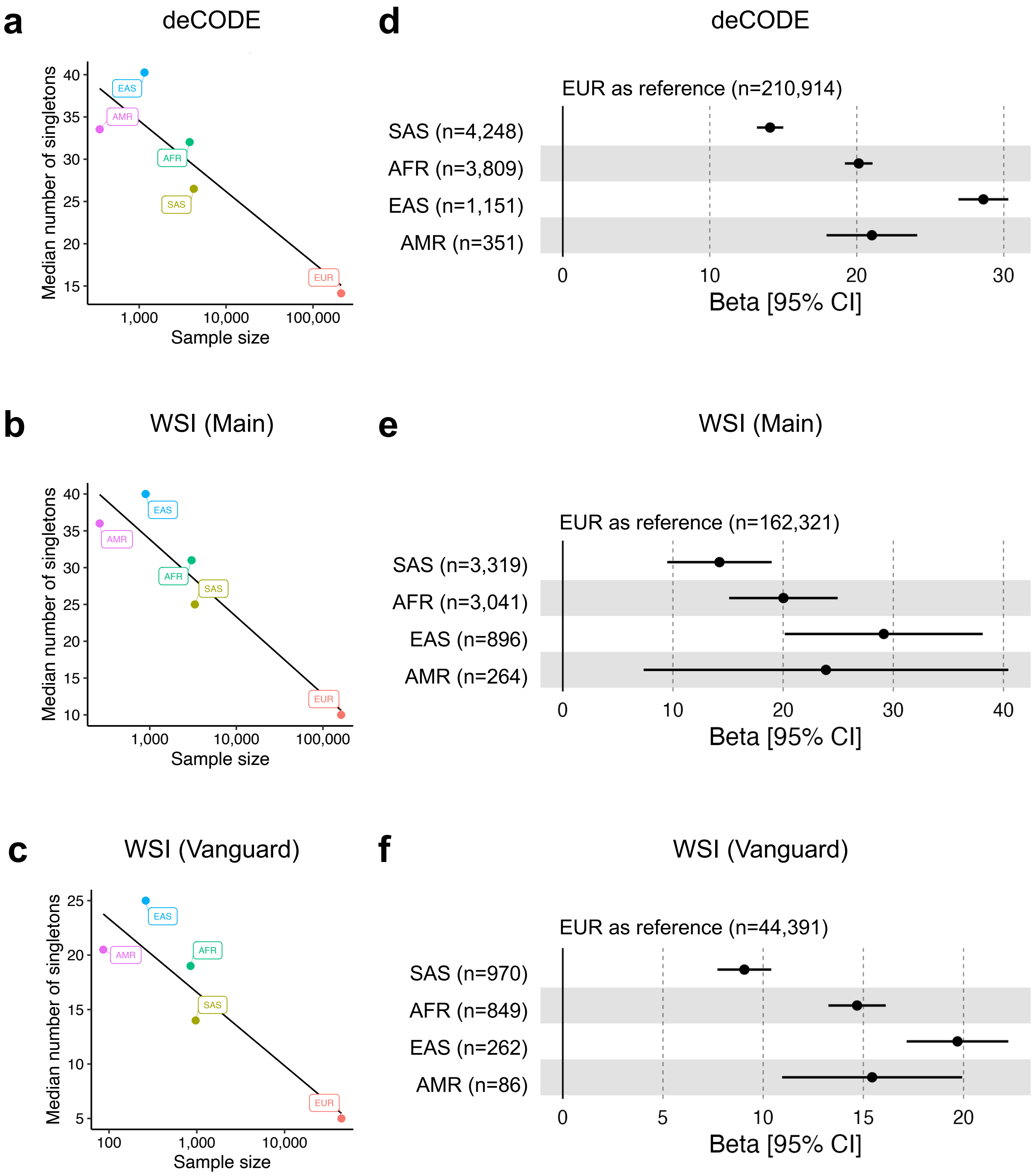

a
deCODE
Median number of singletons
Sample size
EAS
AMR
AFR
SAS
EUR
d
deCODE
EUR as reference (n=210,914)
SAS (n=4,248)
AFR (n=3,809)
EAS (n=1,151)
AMR (n=351)
Beta [95% CI]
b
WSI (Main)
Median number of singletons
Sample size
e
WSI (Main)
EUR as reference (n=162,321)
SAS (n=3,319)
AFR (n=3,041)
EAS (n=896)
AMR (n=264)
Beta [95% CI]
c
WSI (Vanguard)
Median number of singletons
Sample size
f
WSI (Vanguard)
EUR as reference (n=44,391)
SAS (n=970)
AFR (n=849)
EAS (n=262)
AMR (n=86)
Beta [95% CI]

**Supplementary Figure 2: Selection of samples on the basis of ancestry.**

**a-c,** Negative correlation between ancestry group size and the median number of singletons among individuals sequenced by deCODE **(a)**, WSI main **(b)**, and WSI Vanguard **(c)**. **d-f,** Beta coefficients were derived from linear regression with number of singletons as the outcome, ancestry group as the main predictor (European as reference) adjusted for age, sex, and smoking for deCODE **(d)**, WSI main **(e)**, and WSI Vanguard **(f)**. AFR, African; AMR, Admixed American; EAS, East Asian; EUR, European; SAS, South Asian.

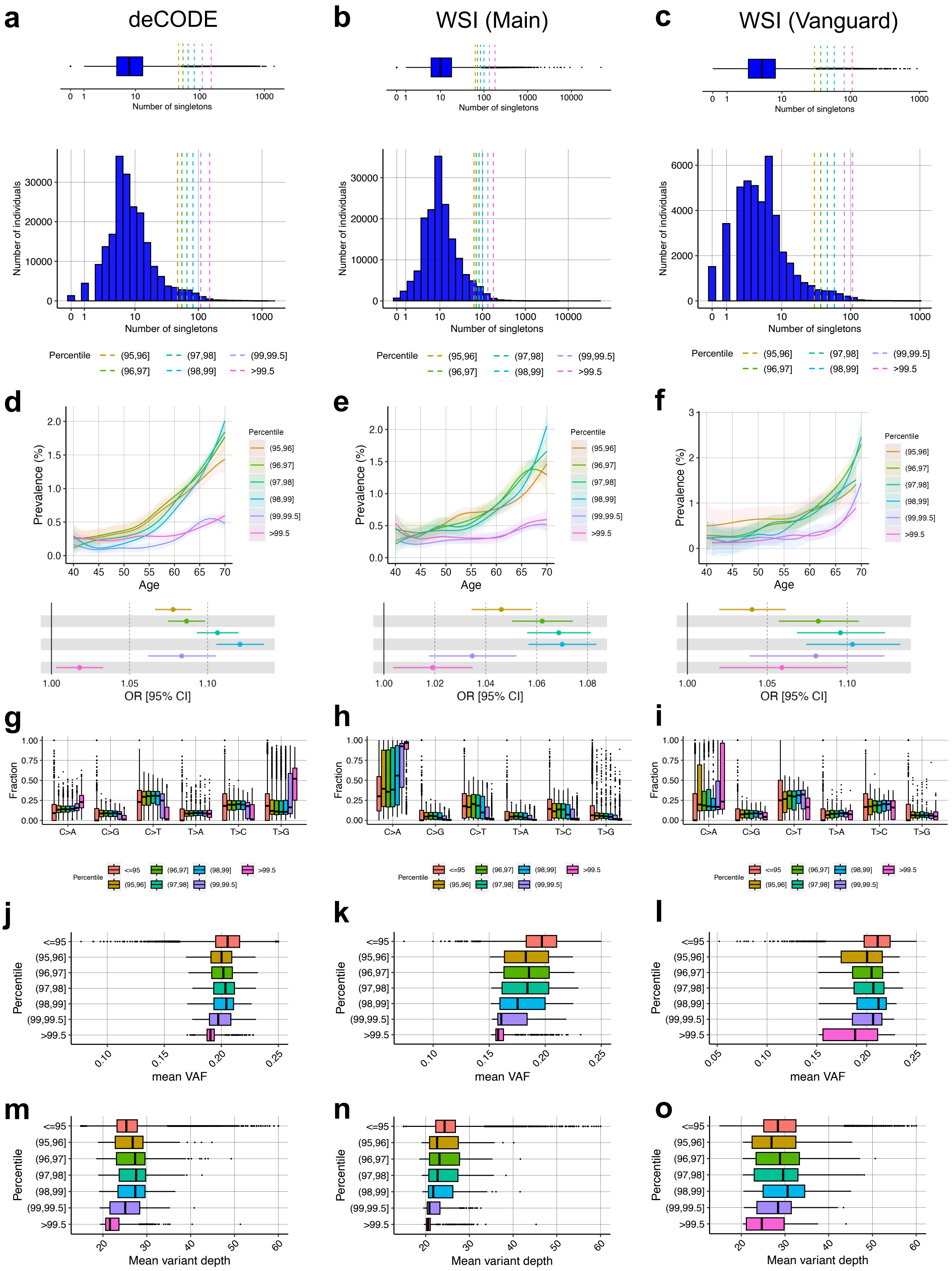
a
deCODE
b
WSI (Main)
c
WSI (Vanguard)
Number of singletons
Number of individuals
Percentile
(95,96]
(96,97]
(97,98]
(98,99]
(99,99.5]
>99.5
d
e
f
Prevalence (%)
Age
OR [95% CI]
g
h
i
Fraction
C>A
C>G
C>T
T>A
T>C
T>G
<=95
j
k
l
mean VAF
m
n
o
Mean variant depth

**Supplementary Figure 3: Characteristics of singletons stratified by singleton burden percentile.**

**a-c,** Distribution of the number of singletons per individual. Selected percentiles indicated on the histogram for deCODE **(a)**, WSI main **(b)**, and WSI Vanguard **(c)**. Hypermutants were defined as individuals with >99.5% percentile for singleton burden. **d-f,** Top panels represent the frequency of individuals within each singleton burden percentile category across age for deCODE **(d)**, WSI main **(e)**, and WSI Vanguard **(f)**. Bottom panels represent the association between singleton burden percentile category as the outcome (<95% percentile as reference) and age at blood sample collection as the predictor adjusted for age, sex, smoking, first four *peddy*-inferred genetic principal components in a logistic regression model. **g-i,** Proportion of nucleotide change (C>A, C>G, C>T, T>A, T>C, T>G) of singletons for each singleton burden percentile category for deCODE **(g)**, WSI main **(h)**, and WSI Vanguard **(i)**. **j-l,** Distribution of mean alternate allele frequency of singletons by singleton burden percentile category for deCODE **(j)**, WSI main **(k)**, and WSI Vanguard **(l)**. **m-o,** Distribution of sequencing depth at singleton site by singleton burden percentile category for deCODE **(m)**, WSI main **(n)**, and WSI Vanguard **(o)**.

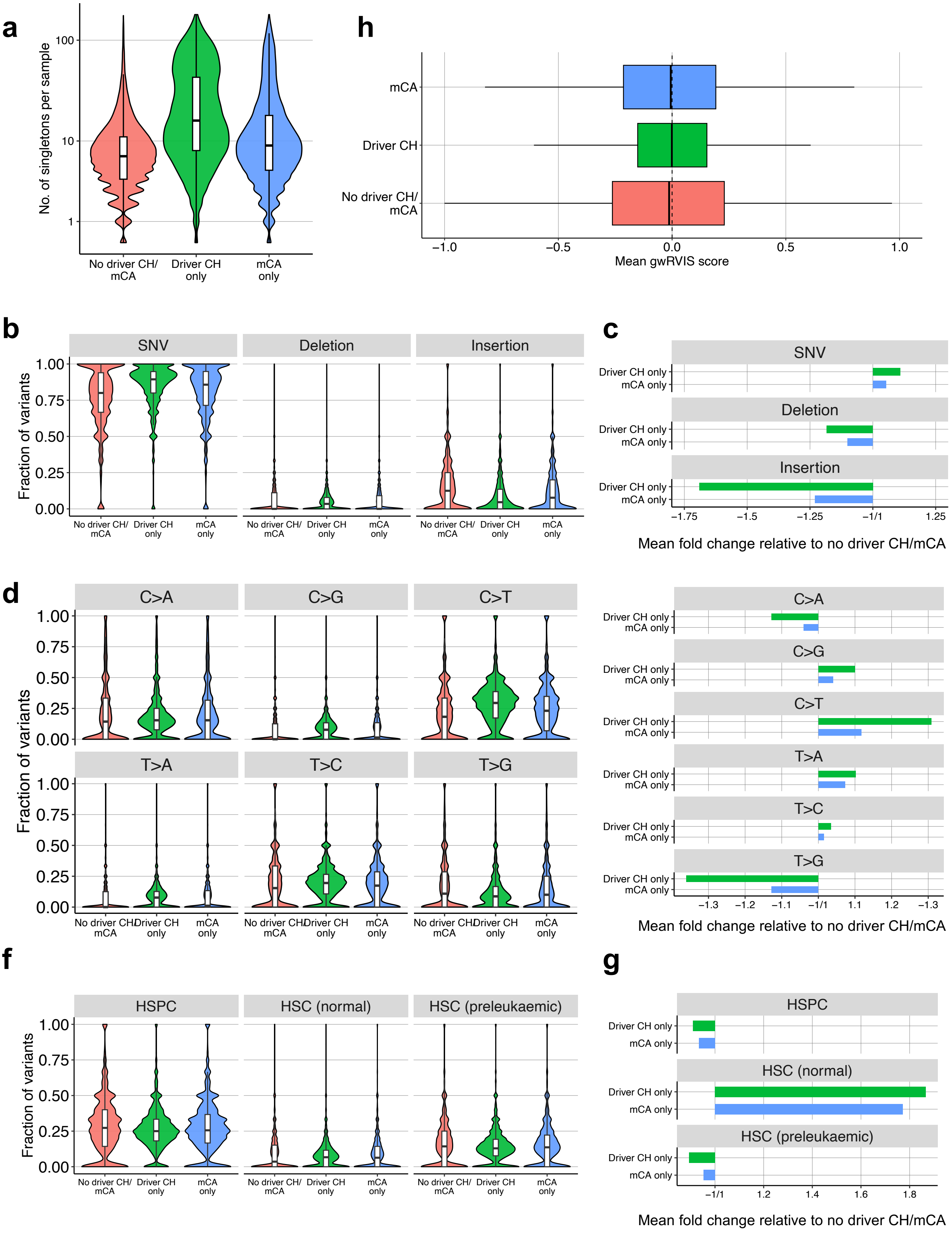

a
No. of singletons per sample
100
10
1
No driver CH/ mCA
Driver CH only
mCA only
h
mCA
Driver CH
No driver CH/ mCA
−1.0
−0.5
0.0
0.5
1.0
Mean gwRVIS score
b
SNV
Deletion
Insertion
Fraction of variants
1.00
0.75
0.50
0.25
0.00
c
Driver CH only
mCA only
−1.75
−1.5
−1.25
−1/1
1.25
Mean fold change relative to no driver CH/mCA
d
C>A
C>G
C>T
T>A
T>C
T>G
−1.3
−1.2
−1.1
−1/1
1.1
1.2
−1.3
f
HSPC
HSC (normal)
HSC (preleukaemic)
g
−1/1
1.2
1.4
1.6
1.8

**Supplementary Figure 4: Characteristics of genomic features of singletons included in the machine learning classifier to detect CH-UD.**

**a,** Number of singletons across non-CH individuals (no driver CH or mCA) and individuals with driver CH or mCA. **b,c,** Fraction of singletons that were single base substitution (single nucleotide variant; SNV), deletion or insertion **(b)**, and the enrichment or depletion of SNV, deletion or insertion in driver CH or mCA individuals relative to non-CH individuals **(c)**. **d,e,** Fraction of singletons by nucleotide change **(d)**, and the enrichment or depletion of nucleotide change in driver CH or mCA individuals relative to non-CH individuals **(e)**. **f,g,** Fraction of singletons located in the open chromatin region of HSPCs, normal HSCs, and pre-leukemic HSCs **(f)**, and the enrichment or depletion of these singletons in driver CH or mCA individuals relative to non-CH individuals **(g)**. HSPCs include normal and pre-leukaemic HSC and normal MPP, LMPP, CMP, GMP, MEP, and CLP. **h,** Distribution of gwRVIS score, an indicator of mutation tolerance at singleton site, across non-CH, driver CH, and mCA individuals. CLP, common lymphoid progenitors; CMP, common myeloid progenitors; GMP, Granulocyte-monocyte progenitors; HSC, haematopoietic stem cells; HSPC, haematopoietic stem and progenitor cells; LMPP, lymphoid-primed multipotent progenitors; MEP, Megakaryocyte–erythroid progenitors; MPP, Mouse multipotent progenitors; SNV, single nucleotide variants.

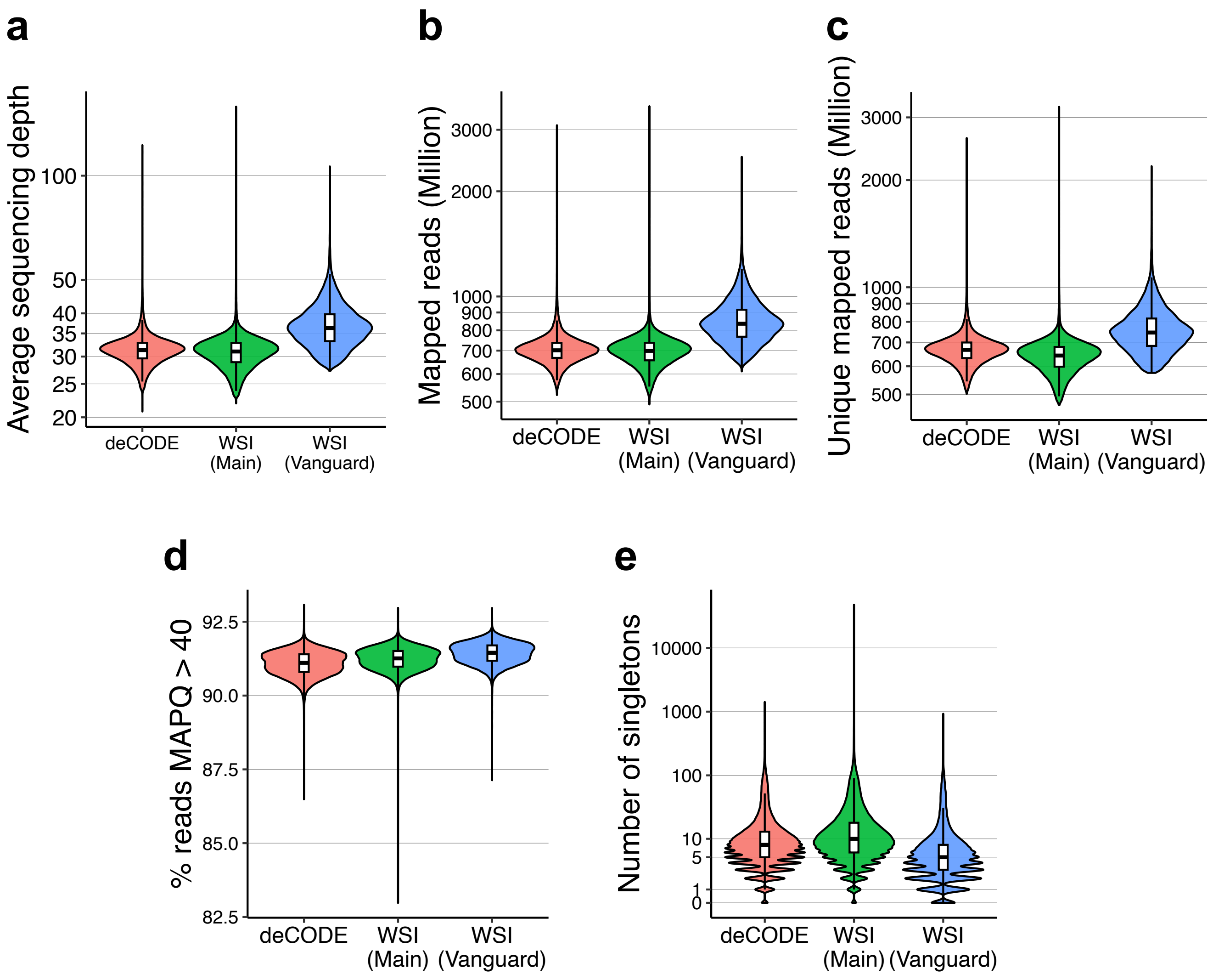
a
Average sequencing depth
100
50
40
35
30
25
20
deCODE
WSI (Main)
WSI (Vanguard)
b
Mapped reads (Million)
3000
2000
1000
900
800
700
600
500
deCODE
WSI (Main)
WSI (Vanguard)
c
Unique mapped reads (Million)
3000
2000
1000
900
800
700
600
500
deCODE
WSI (Main)
WSI (Vanguard)
d
% reads MAPQ > 40
92.5
90.0
87.5
85.0
82.5
deCODE
WSI (Main)
WSI (Vanguard)
e
Number of singletons
10000
1000
100
10
5
1
0
deCODE
WSI (Main)
WSI (Vanguard)

**Supplementary Figure 5: Sequencing metrices by whole-genome sequencing batch.**

**a-e,** Average sequencing depth **(a)**, number of mapped reads **(b)**, number of unique mapped reads **(c)**, percentage of reads with mapping quality score >40 **(d)**, and number of singletons detected **(e)** across deCODE, WSI main, and WSI Vanguard. MAPQ, mapping quality.

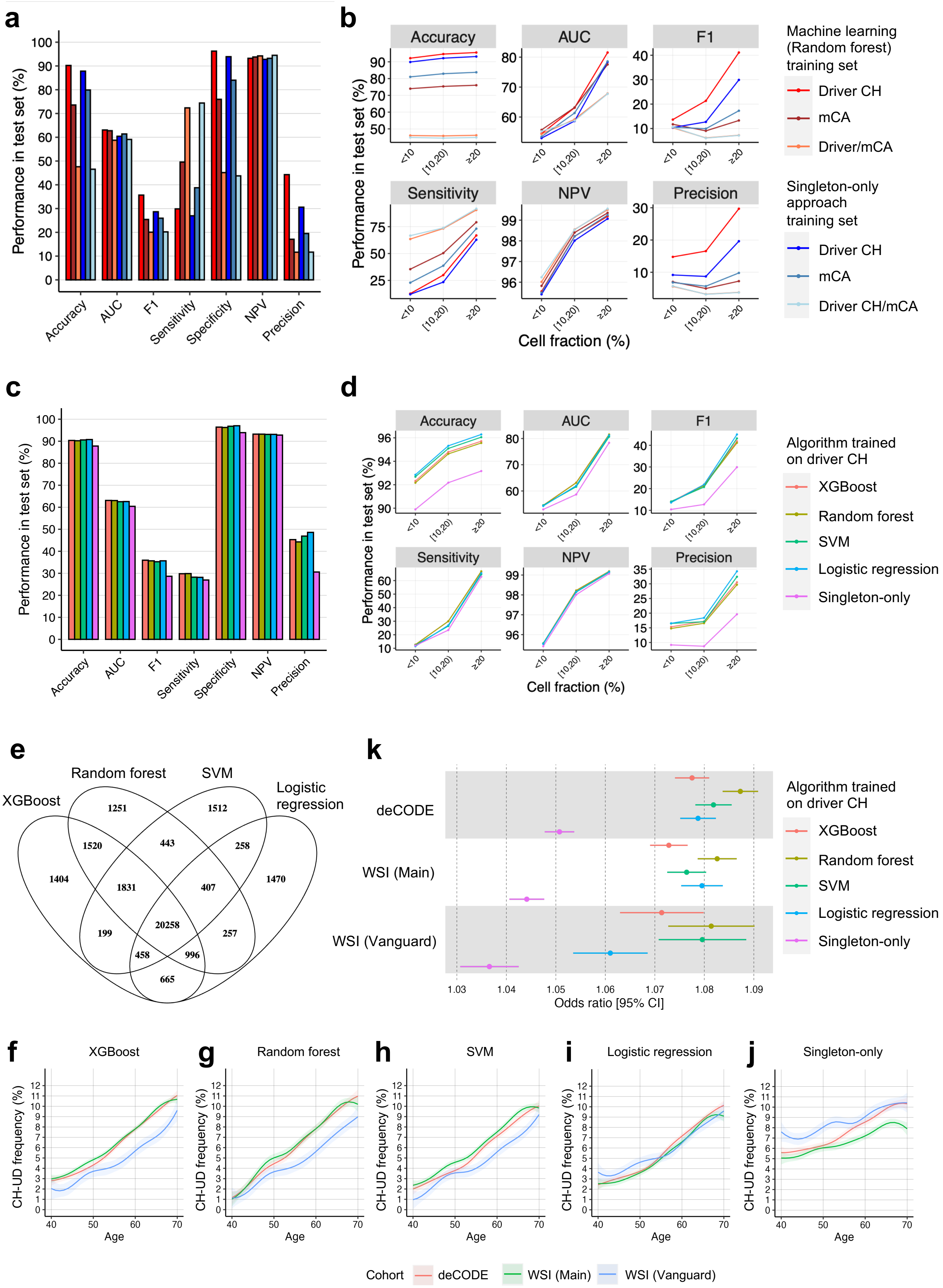
a
b
c
d
e
k
f
g
h
i
j
Performance in test set (%)
Accuracy
AUC
F1
Sensitivity
Specificity
NPV
Precision
Cell fraction (%)
<10
[10,20)
≥20
Machine learning (Random forest) training set
Driver CH
mCA
Driver/mCA
Singleton-only approach training set
Driver CH
mCA
Driver CH/mCA
Algorithm trained on driver CH
XGBoost
Random forest
SVM
Logistic regression
Singleton-only
XGBoost
Random forest
SVM
Logistic regression
1251
1512
1520
443
258
1404
1831
407
1470
20258
199
257
458
996
665
deCODE
WSI (Main)
WSI (Vanguard)
Odds ratio [95% CI]
CH-UD frequency (%)
Age
Cohort
deCODE
WSI (Main)
WSI (Vanguard)

**Supplementary Figure 6: Comparison of different training sets and different machine learning classifiers for the detection of CH-UD.**

**a,b,** Performance in the test set **(a)**, further stratified by the cell fraction of the CH clones **(b)**, when random forest was trained on the various training sets, namely training set consisting of driver CH, mCA, or any CH (driver CH or mCA). **c,d,** Performance in the test set **(c)**, further stratified by the cell fraction of the CH clones **(d)**, when the various machine learning classifiers were trained on the training set consisting of driver CH. The classifiers trained were XGBoost, random forest, SVM, and logistic regression. Additionally, an approach based solely on the number of singletons to distinguished CH from non-CH was assessed here ('singleton-only'). **e,** Degree of overlapping and non-overlapping samples defined as CH-UD by the different classifiers. **f-j,** Frequency of individuals with CH-UD across different age at blood sample collection when cases was inferred using XGBoost **(f)**, random forest **(g)**, SVM **(h)**, logistic regression **(i)** or singleton-only approach **(j)**. **k,** Association between CH-UD as the outcome (non-CH as reference) and age as the main predictor adjusted for sex, smoking, and first four *peddy*-computed genetic principal components in a logistic regression model for the different classifiers and singleton-only approach. AUC, area under curve; NPV, negative predictive value; SVM, support vector machine.

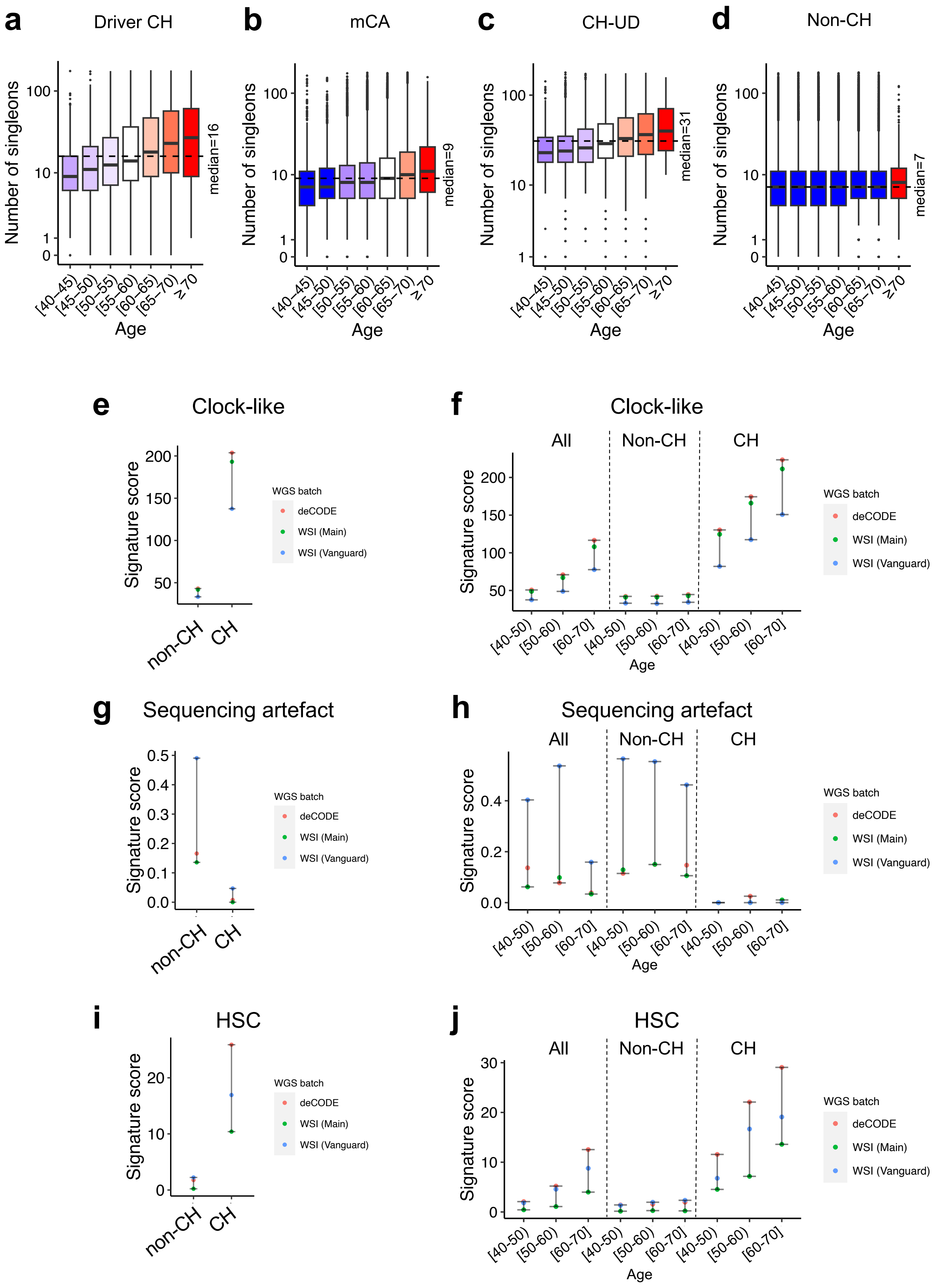
a
Driver CH
Number of singleons
median=16
[40–45) [45–50) [50–55) [55–60) [60–65) [65–70) ≥70
Age
b
mCA
median=9
c
CH-UD
median=31
d
Non-CH
median=7
e
Clock-like
Signature score
non-CH
CH
WGS batch
deCODE
WSI (Main)
WSI (Vanguard)
f
Clock-like
All
Non-CH
CH
[40-50) [50-60) [60-70]
g
Sequencing artefact
h
Sequencing artefact
i
HSC
j
HSC

**Supplementary Figure 7: Validation of detected singletons.**

**a-d**, Number of singletons by age group among driver CH **(a)**, mCA **(b)**, CH-UD **(c),** and individuals with no CH **(d)**. **i-j**, Clock-like, HSC, and sequencing artefact mutational signature enrichment by CH status **(e,g,i)** and further stratified by age **(f,h,j)**.

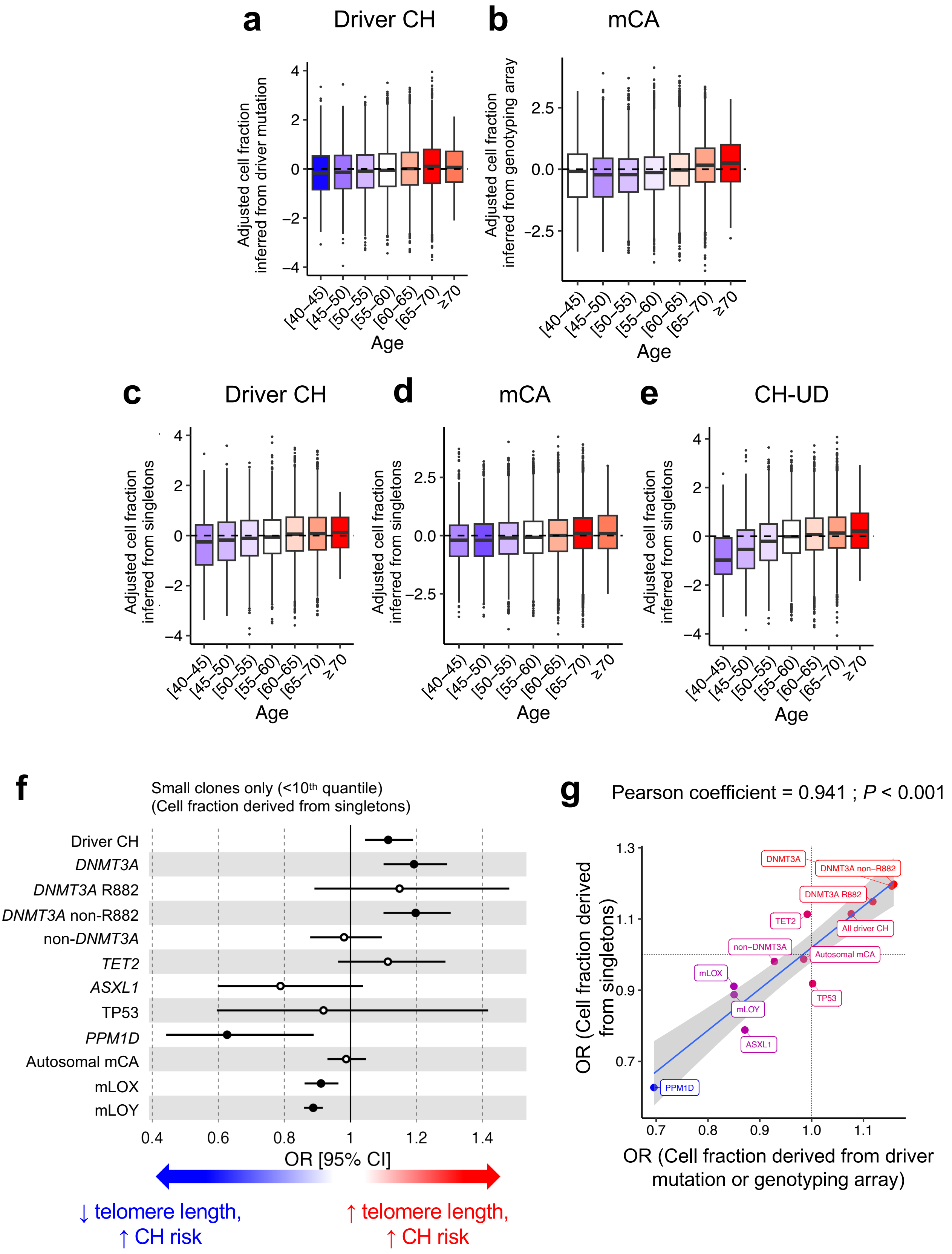

a
Driver CH
Adjusted cell fraction inferred from driver mutation
Age
b
mCA
Adjusted cell fraction inferred from genotyping array
Age
c
Driver CH
Adjusted cell fraction inferred from singletons
Age
d
mCA
Adjusted cell fraction inferred from singletons
Age
e
CH-UD
Adjusted cell fraction inferred from singletons
Age
[40–45) [45–50) [50–55) [55–60) [60–65) [65–70) ≥70
f
Small clones only (<10th quantile)
(Cell fraction derived from singletons)
Driver CH
DNMT3A
DNMT3A R882
DNMT3A non-R882
non-DNMT3A
TET2
ASXL1
TP53
PPM1D
Autosomal mCA
mLOX
mLOY
OR [95% CI]
↓ telomere length, ↑ CH risk
↑ telomere length, ↑ CH risk
g
Pearson coefficient = 0.941 ; P < 0.001
OR (Cell fraction derived from singletons)
OR (Cell fraction derived from driver mutation or genotyping array)
DNMT3A
DNMT3A non−R882
DNMT3A R882
TET2
All driver CH
non−DNMT3A
Autosomal mCA
mLOX
TP53
mLOY
ASXL1
PPM1D

**Supplementary Figure 8: Validation of singleton-derived cell fractions.**

**a,b,** Cell fraction as inferred from driver mutation (for driver CH; **a**) or from genotyping array (for mCA; **b**) across age. The adjusted cell fractions represent the residuals after regressing out the following variables from the cell fraction in a linear regression model: sex, smoking, and first four *peddy*-inferred genetic principal components. Inverse rank normalisation was further applied on residuals. **c-e,** Cell fraction as inferred from singletons across age among individuals with driver CH **(c)**, mCA **(d)**, or CH-UD **(e)**. The adjusted singleton-derived cell fractions represent the residuals after regressing out the following variables from the cell fraction in a linear regression model: coverage at singleton site, sex, smoking, and first four *peddy*-inferred genetic principal components. Inverse rank normalisation was further applied on the residuals. **f,** Association between measured telomere length as predictor with driver CH or mCA status as outcome (non-CH as reference) modelled using logistic regression. Only CH individuals with small clones, defined as <$10^{th}$ quantile of the singleton-derived cell fraction, were included in this analysis. **g,** The odds ratios on the y-axis were from **(f)**, and the odds ratios on the x-axis were derived in the same way as in **(f)**, but the small clones were defined as cell fraction <10% as inferred from driver mutations (for driver CH) or from genotyping array (for mCA). OR, odds ratio.

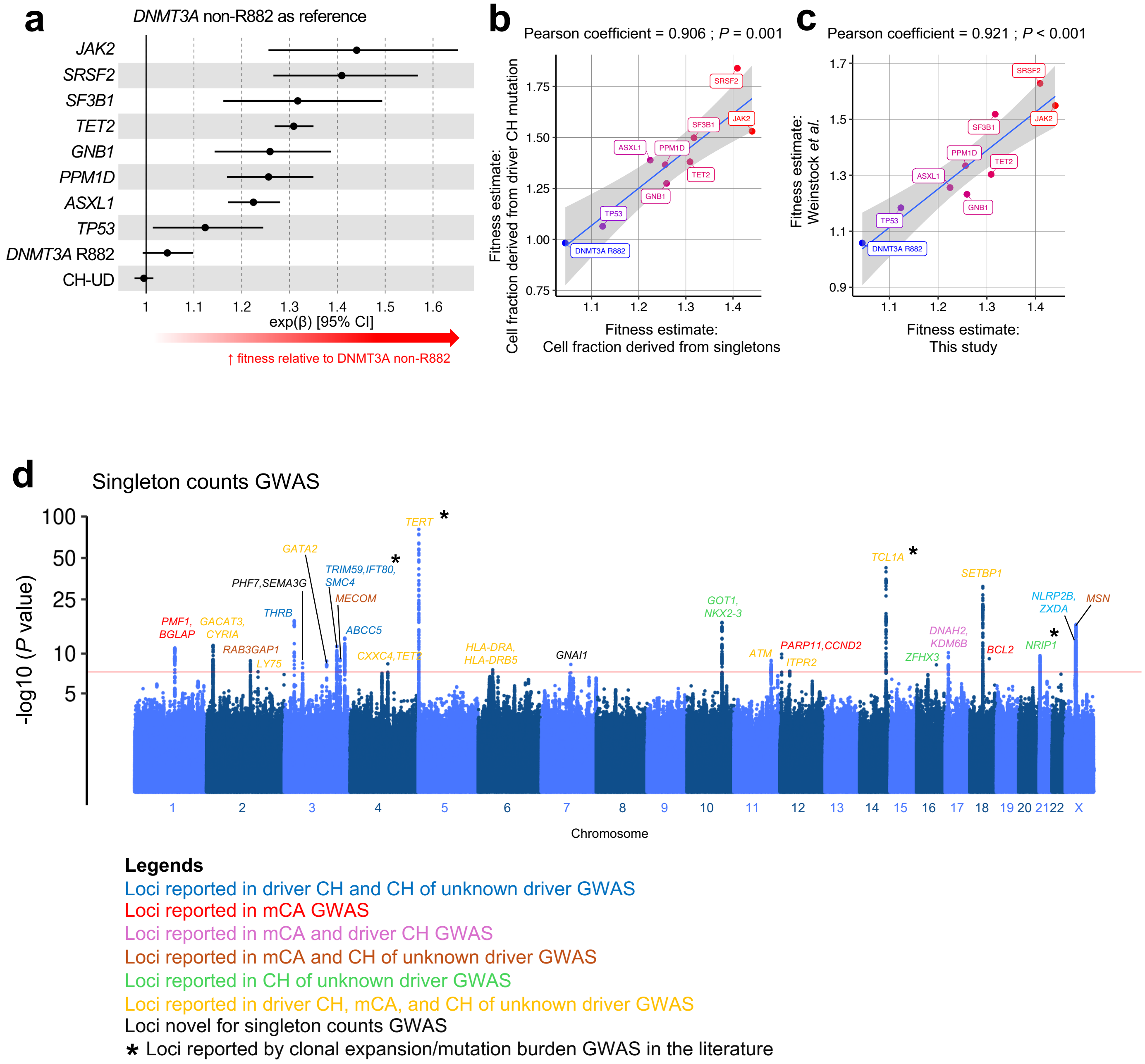
a
DNMT3A non-R882 as reference
JAK2
SRSF2
SF3B1
TET2
GNB1
PPM1D
ASXL1
TP53
DNMT3A R882
CH-UD
1 1.1 1.2 1.3 1.4 1.5 1.6
exp(β) [95% CI]
↑ fitness relative to DNMT3A non-R882
b
Pearson coefficient = 0.906 ; P = 0.001
Fitness estimate: Cell fraction derived from driver CH mutation
Fitness estimate: Cell fraction derived from singletons
c
Pearson coefficient = 0.921 ; P < 0.001
Fitness estimate: Weinstock et al.
Fitness estimate: This study
d
Singleton counts GWAS
-log10 (P value)
Chromosome
PMF1, BGLAP
GACAT3, CYRIA
RAB3GAP1
LY75
THRB
PHF7,SEMA3G
GATA2
TRIM59,IFT80, SMC4
MECOM
ABCC5
CXXC4,TET2
TERT
HLA-DRA, HLA-DRB5
GNAI1
GOT1, NKX2-3
ATM
PARP11,CCND2
ITPR2
TCL1A
ZFHX3
DNAH2, KDM6B
SETBP1
BCL2
NRIP1
NLRP2B, ZXDA
MSN
Legends
Loci reported in driver CH and CH of unknown driver GWAS
Loci reported in mCA GWAS
Loci reported in mCA and driver CH GWAS
Loci reported in mCA and CH of unknown driver GWAS
Loci reported in CH of unknown driver GWAS
Loci reported in driver CH, mCA, and CH of unknown driver GWAS
Loci novel for singleton counts GWAS
* Loci reported by clonal expansion/mutation burden GWAS in the literature

**Supplementary Figure 9: Validation of singletons and singleton-derived cell fractions using PACER.**

**a,** Fitness estimates of driver CH, mCA, and CH-UD inferred from negative binomial generalised linear model with singleton burden as the outcome and CH as the predictor (non-DNMT3A R882 individuals as reference) with adjusted singleton-derived cell fraction as covariate. The adjusted singleton-derived cell fractions were the residuals after regressing out the following variables from the cell fraction in a linear regression model: sequencing depth at singleton site, age, sex, smoking, and first four *peddy*-inferred genetic principal components. **b,** The fitness estimates on the x-axis were from **(a)**, and the fitness estimates on the y-axis were derived in the same way as in **(a)**, but the cell fraction was inferred from the driver mutations. **c,** The fitness estimates on the x-axis were from **(a)**, and the fitness estimates on the y-axis were derived from the published PACER study (Weinstock *et al.*, 2023). In the published PACER study, the cell fraction was inferred from the driver mutations. **d,** GWAS of singleton burden. *P* values were derived from linear regression implemented by SAIGE with singleton burden as the outcome with age, sex, smoking, *peddy*-inferred fraction of European probability, first ten genetic principal components, and adjusted singleton-derived cell fraction as co-variates. The adjusted singleton-derived cell fractions were the residuals after regressing out sequencing depth at singleton site in a linear regression model. Only individuals with CH (driver CH, mCA, and CH-UD) were included for analysis here. GWAS, genome-wide association study.

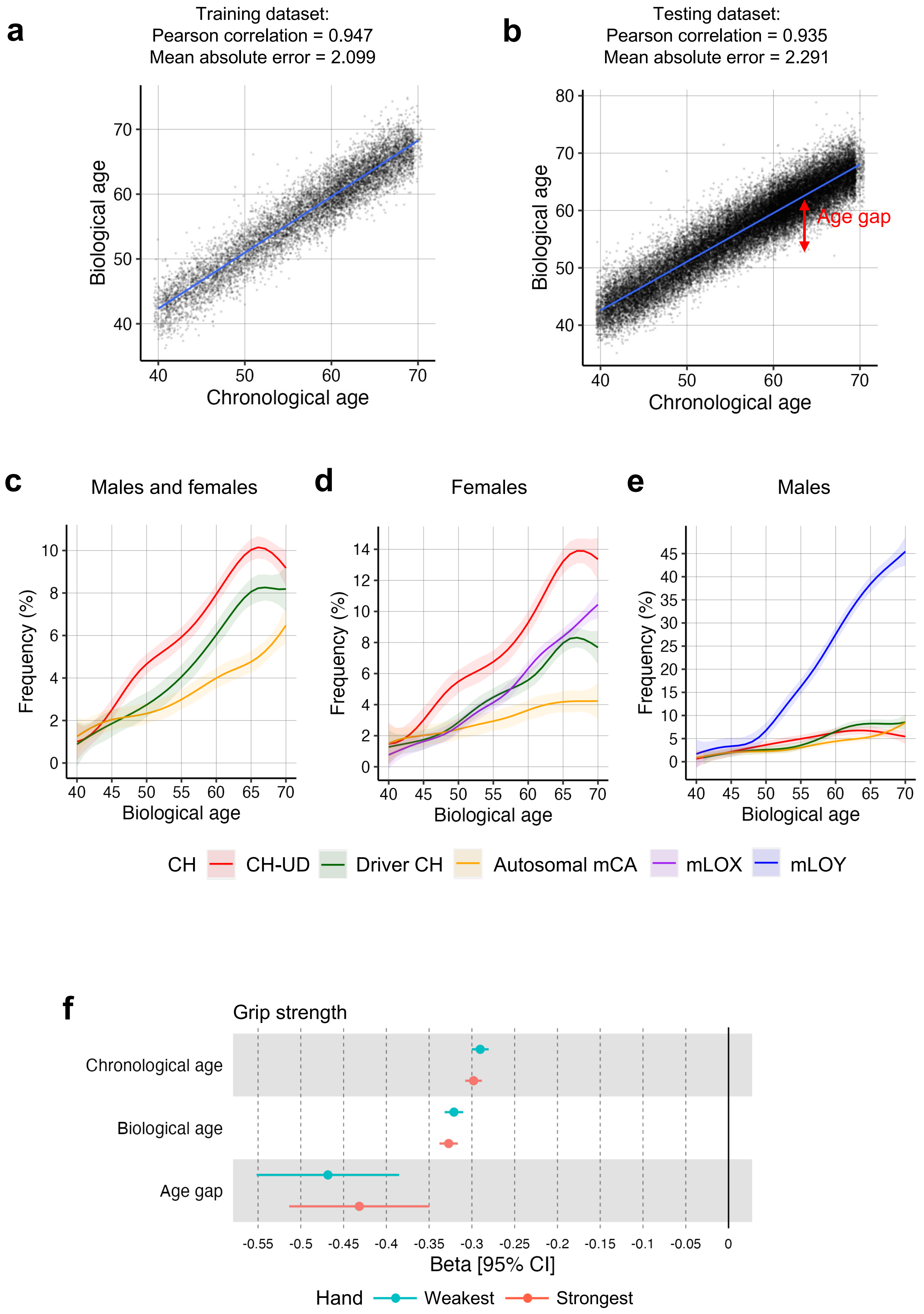

a
Training dataset:
Pearson correlation = 0.947
Mean absolute error = 2.099
Biological age
Chronological age
b
Testing dataset:
Pearson correlation = 0.935
Mean absolute error = 2.291
Age gap
Biological age
Chronological age
c
Males and females
d
Females
e
Males
Frequency (%)
Biological age
CH
CH-UD
Driver CH
Autosomal mCA
mLOX
mLOY
f
Grip strength
Chronological age
Biological age
Age gap
Beta [95% CI]
Hand
Weakest
Strongest

**Supplementary Figure 10: Validation of proteomic-based quantification of biological age and age gap.**

**a,b,** Correlation between biological age and chronological age among individuals included in the training set **(a)** or testing set **(b).** Age gap indicated as the difference between chronological versus biological age, specifically as the residual in the linear regression with chronological age as the predictor and biological age as the outcome. **c-e,** Frequency by biological age for CH-UD, driver CH, autosomal mCA, mLOX, and mLOY among all samples **(c)**, females only **(d)**, and males only **(e)**. **f,** Association between hand grip strength with age and age gap. Beta coefficient derived from linear regression with grip strength as the outcome and age or age gap as the main predictor with sex, Olink batch, time between measurement and sampling plasma protein, and first four *peddy*-inferred genetic principal components as co-variates.

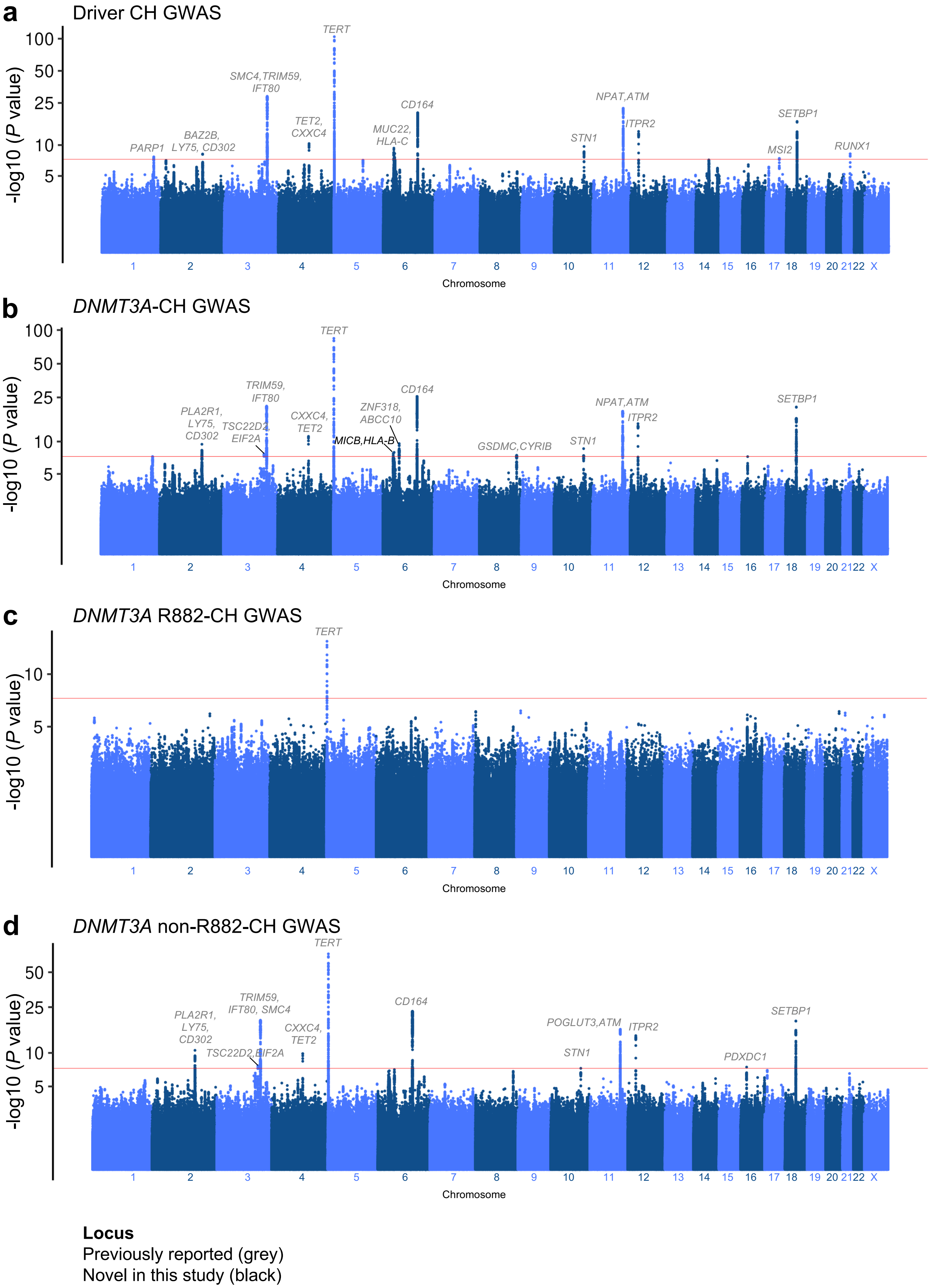
a
Driver CH GWAS
-log10 (P value)
100
50
25
10
5
TERT
SMC4,TRIM59, IFT80
TET2, CXXC4
BAZ2B, LY75, CD302
PARP1
MUC22, HLA-C
CD164
STN1
NPAT,ATM
ITPR2
SETBP1
MSI2
RUNX1
1 2 3 4 5 6 7 8 9 10 11 12 13 14 15 16 17 18 19 20 21 22 X
Chromosome
b
DNMT3A-CH GWAS
-log10 (P value)
100
50
25
10
5
TERT
TRIM59, IFT80
PLA2R1, LY75, CD302
TSC22D2, EIF2A
CXXC4, TET2
ZNF318, ABCC10
MICB,HLA-B
CD164
GSDMC,CYRIB
STN1
NPAT,ATM
ITPR2
SETBP1
1 2 3 4 5 6 7 8 9 10 11 12 13 14 15 16 17 18 19 20 21 22 X
Chromosome
c
DNMT3A R882-CH GWAS
-log10 (P value)
10
5
TERT
1 2 3 4 5 6 7 8 9 10 11 12 13 14 15 16 17 18 19 20 21 22 X
Chromosome
d
DNMT3A non-R882-CH GWAS
-log10 (P value)
50
25
10
5
TERT
TRIM59, IFT80, SMC4
PLA2R1, LY75, CD302
TSC22D2,EIF2A
CXXC4, TET2
CD164
POGLUT3,ATM
STN1
ITPR2
PDXDC1
SETBP1
1 2 3 4 5 6 7 8 9 10 11 12 13 14 15 16 17 18 19 20 21 22 X
Chromosome
Locus
Previously reported (grey)
Novel in this study (black)

**Supplementary Figure 11: Manhattan plots representing the common germline variants included for driver CH, *DNMT3A*-, *DNMT3A* R882-, and *DNMT3A* non-R882-CH GWAS.**

*P* values on the y-axis were derived from METAL software. GWAS was ran on each WGS batch separately using Firth logistic regression implemented using SAIGE software, and then meta-analysed using METAL. For SAIGE, we specified age, sex, smoking, *peddy*-inferred European probability, and first ten genetic principal components as co-variates. Previously reported loci indicated in grey while loci novel to this study indicated in black.

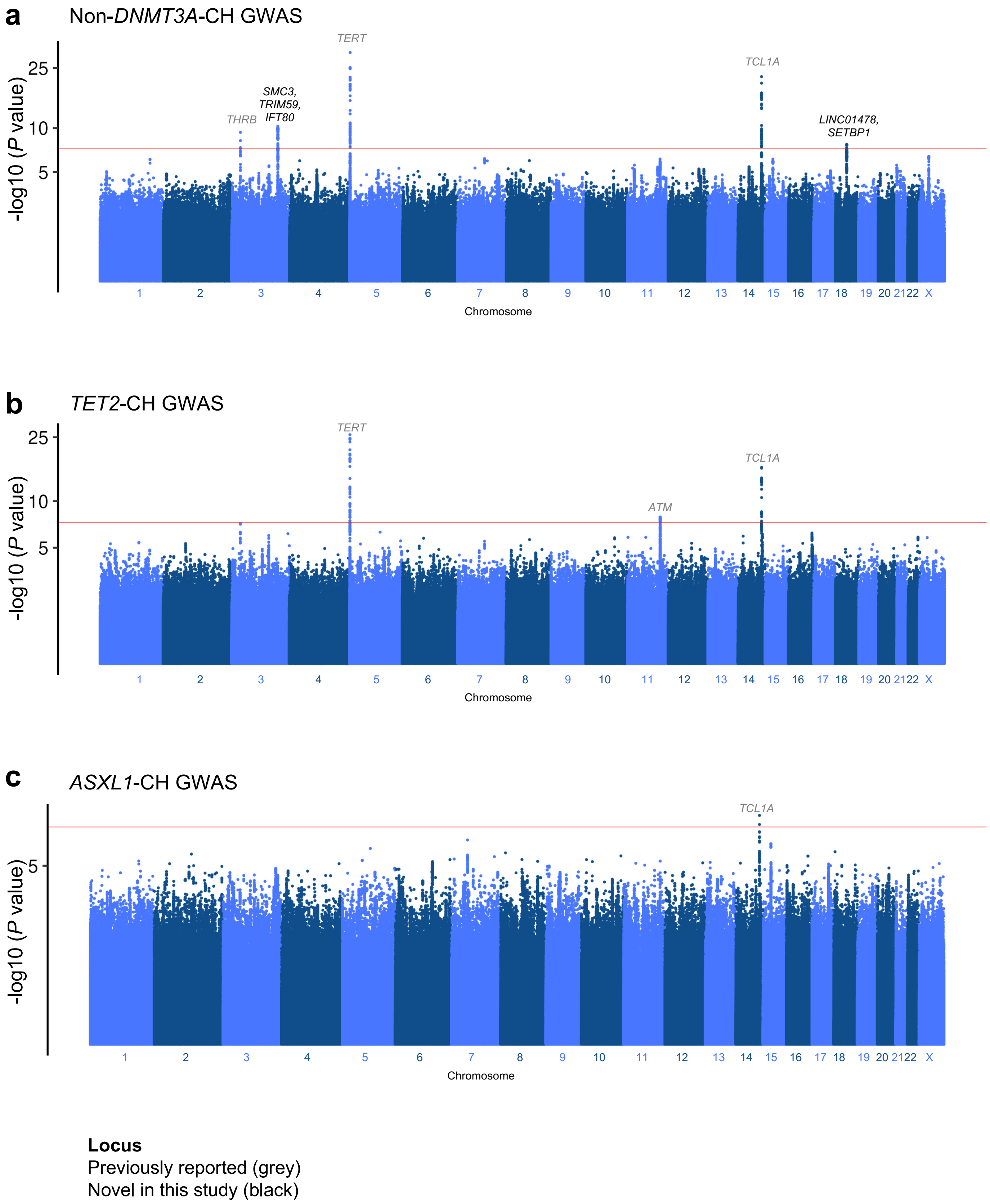
a
Non-DNMT3A-CH GWAS
-log10 (P value)
25
10
5
TERT
TCL1A
SMC3, TRIM59, IFT80
THRB
LINC01478, SETBP1
1 2 3 4 5 6 7 8 9 10 11 12 13 14 15 16 17 18 19 20 21 22 X
Chromosome
b
TET2-CH GWAS
-log10 (P value)
25
10
5
TERT
TCL1A
ATM
1 2 3 4 5 6 7 8 9 10 11 12 13 14 15 16 17 18 19 20 21 22 X
Chromosome
c
ASXL1-CH GWAS
-log10 (P value)
5
TCL1A
1 2 3 4 5 6 7 8 9 10 11 12 13 14 15 16 17 18 19 20 21 22 X
Chromosome
Locus
Previously reported (grey)
Novel in this study (black)

**Supplementary Figure 12: Manhattan plots representing the common germline variants included for non-*DNMT3A*-, *TET2*-, and *ASXL1*-CH GWAS.**

*P* values on the y-axis were derived from METAL software. GWAS was ran on each WGS batch separately using Firth logistic regression implemented using SAIGE software, and then meta-analysed using METAL. For SAIGE, we specified age, sex, smoking, *peddy*-inferred European probability, and first ten genetic principal components as co-variates. Previously reported loci indicated in grey while loci novel to this study indicated in black.

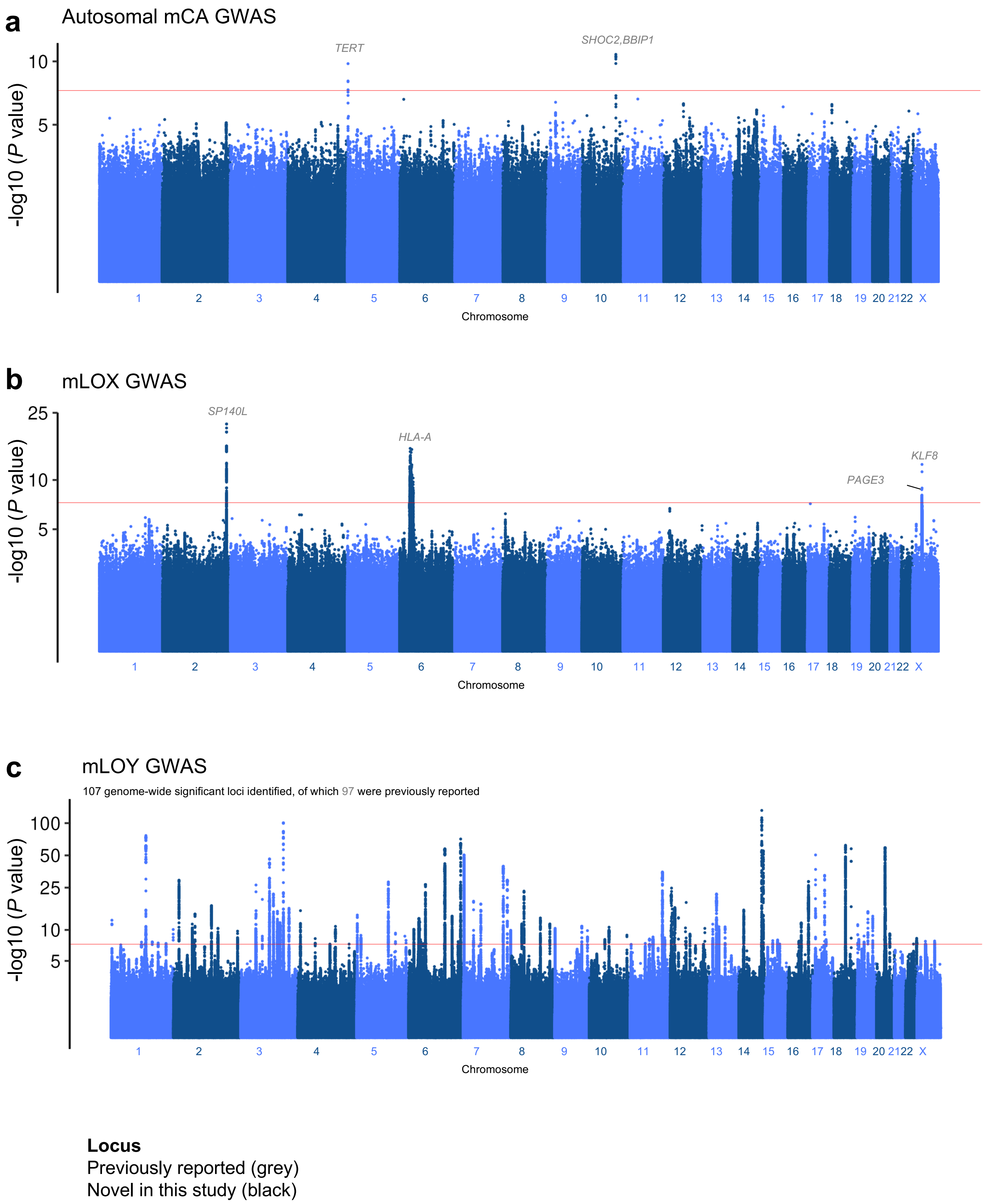
a
Autosomal mCA GWAS
TERT
SHOC2,BBIP1
-log10 (P value)
10
5
1 2 3 4 5 6 7 8 9 10 11 12 13 14 15 16 17 18 19 20 21 22 X
Chromosome
b
mLOX GWAS
SP140L
HLA-A
KLF8
PAGE3
25
10
5
Chromosome
c
mLOY GWAS
107 genome-wide significant loci identified, of which 97 were previously reported
100
50
25
10
5
Chromosome
Locus
Previously reported (grey)
Novel in this study (black)

**Supplementary Figure 13: Manhattan plots representing the common germline variants included for autosomal mCA, mLOX, and mLOY GWAS.**

*P* values on the y-axis were derived from METAL software. GWAS was ran on each WGS batch separately using Firth logistic regression implemented using SAIGE software, and then meta-analysed using METAL. For SAIGE, we specified age, sex, smoking, *peddy*-inferred European probability, and first ten genetic principal components as co-variates. Previously reported loci indicated in grey while loci novel to this study indicated in black.

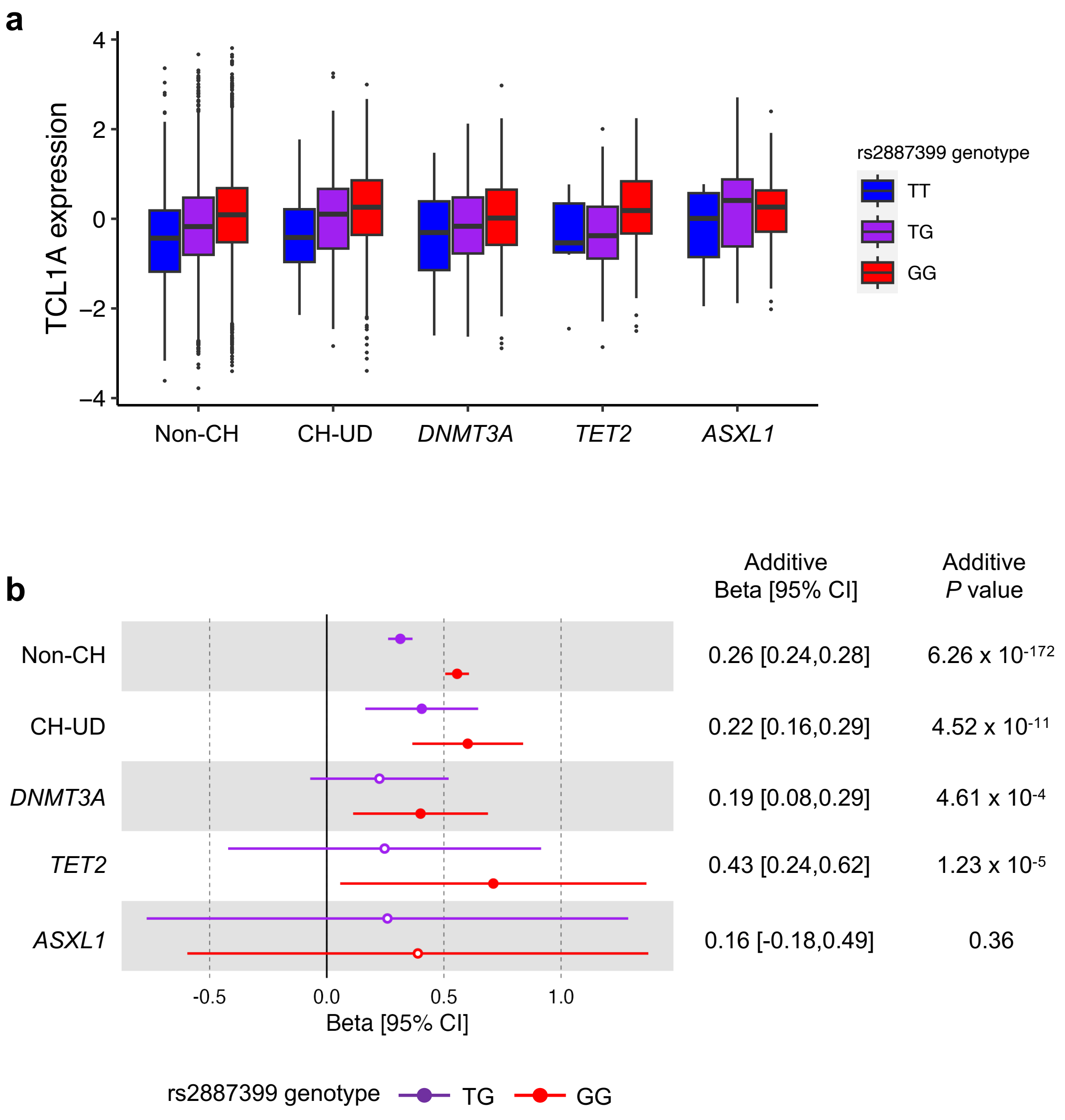
a
TCL1A expression
4
2
0
−2
−4
Non-CH
CH-UD
DNMT3A
TET2
ASXL1
rs2887399 genotype
TT
TG
GG
b
Additive Beta [95% CI]
Additive P value
Non-CH
0.26 [0.24,0.28]
6.26 x 10-172
CH-UD
0.22 [0.16,0.29]
4.52 x 10-11
DNMT3A
0.19 [0.08,0.29]
4.61 x 10-4
TET2
0.43 [0.24,0.62]
1.23 x 10-5
ASXL1
0.16 [-0.18,0.49]
0.36
-0.5
0.0
0.5
1.0
Beta [95% CI]
rs2887399 genotype
TG
GG

**Supplementary Figure 14: Association between rs2887399 genotype and TCL1A plasma protein level.**

**a,** TCL1A protein levels stratified by CH subtype and rs2887399 genotype. **b,** Association between rs2887399 TG or GG genotype (purple and red, respectively) with TCL1A protein levels for specific CH subtypes. Beta coefficients and *P* values were derived from linear regression with TCL1A protein level as the predictor and rs2887399 TG or GG genotype as the predictor (reference: TT genotype) adjusted for age, sex, smoking, and first four *peddy*-inferred principal components. rs2887399 genotype was also modelled in an additive fashion whereby TT, TG, and GG genotypes were treated as numeric values, i.e., 0, 1, 2 respectively. Lymphocyte counts have been regressed out from the TCL1A protein levels here. Specifically, the TCL1A protein levels here are the residuals computed using linear regression model with protein level as the outcome and lymphocyte count as the predictor. Solid circles represent $P < 0.05$ and hollow circles represent $P \geq 0.05$.

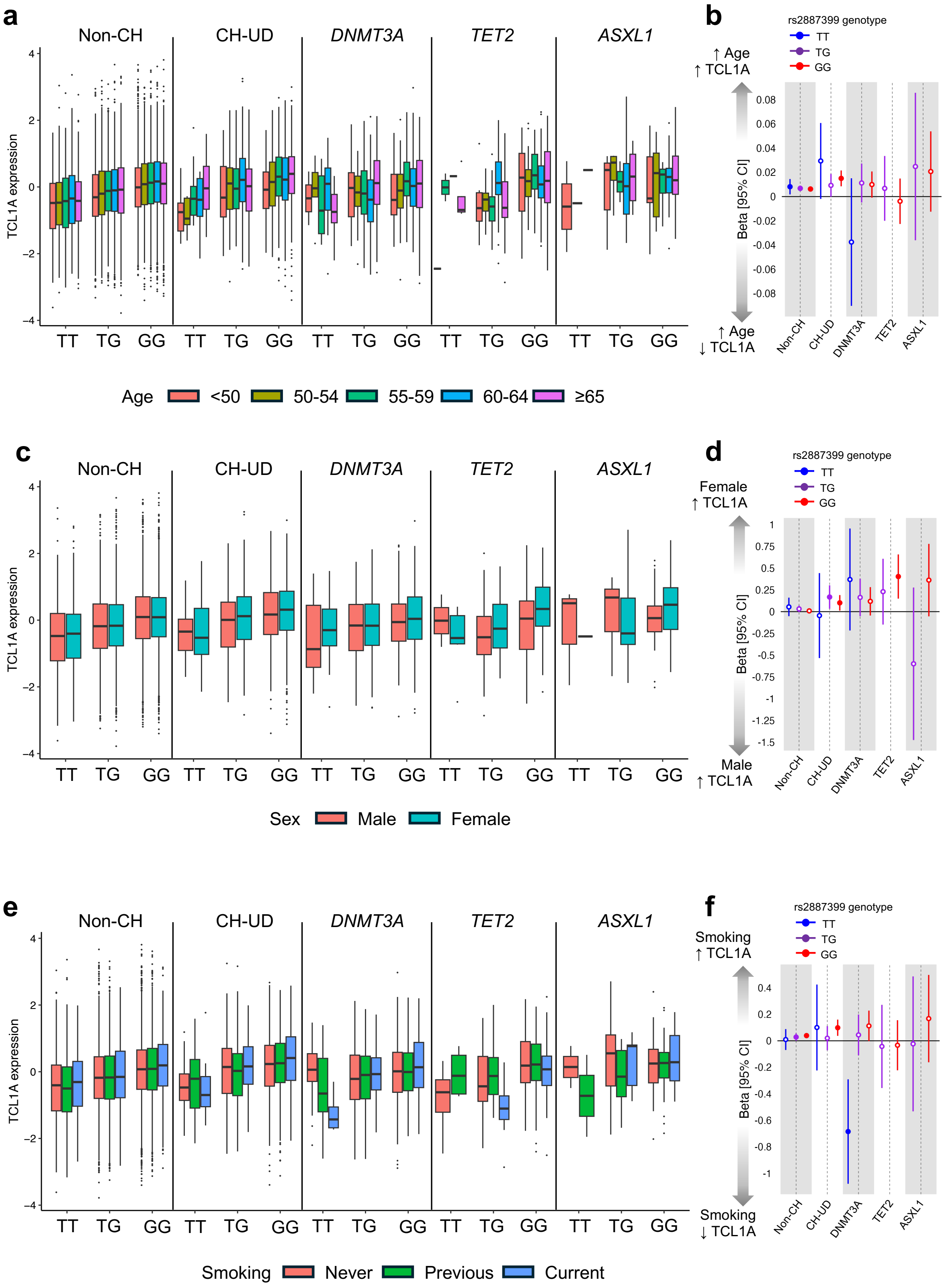
a
Non-CH
CH-UD
DNMT3A
TET2
ASXL1
TCL1A expression
TT
TG
GG
Age
<50
50-54
55-59
60-64
≥65
b
rs2887399 genotype
TT
TG
GG
↑ Age
↑ TCL1A
↑ Age
↓ TCL1A
Beta [95% CI]
Non-CH
CH-UD
DNMT3A
TET2
ASXL1
c
Sex
Male
Female
d
Female
↑ TCL1A
Male
↑ TCL1A
e
Smoking
Never
Previous
Current
f
Smoking
↑ TCL1A
Smoking
↓ TCL1A

**Supplementary Figure 15: Association between age, sex, and smoking with TCL1A plasma protein level.**

**a,** TCL1A protein levels stratified by age, CH subtype and rs2887399 genotype. **b,** For each rs2887399 genotype and each CH subtype, beta coefficients and *P* values were derived from linear regression with TCL1A protein level as the outcome and age as the quantitative predictor adjusted for sex, smoking status, and first four *peddy*-inferred principal components (PCs). **c,** TCL1A protein levels stratified by sex, CH subtype, and rs2887399 genotype. **d,** For each rs2887399 genotype and each CH subtype, beta coefficients and *P* values were derived from linear regression with TCL1A protein as the outcome and sex as the predictor (male as reference) adjusted for age, smoking status, and first four *peddy*-inferred PCs. **e,** TCL1A protein levels stratified by smoking status, CH subtype, and rs2887399 genotype. **f,** For each rs2887399 genotype and each CH subtype, beta coefficients and *P* values were derived from linear regression with TCL1A protein level as the outcome and smoking status as the predictor (never smoker as reference) adjusted for age, sex, and first four *peddy*-inferred PCs. Lymphocyte counts have been regressed out from the TCL1A protein levels here. Specifically, the TCL1A protein levels here are the residuals computed using linear regression model with protein level as the outcome and lymphocyte count as the predictor. Solid circles represent $P < 0.05$ and hollow circles represent $P \geq 0.05$.

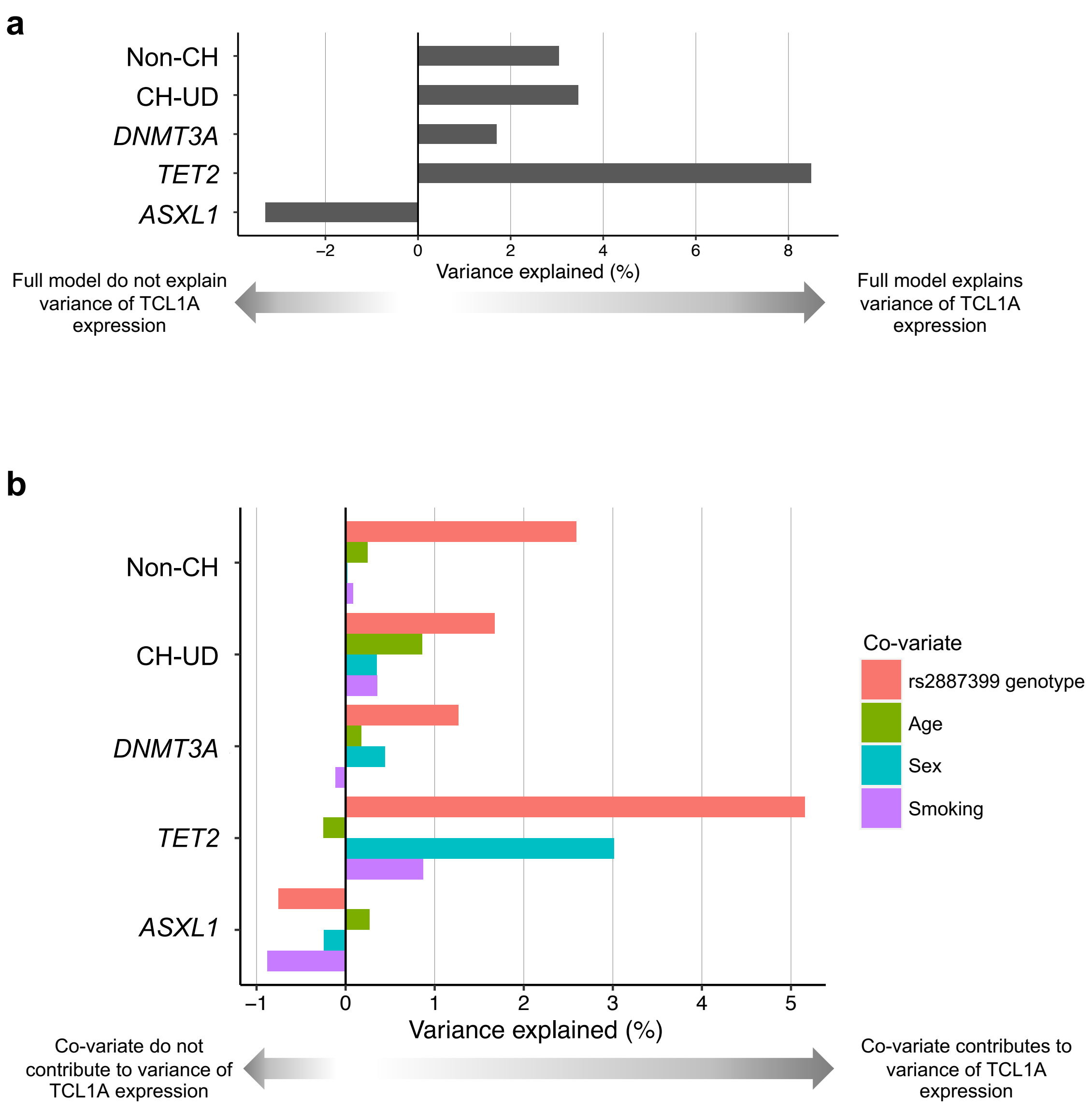
a
Non-CH
CH-UD
DNMT3A
TET2
ASXL1
−2
0
2
4
6
8
Variance explained (%)
Full model do not explain variance of TCL1A expression
Full model explains variance of TCL1A expression
b
Non-CH
CH-UD
DNMT3A
TET2
ASXL1
−1
0
1
2
3
4
5
Variance explained (%)
Co-variate
rs2887399 genotype
Age
Sex
Smoking
Co-variate do not contribute to variance of TCL1A expression
Co-variate contributes to variance of TCL1A expression

**Supplementary Figure 16: Proportion of variance of TCL1A protein levels explained by rs2887399 genotype, age, sex, and smoking.**

**a,** For each CH subtype, the proportion of variance of TCL1A protein levels explained by the full model. The model was a multivariate linear regression with TCL1A protein level as the outcome and rs2887399 genotype, age, sex, and smoking as the co-variates. **b,** For each CH subtype, the proportion of variance of TCL1A protein levels explained by rs2887399 genotype, age, sex, or smoking status. For each co-variate, a univariate linear regression was fitted with TCL1A protein level as the outcome and the corresponding co-variate as the predictor. The proportion of variance explained was derived from the adjusted r-squared multiplied by 100.

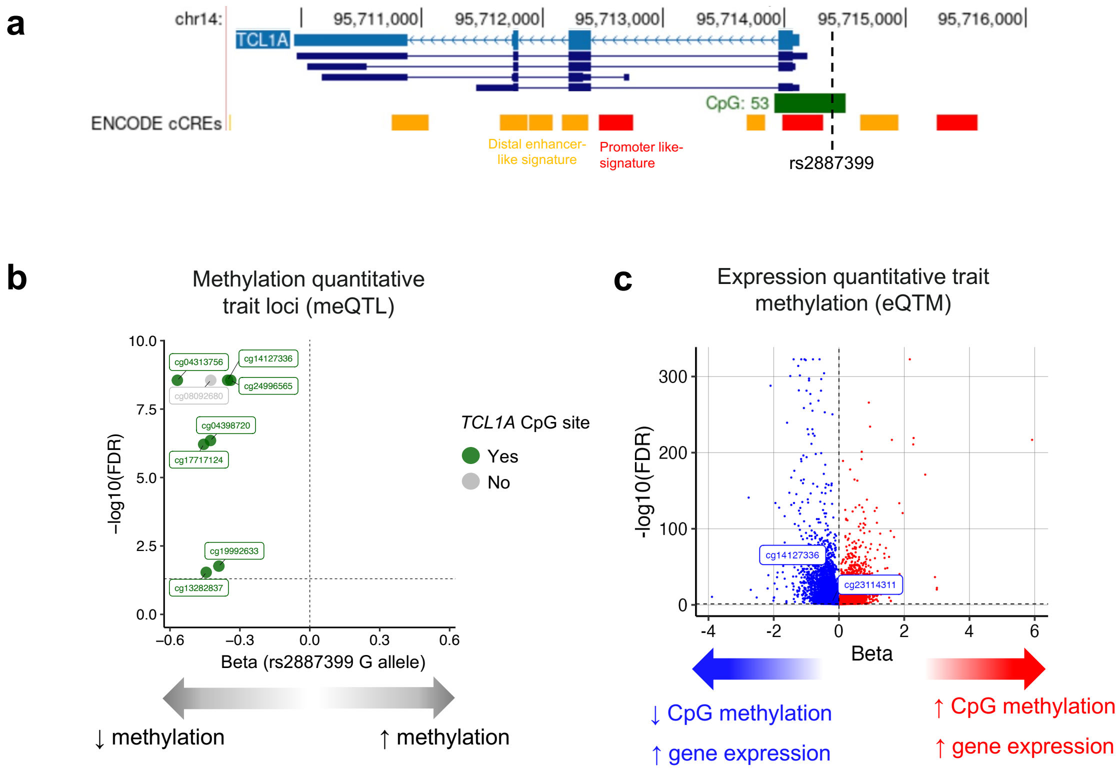
a
chr14:
95,711,000
95,712,000
95,713,000
95,714,000
95,715,000
95,716,000
TCL1A
CpG: 53
ENCODE cCREs
Distal enhancer-like signature
Promoter like-signature
rs2887399
b
Methylation quantitative trait loci (meQTL)
−log10(FDR)
Beta (rs2887399 G allele)
TCL1A CpG site
Yes
No
↓ methylation
↑ methylation
c
Expression quantitative trait methylation (eQTM)
-log10(FDR)
Beta
cg14127336
cg23114311
↓ CpG methylation
↑ gene expression
↑ CpG methylation
↑ gene expression

**Supplementary Figure 17: Association between rs2887399 genotype and *TCL1A* promoter methylation.**

**a,** UCSC Genome Browser view of the location, in hg38, of the *TCL1A* gene, CpG site, and ENCODE-annotated candidate cis-regulatory elements (cCREs). **b,** Significant methylation quantitative trait loci (mQTL) of rs2887399 G allele (FDR < 0.05). Summary statistics were retrieved from Villicaña *et al*. **c,** Expression quantitative trait methylation (eQTM) for 34,801 CpG sites. *TCL1A* CpG sites annotated. Summary statistics were retrieved from Hoang *et al*.

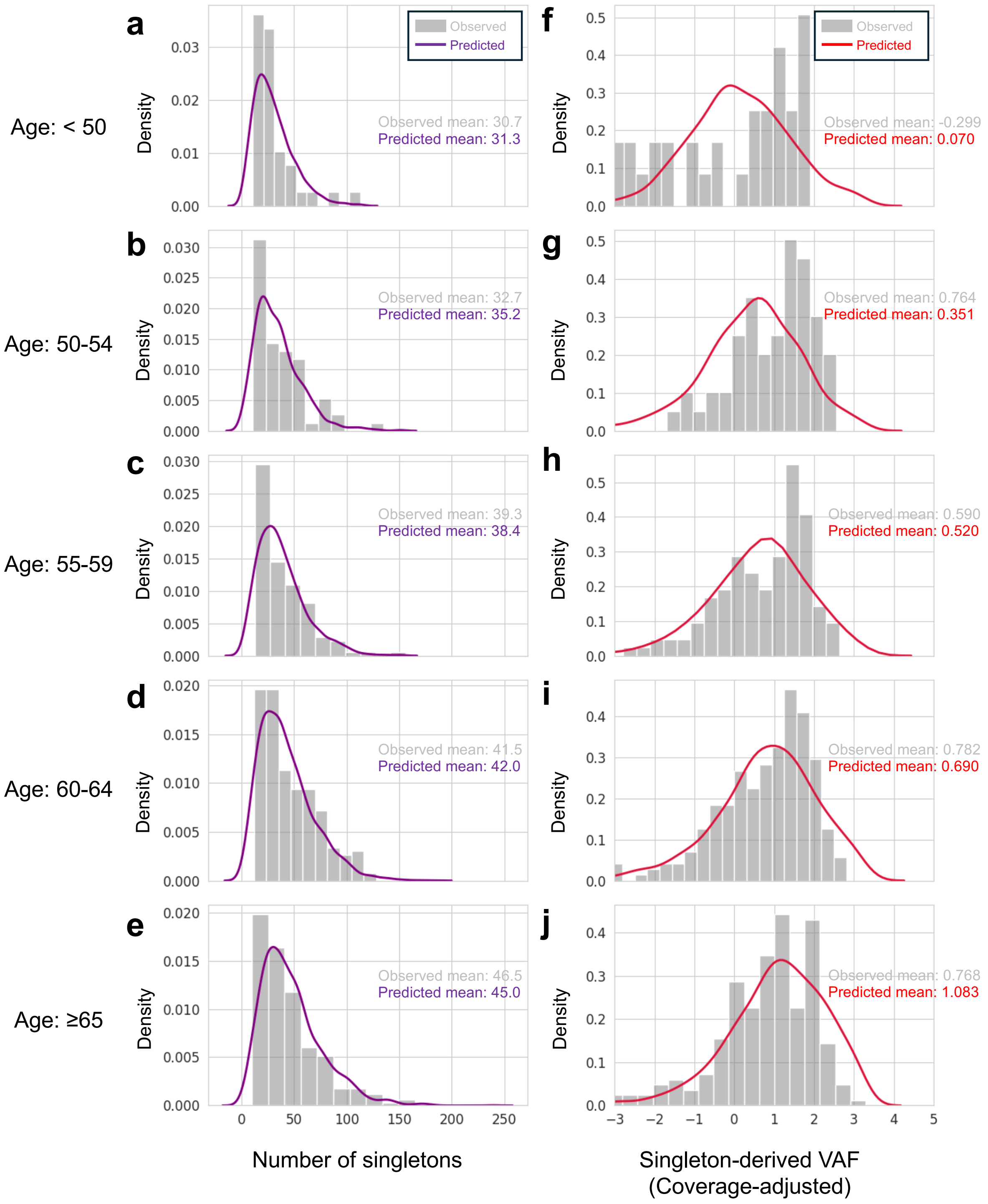

Age: < 50
Age: 50-54
Age: 55-59
Age: 60-64
Age: ≥65
a
b
c
d
e
f
g
h
i
j
Density
Observed
Predicted
Observed mean: 30.7
Predicted mean: 31.3
Observed mean: 32.7
Predicted mean: 35.2
Observed mean: 39.3
Predicted mean: 38.4
Observed mean: 41.5
Predicted mean: 42.0
Observed mean: 46.5
Predicted mean: 45.0
Observed mean: -0.299
Predicted mean: 0.070
Observed mean: 0.764
Predicted mean: 0.351
Observed mean: 0.590
Predicted mean: 0.520
Observed mean: 0.782
Predicted mean: 0.690
Observed mean: 0.768
Predicted mean: 1.083
Number of singletons
Singleton-derived VAF (Coverage-adjusted)

**Supplementary Figure 18: Age-stratified joint model fit to singleton burden.**

**a-j,** Observed data (grey bars) and model posterior predictive distributions (purple and red colored lines) for the study population across five age groups for the distribution of singleton burden **(a-e)** and singleton-derived variant allele frequency (VAF) with singleton coverage regressed out **(f-j)**. VAF was adjusted for the singleton coverage. Specifically, the VAF values here were the residuals computed using linear regression model with VAF as the outcome and singleton coverage as the predictor.

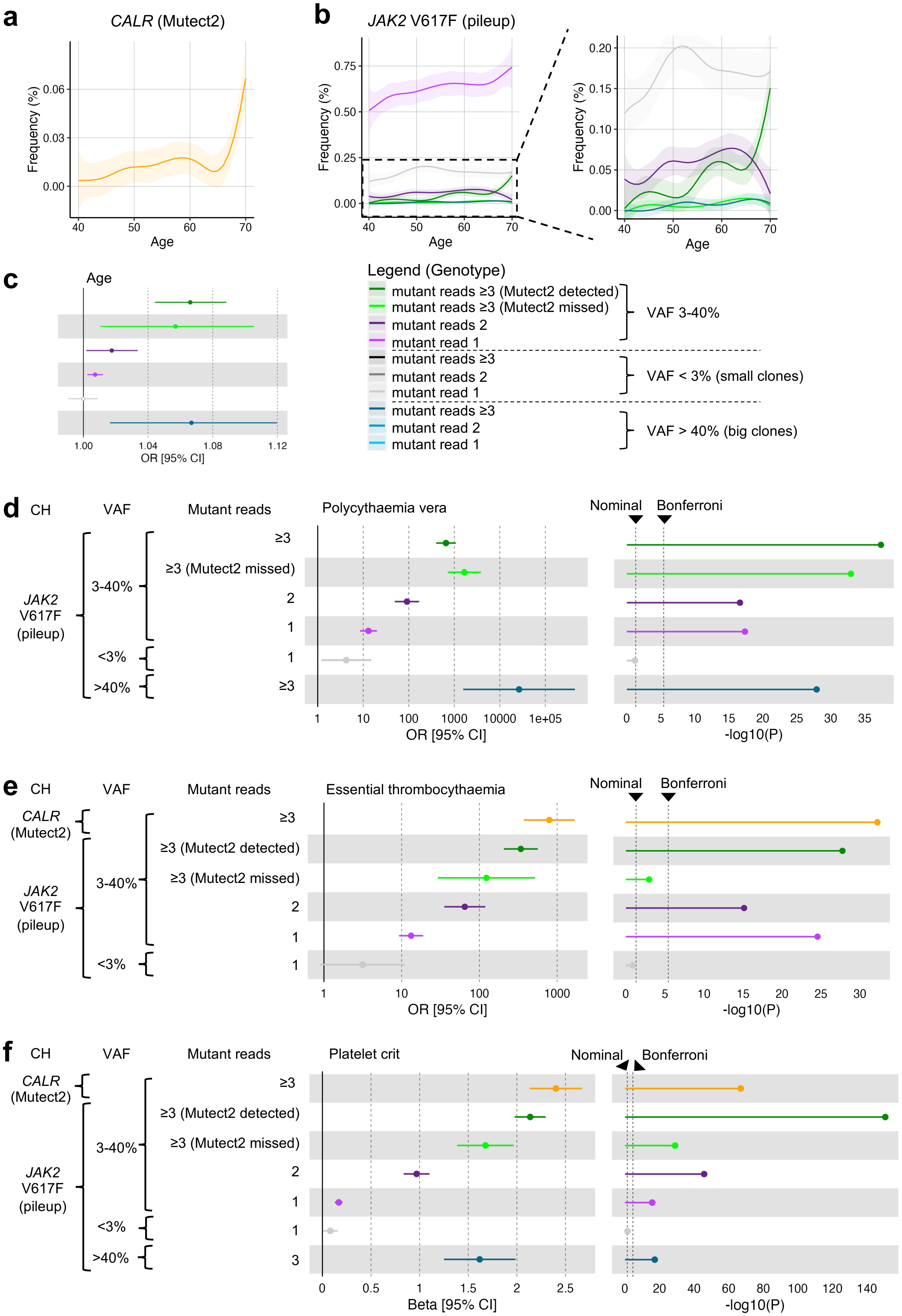
a
CALR (Mutect2)
Frequency (%)
Age
b
JAK2 V617F (pileup)
Frequency (%)
Age
c
Age
OR [95% CI]
Legend (Genotype)
mutant reads ≥3 (Mutect2 detected)
mutant reads ≥3 (Mutect2 missed)
mutant reads 2
mutant read 1
VAF 3-40%
mutant reads ≥3
mutant reads 2
mutant read 1
VAF < 3% (small clones)
mutant reads ≥3
mutant read 2
mutant read 1
VAF > 40% (big clones)
d
CH
VAF
Mutant reads
Polycythaemia vera
Nominal
Bonferroni
JAK2 V617F (pileup)
3-40%
<3%
>40%
≥3
≥3 (Mutect2 missed)
2
1
1
≥3
OR [95% CI]
-log10(P)
e
CH
VAF
Mutant reads
Essential thrombocythaemia
Nominal
Bonferroni
CALR (Mutect2)
JAK2 V617F (pileup)
3-40%
<3%
≥3
≥3 (Mutect2 detected)
≥3 (Mutect2 missed)
2
1
1
OR [95% CI]
-log10(P)
f
CH
VAF
Mutant reads
Platelet crit
Nominal
Bonferroni
CALR (Mutect2)
JAK2 V617F (pileup)
3-40%
<3%
>40%
≥3
≥3 (Mutect2 detected)
≥3 (Mutect2 missed)
2
1
1
3
Beta [95% CI]
-log10(P)

**Supplementary Figure 19: Detection and validation of Mutect2-detected *CALR* indels and pileup-detected *JAK2* V617F variants.**

**a-b,** Assessment of CH frequency by age for *CALR* **(a)** and *JAK2* V617F **(b)** mutation carriers. For *JAK2* V617F, we assessed several genotypes based on the number of reads supporting the mutant allele (1, 2, ≥3) and VAF (<3%, 3-40%, >40%). In our current study, *JAK2* mutations was originally detected using Mutect2 and required ≥3 mutant reads and 3-40% VAF. **c,** Further assessment of *JAK2* V617F genotypes using logistic regression with genotype as the outcome and age as the predictor adjusted for sex, smoking, and first four *peddy*-inferred genetic principal components (PCs). **d,** Association between *JAK2* V617F genotypes with polycythaemia vera (PV). Odds ratio was derived from Firth logistic regression with genotype as the predictor (non-CH as reference) and disease as the outcome adjusted for age, sex, smoking, waist circumferences, body mass index, and first four *peddy*-inferred genetic PCs. **e,** Association between *CALR* and *JAK2* V617F genotypes with essential thrombocythaemia (ET). Odds ratios were derived from Firth logistic regression with genotype as the predictor (non-CH as reference) and disease as the outcome adjusted for age, sex, smoking, waist circumferences, body mass index, and first four *peddy*-inferred genetic PCs. Bonferroni threshold for binary phenotypes was $P < 3.78 \times 10^{-6}$ while nominal threshold was defined with $P < 0.05$ **(d,e)**. **f,** Association between *CALR* and *JAK2* V617F genotypes with platelet crit. Beta coefficients were derived from linear regression with genotype as the predictor (non-CH as reference) and platelet crit as the outcome adjusted for age, sex, smoking, waist circumferences, body mass index, and first four *peddy*-inferred genetic PCs. Bonferroni threshold for quantitative phenotypes was $P < 2.97 \times 10^{-5}$ while nominal threshold was defined with $P < 0.05$. VAF, variant allele frequency.

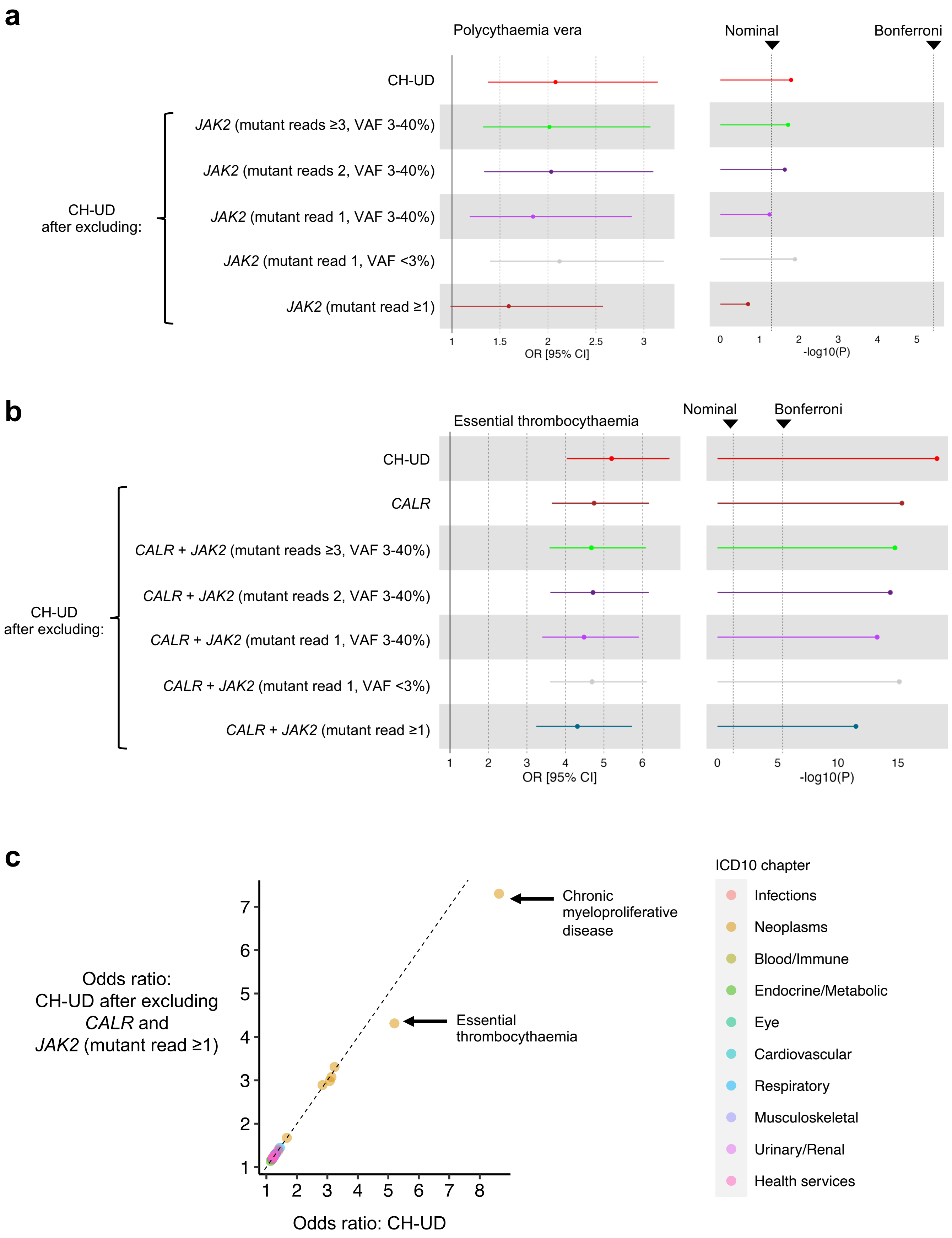

a
Polycythaemia vera
Nominal
Bonferroni
CH-UD
JAK2 (mutant reads ≥3, VAF 3-40%)
JAK2 (mutant reads 2, VAF 3-40%)
CH-UD after excluding:
JAK2 (mutant read 1, VAF 3-40%)
JAK2 (mutant read 1, VAF <3%)
JAK2 (mutant read ≥1)
1 1.5 2 2.5 3
OR [95% CI]
0 1 2 3 4 5
-log10(P)
b
Essential thrombocythaemia
Nominal
Bonferroni
CH-UD
CALR
CALR + JAK2 (mutant reads ≥3, VAF 3-40%)
CALR + JAK2 (mutant reads 2, VAF 3-40%)
CH-UD after excluding:
CALR + JAK2 (mutant read 1, VAF 3-40%)
CALR + JAK2 (mutant read 1, VAF <3%)
CALR + JAK2 (mutant read ≥1)
1 2 3 4 5 6
OR [95% CI]
0 5 10 15
-log10(P)
c
Odds ratio: CH-UD after excluding CALR and JAK2 (mutant read ≥1)
1 2 3 4 5 6 7
Chronic myeloproliferative disease
Essential thrombocythaemia
1 2 3 4 5 6 7 8
Odds ratio: CH-UD
ICD10 chapter
Infections
Neoplasms
Blood/Immune
Endocrine/Metabolic
Eye
Cardiovascular
Respiratory
Musculoskeletal
Urinary/Renal
Health services

**Supplementary Figure 20: Association between binary phenotypes with CH-UD before versus after excluding Mutect2-detected *CALR* indels and pileup-detected *JAK2* V617F variants.**

**a,** Association between polycythaemia vera (PV) with CH-UD before versus after excluding *CALR* indels and additional *JAK2* V617F variants. **b,** Association between essential thrombocythaemia (ET) with CH-UD before versus after excluding *CALR* indels and additional *JAK2* V617F variants. **c,** Comparison of summary statistic of selected phenotypes as shown in Figure 6a, stratified by ICD10 chapters, before versus after excluding *CALR* indels and additional *JAK2* V617F variants.. Odds ratios were derived from Firth logistic regression with CH-UD as the predictor (non-CH as reference) and phenotype as the outcome adjusted for age, sex, smoking, waist circumferences, body mass index, and first four *peddy*-inferred genetic principal components.

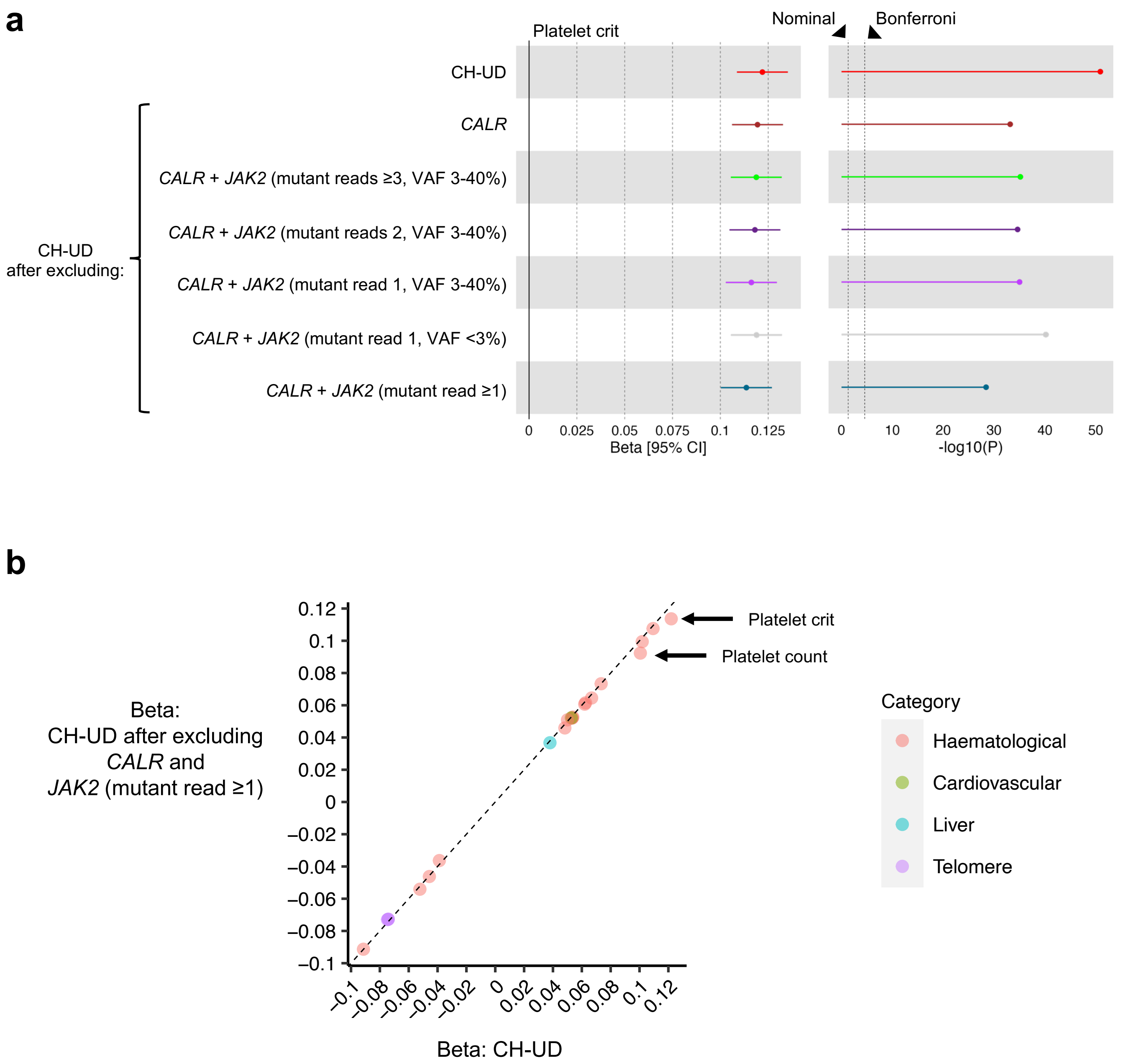
a
Platelet crit
Nominal
Bonferroni
CH-UD
CALR
CALR + JAK2 (mutant reads ≥3, VAF 3-40%)
CALR + JAK2 (mutant reads 2, VAF 3-40%)
CALR + JAK2 (mutant read 1, VAF 3-40%)
CALR + JAK2 (mutant read 1, VAF <3%)
CALR + JAK2 (mutant read ≥1)
CH-UD after excluding:
0 0.025 0.05 0.075 0.1 0.125
Beta [95% CI]
0 10 20 30 40 50
-log10(P)
b
Beta: CH-UD after excluding CALR and JAK2 (mutant read ≥1)
Beta: CH-UD
Platelet crit
Platelet count
Category
Haematological
Cardiovascular
Liver
Telomere
0.12 0.1 0.08 0.06 0.04 0.02 0 −0.02 −0.04 −0.06 −0.08 −0.1
−0.1 −0.08 −0.06 −0.04 −0.02 0 0.02 0.04 0.06 0.08 0.1 0.12

**Supplementary Figure 21: Association between quantitative phenotypes with CH-UD before versus after excluding Mutect2-detected *CALR* indels and pileup-detected *JAK2* V617F variants.**

**a,** Association between platelet crit with CH-UD before versus after excluding *CALR* variants and additional *JAK2* V617F. **b,** Comparison of summary statistic of selected phenotypes as shown in Figure 6b before versus after excluding *CALR* indels and additional *JAK2* V617F. Beta coefficients were derived from linear regression with CH-UD as the predictor (non-CH as reference) and phenotype as the outcome adjusted for age, sex, smoking, waist circumferences, body mass index, and first four *peddy*-inferred genetic principal components.

**a**

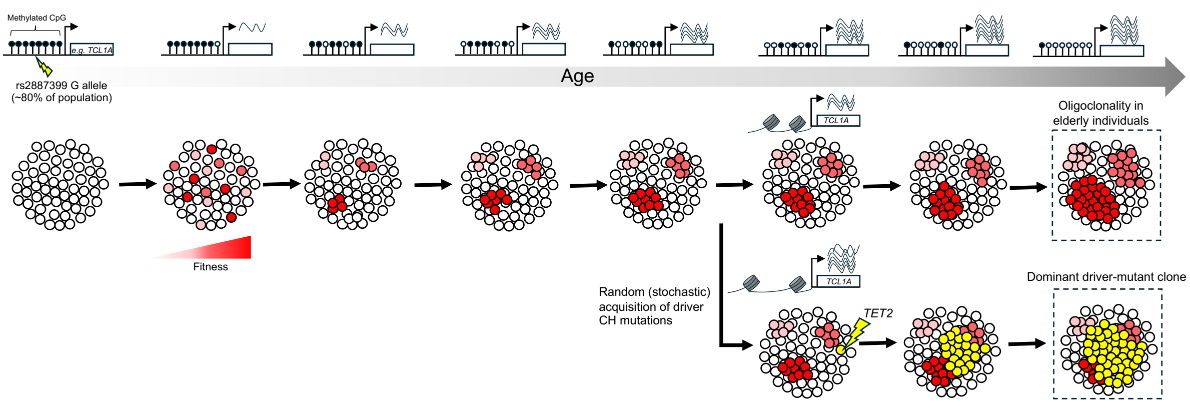

Methylated CpG
e.g. TCL1A
rs2887399 G allele
(~80% of population)
Age
Fitness
TCL1A
Oligoclonality in
elderly individuals
Random (stochastic)
acquisition of driver
CH mutations
TCL1A
TET2
Dominant driver-mutant clone

**b**

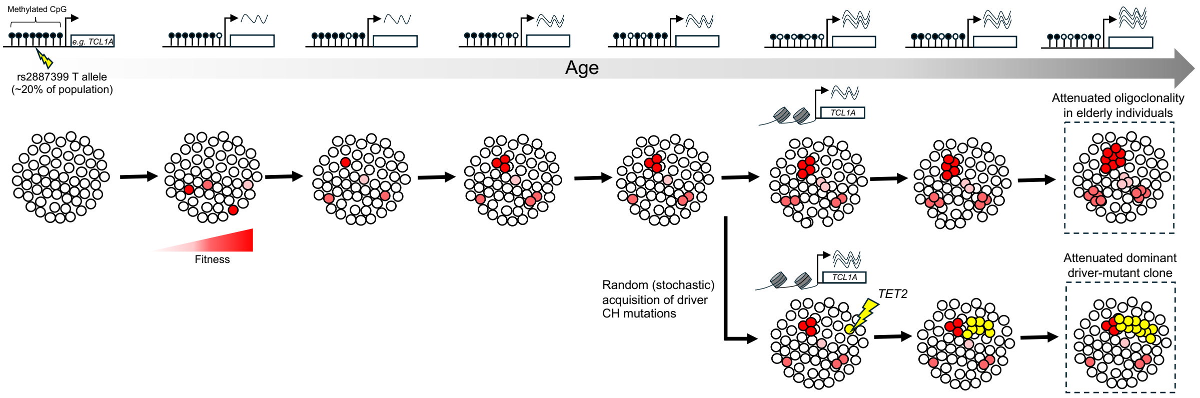

Methylated CpG
e.g. TCL1A
rs2887399 T allele
(~20% of population)
Age
Fitness
TCL1A
Attenuated oligoclonality
in elderly individuals
Random (stochastic)
acquisition of driver
CH mutations
TCL1A
TET2
Attenuated dominant
driver-mutant clone

**Supplementary Figure 22: Proposed model on the shared genetic predisposition and clonal dynamics leading up to the detection of CH-UD and non-*DNMT3A* CH.**

**a,b,** Aging is characterised by the global demethylation of CpG islands that are enriched in promoter regions, including that of *TCL1A* gene[1]. The rate of *TCL1A* promoter demethylation is higher among individuals with the G allele of the *TCL1A* promoter risk genetic variant (rs2887399) relative to the T allele (Supplementary Figures 17a and b). *TCL1A* promoter demethylation is in turn associated with increased *TCL1A* gene expression (Supplementary Figure 17c). Collectively, this is consistent with our observation among UKB participants that TCL1A plasma protein levels increase with age (Figure 5e and Supplementary Figures 15a and b), and the levels are higher among individuals with rs2887399-G relative to -T (Supplementary Figures 14a and b), even in the absence of driver CH mutations. *TCL1A* activation is likely to be stochastic and transient across the HSC population with some cells with high *TCL1A* gene expression while others with low-to-modest *TCL1A* gene expression[2]. This is reminiscent of the presence of multiple 'driverless' clones observed in healthy individuals inferred from WGS of HSC-derived colonies[3,4]. CH-UD becomes increasing pervasive across the general population (Figures 2b-d) as clone sizes and passenger mutation burden increases above the detection threshold of WGS. A subset of individuals may randomly (stochastically) acquire a fitness-enhancing driver mutation, such as *TET2* and *ASXL1*, that increases clonal expansion rate beyond 'driverless' clones (Supplementary Figure 9a). This is reminiscent of the presence of one dominant CH clone observed in >90% of driver CH individuals[5]. This contrasts with oligoclonality (the presence of multiple independent clones) observed in 'driverless' elderly individuals[3]. The most notable inherited genetic risk factor of clonal expansion is rs2887399 where the G allele is associated with higher rate of clonal expansion (Figure 4b and Supplementary Figure 9d). In the presence of *TET2* or *ASXL*1 mutation, the *TCL1A* promoter becomes more accessible when the G allele is also present[2] **(a)**. On the other hand, clonal expansion, and the *TCL1A* promoter accessibility in the presence of *TET2* or *ASXL1* mutation, are attenuated in the presence of the T allele **(b)**. Counterintuitively, the G allele is present in approximately 4 in 5 individuals, suggesting that *TCL1A* activation, and perhaps clonal expansion, may confer fitness/reproductive/evolutionary advantage that outweighs the risk of aging-related phenotypes associated with CH.

### Supplementary Notes

**Supplementary Notes 1: Singleton identification and cohort selection on the basis of ancestry and hypermutant status**

Putative passenger mutations were identified from singletons. A singleton is defined as a genetic variant that is detected only once in the entire study cohort. Singleton may be of germline or somatic origin.

We first identified mutations using a germline mutation caller from whole-genome sequencing data and subsequently applied a set of previously described filters to enrich for singletons of somatic origin[6]. Specifically, we retained variants meeting the following criteria: (1) variant allele frequency ≤25%, (2) variant site supported by 15-60 reads, (3) alternate allele supported by ≥3 reads, and (4) not reported in Genome Aggregation Database (gnomAD; Supplementary Figure 1a). We further retained singletons at sites with genotypes missingness ≤10%, i.e., mutation status was successfully determined at the site where the singleton was detected in >90% of individuals. Singletons at sites with genotypes missingness >10% were enriched for indels, in particular insertion, which are presumed to be sequencing artifacts, and therefore not included in downstream analysis (Supplementary Figures 1b and c). Next, we further retained autosomal variants, but not variants located on sex chromosomes. This is because we observed high rates of genotype missingness across the entire length of chromosomes X and Y (Supplementary Figure 1d).

To confidently classify a genetic variant as a singleton, a large sample size of study cohort is desirable to reduce the fraction of singletons that are rare variants of germline origin. Indeed, we observed lower number of singletons detected with increasing sample size defined by ancestry group (Supplementary Figure 2a-f). Specifically, Europeans, which constitute majority of UKB participants, had the smallest number of singletons detected, followed by South Asians, African, Admixed Americans, and East Asians. Moreover, the current reference genome is essentially a European genome. Therefore, singletons detected in non-European ancestry may have a higher enrichment of rare variants of germline origin. Hence, we only included European individuals in downstream analysis.

Among European individuals, we further excluded individuals with exceedingly high number of singletons. These “hypermutants” were defined as individuals with the number of singletons above the 99.5 percentile (Supplementary Figure 3a-c). Older individuals are expected to have higher number of singletons as passenger mutations accumulate throughout a human's lifespan. In contrast, we observed weaker association between age and frequency of hypermutants compared to non-hypermutants (Supplementary Figure 3d-f). Furthermore, hypermutants demonstrated genomic-related characteristics that defer from non-hypermutants. Firstly, compared to non-hypermutants, hypermutants were depleted for singletons with C>T transition (Supplementary Figure 3g-i). One source of C>T transition is advancing age due to spontaneous deamination of 5-methyl-cytosine[7]. Hypermutants were also enriched for C>A and T>G but depleted of C>G and T>C nucleotide changes. Secondly, compared to non-hypermutants, the alternative allele of singletons of hypermutants were supported by lesser number of reads as reflected by the lower variant allele frequency (Supplementary Figure 3j-l). Thirdly, compared to non-hypermutants, the sites of singletons of hypermutants had lower coverage (Supplementary Figure 3m-o). Collectively, these suggest singletons identified among hypermutants are likely to be enriched for sequencing artifacts compared to non-hypermutants. Therefore, hypermutants were excluded from downstream analysis.

**Supplementary Notes 2: Features of singletons included in machine learning classifiers for identifying CH-UD.**

We identified main features of singletons to be included in the machine learning classifiers for identifying CH-UD. Here, we assessed whether these features were different between driver CH or mCA from non-CH (individuals without driver CH or mCA). We anticipate singletons from driver CH and mCA individuals to be enriched for characteristics of somatic processes while singletons from non-CH individuals to be enriched for characteristics of sequencing artefacts or variants of germline origin.

The first feature is simply the absolute number of singletons. We observed higher number of singletons among driver CH and mCA individuals (mean = 29 and 17, respectively) compared to non-CH individuals (mean = 10; Supplementary Figure 4a). To date, individuals with CH-UD are identified solely on the basis of the absolute number of singletons[6,8]. In this study, we included additional genetic features of singletons, as follows, that may be predictive of driver CH of mCA, and by extension, CH-UD.

The second feature is the fraction of singletons classified as substitution, deletion or insertion. We anticipate indels to be enriched for sequencing artifacts. Compared to non-CH individuals, we observed driver CH and mCA individuals to have higher fraction of singletons consisting of substitution but lower fraction of singletons consisting of indels (Supplementary Figures 4b and c).

The third feature is the fraction of singletons classified based on nucleotide change (C>A, C>G, C>T, T>A, T>C, T>G). We anticipate aging-related nucleotide change to be enriched among driver CH and mCA individuals. Indeed, we observed C>T transitions to be enriched among driver CH and mCA individuals compared to non-CH individuals (Supplementary Figure 4d and se). One source of C>T transition is advancing age due to spontaneous deamination of 5-methyl-cytosine[7]. We also observed T>G to be depleted among driver CH and mCA individuals compared to non-CH individuals. But the source of this depletion is unclear.

The fourth feature is the fraction of singletons classified based on their location on corresponding opened chromatin regions in haematopoietic stem or progenitor cells (HSPCs) or HSCs. These opened chromatin regions were defined using a published ATAC-sequencing dataset[9]. Compared to non-CH individuals, we observed a higher fraction of singletons in driver CH and mCA individuals to be located within opened chromatin regions of HSCs but not HSPCs (Supplementary Figure 4f and g). This may suggest that the singletons detected originated in the stem cells rather than the downstream and more differentiated progenitor cells. Intriguingly, higher fraction of singletons was observed to be located within opened chromatin regions of normal, but not pre-leukaemic, HSCs. This may suggest that passenger mutations that arose early during the course of clonal expansion are more likely to be detected compared mutations that arose later. Nevertheless, we concede that the correlation between chromatin conformation and mutation rates is complex and therefore further mechanistic studies are required to elucidate the relationship between the two[10].

The fifth and final feature is mutation tolerance score of the singletons. We hypothesise that passenger mutations are located on genomic sites that are more tolerable to mutations. Indeed, compared to non-CH individuals, we observed singletons from driver CH and mCA individuals to have higher mutation tolerance score (Supplementary Figure 4h).

Taken together, both driver CH and mCA individuals demonstrated singleton features that were difference from non-CH individuals. In particular, relative to non-CH individuals, the singleton features of driver CH individuals were more distinct than mCA individuals. Collectively, these singletons features may be informative for predicting CH-UD in downstream multi-parametric (machine learning) approach.

**Supplementary Notes 3: Training and testing of machine learning classifiers for identifying CH-UD.**

We first benchmarked three different positive control sets, namely driver CH, mCA, and combined driver CH and mCA. The negative control set was individuals ≤44 years old without driver CH or mCA. As the different WGS batches demonstrated variable sequencing metrices, we performed the benchmarking analysis and subsequent CH-UD prediction for each batch separately (Supplementary Figures 5a-e).

Using random forest, we observed better performance when we trained our classifier on driver CH compared to training on either mCA or combined driver CH and mCA. When applied on the test set consisting of combined driver CH and mCA, random forest demonstrated area under curve (AUC), sensitivity, and negative predictive value (NPV) of 63.1%, 29.9%, and 93.2%, respectively (Supplementary Figure 6a). We noted clone size-dependent performance, with better performance observed for detecting larger clones (AUC, sensitivity, and NPV of 81.5%, 66.9%, and 99.2%, respectively, for cell fraction ≥0.2 versus 54.5%, 12.7%, and 95.6%, respectively, for cell fraction <0.1; Supplementary Figures 6b). Therefore, we proceeded with benchmarking additional machine learning (ML) classifiers using the training set consisting of driver CH.

We benchmarked four different ML classifiers, namely random forest, XGBoost, support vector machine (SVM), and logistic regression. We also included a singleton-only approach for benchmarking (see Methods). We observed the performance metrices to be similar across all four ML classifiers, and all ML classifiers (range of AUC, sensitivity, and NPV of 62.5-63.1%, 28.1-29.9%, and 93.1-93.2%) modestly outperformed the singleton-only approach (overall AUC, sensitivity, and NPV of 60.5%, 27.0%, and 92.8%, respectively; Supplementary Figures 6c and d). Nevertheless, we noted non-overlapping CH-UD cases identified by each classifier. Specifically, 20,258 CH-UD cases were identified by all four ML classifiers, 7,034 CH-UD cases were identified by two or three ML classifiers, while 5,637 CH-UD cases were identified exclusively by one ML classifier (Supplementary Figures 6e). Therefore, beyond these performance metrices, we performed further evaluation of the different ML classifiers.

To this end, CH-UD that were detected using ML classifiers demonstrated better agreement of age-frequency dependency across the different WGS batches compared to the singleton-only approach (Supplementary Figures 6f-j). Compared to other ML classifiers, CH-UD detected using random forest demonstrated the strongest age-frequency association (Supplementary Figure 6k), and a population frequency of ~1% at 40 years old (Supplementary Figure 6g); similar with that for driver CH[11,12]. Based on these collective assessments, we proceeded with CH-UD detected by the random forest classifier for downstream analyses.

**Supplementary Notes 4: Influence of rs2887399 genotype on *TCL1A* promoter methylation.**

We have shown that rs2887399 genotype may influence TCL1A protein levels in the absence of driver CH mutations (Supplementary Figure 14). We now sought to investigate whether rs2887399 genotype may influence *TCL1A* expression, in part, by influencing *TCL1A* promoter methylation.

We noted a CpG island at the 5' end of the *TCL1A* gene (chr14: 95,713,920-95,714,511) and that rs28877399 was located within this CpG island (chr14: 95,714,358; Supplementary Figure 17a). Methylation quantitative trait loci (meQTL) in blood[13] revealed rs28877399 G allele to be associated with decreased methylation at eight CpG sites, seven of which were located within the *TCL1A* CpG island (Supplementary Figure 17b).

Expression quantitative trait methylation (eQTM) in blood revealed demethylation of CpG sites was associated with increased gene expression in general[14]. Specifically, methylation of 15,206 CpG (44%) sites were associated with increased gene expression while demethylation 19,595 (56%) CpG sites were associated with decreased gene expression. Two *TCL1A* CpG sites were identified from this dataset and both sites were associated with increased *TCL1A* expression (Supplementary Figure 17c).

**Supplementary Notes 5: Bayesian hierarchical model of stochastic *TCL1A* activation and clonal expansion.**

*TCL1A* activation contributes to clonal expansion in driver CH[2]. But the role of *TCL1A* activation in clonal expansion in the absence of driver mutations has not been explored. *TCL1A* is expressed at low levels in the HSC compartment, possibly reflecting a transient activation pattern, where CpG sites at promoters stochastically fluctuate between a methylated and an unmethylated state[15]. Furthermore, functional and phenotypic HSCs are rare in whole blood samples. Collectively, shifting gene expression and methylation states of *TCL1A* present challenges to investigate its contribution to HSC dynamics. Therefore, we developed a Bayesian hierarchical model (BHM) of *TCL1A* activation and clonal expansion, and subsequently compared the model's prediction to that observed among UKB participants with TCL1A plasma protein and singleton data available.

We have shown increased VAF-adjusted singleton burden (PACER score) with increased TCL1A plasma protein levels (beta [95% CI] = 1.18 [0.059, 2.30], *P* = 0.039). Our simulation of stochastic *TCL1A* activation demonstrated general agreement between the model posterior predictive distributions and observed distributions for singleton burden across the different age groups with predicted mean of 40.51 versus observed mean of 40.59 (predictive p-value (PPP) = 0.50; Supplementary Figure 18a-e). The model posterior predictive distributions were also in general agreement with the observed distributions for singleton-derived VAF with predicted mean of 0.52 versus observed mean of 0.54 (predictive p-value (PPP) = 0.15; Supplementary Figure 18f-j). As reference, a PPP value of 0.5 indicates a model perfectly centred on the data, whereas the range 0.05-0.95 is typically considered a “good” fit. These suggest stochastic *TCL1A* activation was associated with clonal expansion among CH-UD individuals.

We further benchmarked our BHM against null models, namely a fully neutral model and one with a non-decaying TCL1A deactivation rate. The fully neutral model was very strongly rejected, while the model with a non-decaying TCL1A deactivation rate underperformed and was also rejected. Furthermore, the inferred decay rate parameter in the main BHM excluded zero, providing further evidence against the non-decaying deactivation rate model.

**Supplementary Notes 6: Benchmarking Mutect2-detected *CALR* indels and pileup-detected *JAK2* V617F variants.**

In this study, we performed driver CH variant calling in accordance with the consensus driver CH gene list and using somatic mutation caller (Mutect2) proposed by the Trans-Omics for Precision Medicine (TOPMed) programme[16]. To investigate whether the association between CH-UD and essential thrombocythaemia (ET) may be explained by somatic variants not captured by the current consensus driver CH detection workflow, we proceeded to identify *CALR* indels using Mutect2 and additional *JAK2* V617F variants using pileup.

*Bona fide* somatic variants are expected to demonstrate an increase in carrier frequency with older age. Here, we observed *CALR* mutation carrier frequency increased with age (Supplementary Figure 19a). For *JAK2* V617F, we observed clones with variant allele frequency (VAF) of ≥3% supported by ≥1 mutant read demonstrated increased mutation carrier frequency with older age (Supplementary Figure 19b and c).

Polycythaemia vera (PV) is almost exclusively associated with *JAK2* V617F[17]. Here, we observed *JAK2* V617F to be associated with PV. Notably, individuals with ≥1 mutant read were associated with PV, even for small clones (<3% VAF: OR [95% CI] = 4.24 [1.22,14.8]; $P_{\lambda\text{-adjusted}}$ = 0.066); Supplementary Figure 19d). Approximately 50-60% and 20-25% of ET cases are associated with *JAK2* V617F and *CALR*, respecitvely[17]. Here, we observed both *JAK2* V617F and *CALR* indels to be associated with ET (Supplementary Figure 19e). Platelet crit is a feature associated, but not exclusively, with ET. Here, we observed both *JAK2* V617F and *CALR* indels to be associated with platelet crit (Supplementary Figure 19f).

**Supplementary Notes 7: Association between CH-UD versus essential thrombocythaemia and platelet crit after removing *CALR* indels and residual *JAK2* V617F variants.**

We hypothesise that the association between CH-UD versus essential thrombocythaemia (ET) and platelet crit may be explained, in part, by hitherto undetected *CALR* indels and *JAK2* V617F variants.

We observed CH-UD to be associated with polycythaemia vera (PV), a disease almost exclusively associated with *JAK2* V617F (OR [95% CI] = 2.08 [1.37,3.14]; $P_{\lambda\text{-adjusted}}$ = 0.016). CH-UD was no longer associated with PV after removing individuals with ≥1 read supporting *JAK2* V617F (OR [95% CI] = 1.59 [0.98,2.57]; $P_{\lambda\text{-adjusted}}$ = 0.20; Supplementary Figure 20a). This suggests that majority of hitherto undetected *JAK2* V617F carriers were removed from CH-UD individuals. The association between CH-UD and ET was attenuated, but remained statistically significant at Bonferroni threshold, after excluding *CALR* indels and *JAK2* V617F carriers (OR [95% CI] = 4.31 [3.24,5.73]; $P_{\lambda\text{-adjusted}}$ = 3.15 x $10^{-12}$; Supplementary Figures 20b). The association between CH-UD and platelet crit was similarly attenuated, but remained statistically significant at Bonferroni threshold, after excluding *CALR* indels and *JAK2* V617F carriers (beta [95% CI] = 0.11 [0.10,0.13]; $P_{\lambda\text{-adjusted}}$ = 3.49 x $10^{-29}$; Supplementary Figures 21a). It is noteworthy that the attenuation of ET and platelet crit effect sizes was not likely due to reduction in CH-UD sample size after removing these mutation carriers but rather this is due the elimination of *CALR* indels and additional *JAK2* V617F mutation carriers from CH-UD individuals. This is because the effect size of phenotypes other than ET and platelet crit were not attenuated after versus before removal of *CALR* indels and additional *JAK2* V617F from CH-UD individuals (Supplementary Figures 20c and 21b).

Collectively, these suggest that the association between CH-UD versus ET and platelet crit may not be explained entirely by hitherto undetected *CALR* indels and *JAK2* V617F variants.
